# Development of an explicit and implicit knowledge identification tool for the analysis of the decision-making process of traditional Asian medicine doctors

**DOI:** 10.1101/2021.12.13.21267754

**Authors:** Musun Park, Min Hee Kim, So-young Park, Minseo Kang, Inhwa Choi, Chang-Eop Kim

## Abstract

**Background and objectives:** While pattern identification (PI) is an essential process for diagnosis and treatment in traditional Asian medicine (TAM), it is difficult to objectify since it relies heavily on implicit knowledge. Here, we propose a machine learning-based analysis tool to objectify and evaluate the clinical decision-making process of PI in terms of explicit and implicit knowledge.

**Methods:** Clinical data for the development of the analysis tool were collected using a questionnaire administered to allergic rhinitis (AR) patients and the diagnosis and prescription results of TAM doctors based on the completed AR questionnaires. Explicit knowledge and implicit knowledge were defined based on the explicit and implicit importance scores of the AR questionnaire, which were obtained through doctors’ explicit scoring and feature evaluations of machine learning models, respectively. The analysis tool consists of eight evaluation indicators used to compare, analyze and visualize the explicit and implicit knowledge of TAM doctors.

**Results:** The analysis results for 8 doctors showed that our tool could successfully identify explicit and implicit knowledge in the PI process. We also conducted a postquestionnaire study with the doctors who participated to evaluate the applicability of our tool.

**Conclusions:** This study proposed a tool to evaluate and compare decision-making processes of TAM doctors in terms of their explicit and implicit knowledge. We identified the differences between doctors’ own explicit and implicit knowledge and the differences among TAM doctors. The proposed tool would be helpful for the clinical standardization of TAM, doctors’ own clinical practice, and intern/resident training.

## 1. Introduction

Traditional Asian medicine (TAM) is a medicine that has traditionally been practiced in Asia over thousands of years and mainly includes traditional Chinese medicine, traditional Japanese medicine and traditional Korean medicine. With accumulated evidence of its efficacy and safety, TAM is widely used for various diseases. In TAM, pattern identification (PI) is the essential process in diagnosis and treatment. PI is a tool of TAM that results in a diagnostic conclusion given a cluster of concurrent symptoms and signs [1]; however, the process of collecting information is based on patients’ subjective description and doctors’ subjective assessment. Therefore, the standardization and objectification of PI is needed to improve the quality of medical care in TAM. Standardization and objectification reduce variation in clinical treatment and patient outcome and thus improve the quality of patient care in many areas of medicine [2-3]. From these perspectives, there have been many trials to standardize PI for many diseases [4-6]. However, these trials have demonstrated the considerable heterogeneity of the decision-making process between clinicians, and this heterogeneity makes it challenging to standardize and objectify PI.

Knowledge used in doctors’ clinical decision-making processes can be classified into two different categories: explicit and implicit knowledge. Explicit knowledge refers to knowledge that can be expressed in words and in a form that can be codified. In contrast, implicit knowledge refers to knowledge that an individual obtains through experience but does not convey to others [7-8]. While explicit knowledge can be simply evaluated via interview or questionnaire, implicit knowledge is difficult to assess. It is well known that the clinical decision-making process is based on implicit knowledge [9-10]. That is, the same patient can be assessed through a different PI process because each doctor has different implicit knowledge. Therefore, to overcome the heterogeneity of the decision-making process, a method of extracting and evaluating each doctor’s implicit knowledge will be necessary.

One way to overcome this problem is to use machine learning (ML) techniques to extract quantitative information on implicit knowledge from knowers. With the supervised learning-based method, it is possible to identify intrinsic patterns that cannot be expressed with explicit knowledge [11]. By learning the supervised learning model using the clinical feature and by analyzing the model’s intrinsic pattern, it will be possible to extract the implicit knowledge applied to the medical field.

To the best of our knowledge, there are many cases in which ML has been applied to clinical decision-making processes [12-13], but no research has been conducted to understand implicit knowledge in clinical decision-making processes. Our study extracted and evaluated implicit knowledge in clinical decision-making for the first time. Although PI for allergic rhinitis (AR) was studied, this methodology could also be applicable for universal clinical decision-making studies. The methodology can reveal the relationship between clinical knowledge and processes in practice and further improve the curriculum of clinicians and students.

In this study, we propose a ML-based analysis tool to evaluate the clinical decision-making process of PI in terms of explicit and implicit knowledge. For a proof of concept, we developed our tool based on diagnosis and prescription data for AR. Explicit knowledge and implicit knowledge were operationally defined based on the relevant scores of the AR questionnaire, which were obtained from doctors’ explicit scoring and machine-learning-based evaluation of each feature, respectively. We developed a method to efficiently compare, analyze and visualize explicit and the implicit knowledge that contributes to doctors’ clinical decision-making processes. We also conducted a postquestionnaire study for the doctors who participated to evaluate the applicability of our tool.

## 2. Materials and Methods

### 2.1. Study design and ethics

We aimed to explore a disease with high prevalence, and we had previously developed a PI questionnaire for AR, so we chose AR as the topic disease in this study. The Allergic Rhinitis Diagnostic Analysis Assessment Tool was created via the four steps described below (Figure 1). This study was approved by the Institutional Review Board of Kyung Hee University Hospital at Gangdong (KHNMC-OH-IRB 2018-01-001). Written informed consent was obtained by the investigator from all participants prior to enrollment.

**Figure 1.**
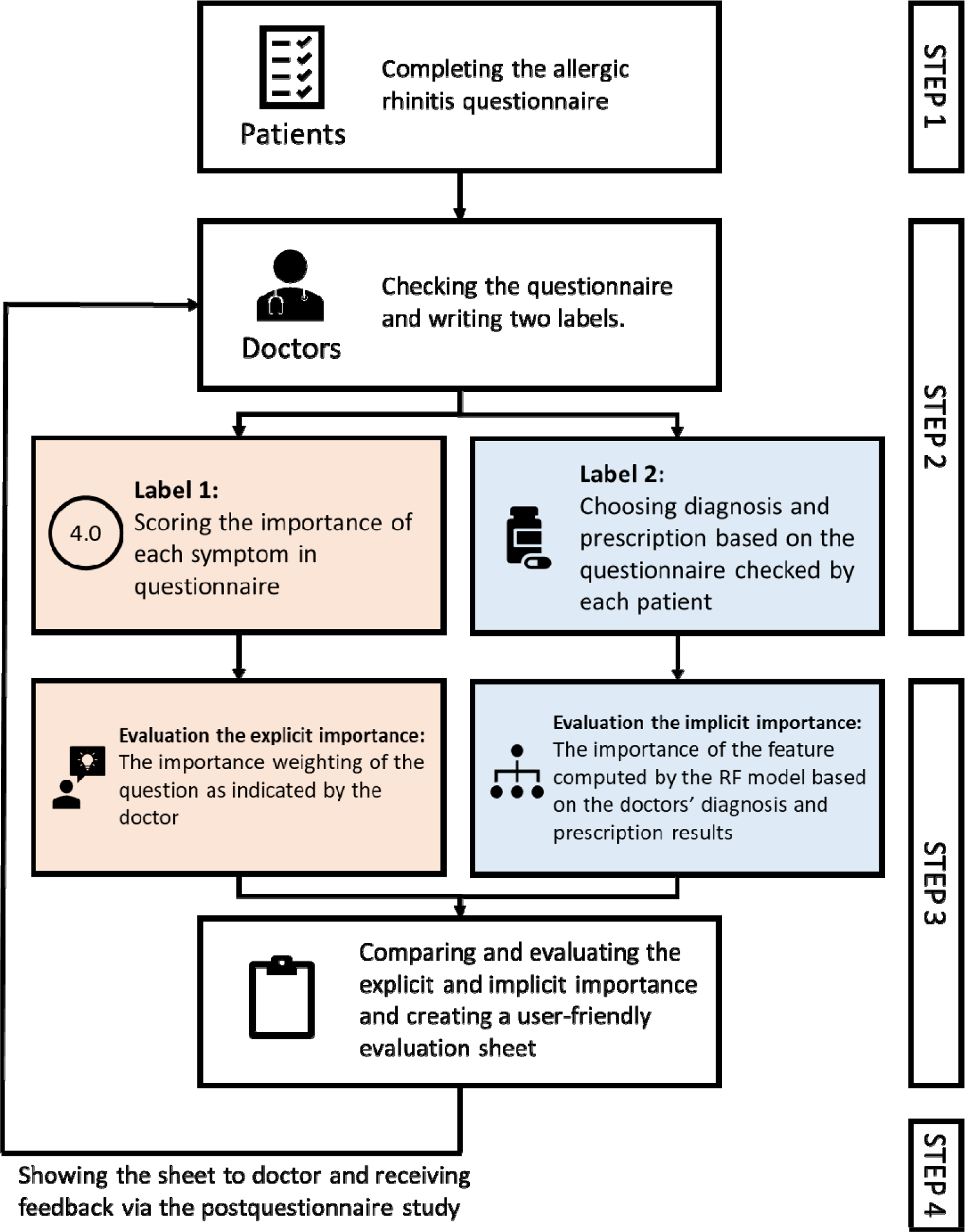
The framework of the Allergic Rhinitis Diagnostic Analysis Assessment Tool. RF, Random Forest

### 2.2. Participants, sample size, and the questionnaire (Step 1)

The inclusion criteria were as follows: (1) age 15-65 years; (2) presence of two or more nasal symptoms (rhinorrhea, nasal congestion, nasal itching and sneezing); (3) presence of nasal symptoms for over two consecutive years; and (4) positive reaction to one or more allergens as evaluated in the skin prick test. The exclusion criteria were as follows: (1) presence of rhinosinusitis of upper respiratory infections; (2) participation in another clinical study within the past 3 months; and (3) clinical judgment suggesting that the individual’s physical or mental status was not appropriate for the study protocol. This study was not intended to validate the effects of certain interventions on AR but rather to explore the explicit and implicit knowledge of doctors. Moreover, there were no comparable studies to refer to for the calculation of the sample size. Therefore, in this exploratory study, we determined a required sample size of 100, which enabled the use of the RF model, with expert opinions in otorhinolaryngology and statistics. The gender ratio in each group was 1:1. We used the PI questionnaire developed from previous studies (Supplementary Table 1) [14-18].

### 2.3. Labeling by doctors (Step 2)

A total of eight TAM doctors who had various levels of clinical experience and who were TAM otolaryngologists or TAM otolaryngology residents were included in this study (Table 1). First, they scored each symptom on the PI questionnaire according to its degree of importance for diagnosing and treating AR patients (label 1) on a 5-point Likert scale (0=Not at all important, 1=Slightly important, 2=Moderately important, 3=Very important, 4=Extremely important). Second, they chose a PI diagnosis and herbal prescription based on the PI questionnaire completed by each patient (label 2) among the presented options. We presented three PI options (lung-heat pattern, lung-cold pattern or spleen qi deficiency pattern) [14] and the five herbal prescription options that are most frequently used for patients with AR in Korea (Galgeun-tang, Socheongryong-Tang, Hyeonggaeyeongyo-tang, Yeotaektonggi-tang, and Bojungikgi-tang) [19-20].

**Table 1.**
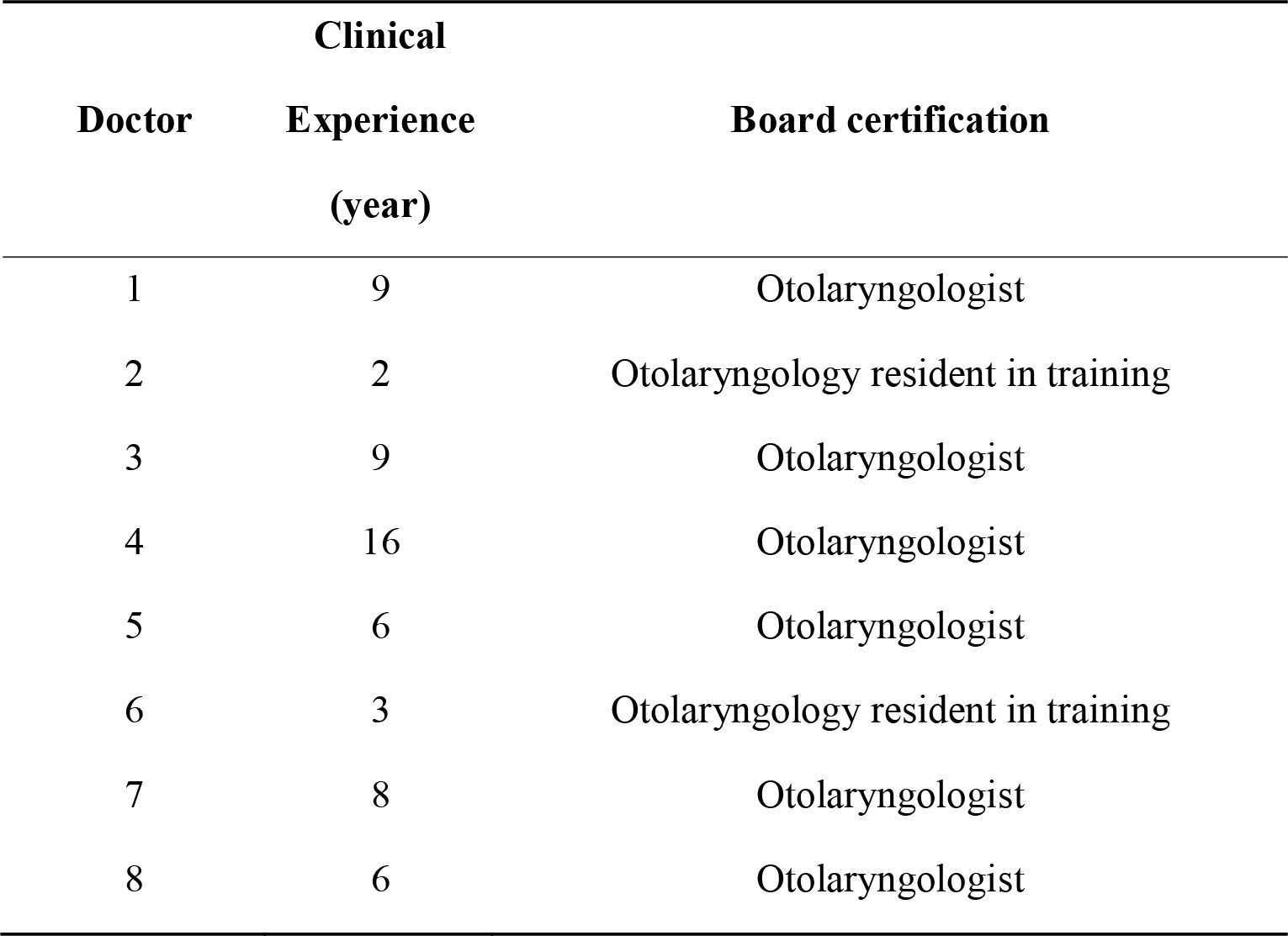
Characteristics of doctors

### 2.4. Evaluation of explicit and implicit knowledge (Step 3-1)

To assess explicit and implicit knowledge in the clinical decision-making process, we first defined the explicit and implicit importance scores for the features. The explicit importance score was defined for the features obtained from doctors’ label 1 task (indicating the degree of importance of each symptom for diagnosing and treating AR patients). Each importance score was normalized so that the total possible was importance score was 100.

Implicit importance was defined based on the weights of the features learned by the machine learning algorithm when the learning algorithm effectively reproduced the doctor’s decision-making process. To calculate the implicit importance score, we adopted a random forest (RF) model that could effectively capture the complex, nonlinear relationships of the datasets while providing relatively transparent information about the model compared to that of more complex state-of-the-art models such as deep neural networks [21-22]. In the training process of the RF classifier, the PI questionnaire and diagnosis/prescription results were used as features and labels, respectively. Then, the implicit importance score was evaluated as the feature importance of the RF classifier. The hyperparameters of the RF model were selected by the grid search algorithm (max_depth = 6, n_estimator = 50). The implicit importance score was evaluated for each feature by creating 100,000 RF models and summing all of the calculated feature importance values from each model. The computed feature importance values were also normalized to 100 for comparison to the explicit importance score. Python, scikit-learn library [23] and Cytoscape software [24] were used for the construction of the RF model and the calculation of the feature importance.

### 2.5. Development of the tool (Step 3-2)

Based on the above definition and evaluation methods of doctors’ explicit and implicit knowledge, we developed a tool to evaluate and compare the explicit and implicit knowledge of otolaryngologists. The tool consists of eight evaluation indicators (three main indicators and five supplemental indicators) derived from six importance scores. (Tables 2 and 3).

**Table 2.** Lists of importance scores

| Label | Type of importance score | Calculation method |
| --- | --- | --- |
| 1 | Each doctor's explicit importance score | Scoring the importance of each symptom on the PI<sup>1</sup> questionnaire |
| 2 | Each doctor's implicit importance score for diagnosis | Calculating the feature importance score from the RF<sup>2</sup> classifier based on the doctor's diagnosis results |
| 3 | Each doctor's implicit importance score for prescription | Calculating the feature importance score from the RF classifier based on the doctor's prescription results |
| 4 | Average explicit importance score among all doctors | Scoring the importance of each symptom on the PI questionnaire |
| 5 | Average implicit importance score for diagnosis among all | Calculating the feature importance score from the RF |

|  | doctors | classifier based on the doctor's diagnosis results |
| --- | --- | --- |
| 6 | Average implicit importance score for prescription among all doctors | Calculating the feature importance score from the RF classifier based on the doctor's prescription results |
| <sup>1</sup> Pattern identification; <sup>2</sup> Random forest |  |  |

**Table 3.** Lists of indicator labels

| Indicator label | Evaluation indicator |
| --- | --- |
| Main 1 (M1) | Comparison of the individual and average explicit and implicit importance scores |
| Main 2 (M2) | Intraindividual correlation of the explicit and implicit importance score |
| Main 3 (M3) | Radar chart analysis |
| Supplementary 1 (S1) | Intraindividual comparison of the explicit and implicit importance scores for diagnosis |
| Supplementary 2 (S2) | Intraindividual comparison of the explicit and implicit importance scores for prescription |
| Supplementary 3 (S3) | Comparison of the individual and average explicit importance scores |
| Supplementary 4 (S4) | Comparison of the individual and average implicit importance scores for diagnosis |
| Supplementary 5 (S5) | Comparison of the individual and average implicit importance scores for prescription |

### 2.6. Postquestionnaire study (Step 4)

To evaluate the tool’s utility in practical settings, all doctors participated in the study answered a post-questionnaire (Supplementary Table 2). The questionnaire included questions about the appropriateness of the tool, the convenience or difficulty of data collection, the potential impact of the tool on education and clinical treatment improvements, the limitations of the tool and the overall satisfaction with the tool and let the all doctors freely comment if they want.

## 3. Results

### 3.1. Construction of the tool (Table 3 and supplementary material 1)

First, to provide an overview of the differences in all importance scores, the evaluation indicator Main 1 (Supplementary material 1, M1) was visualized with line graphs that consisted of all six scores. Scores were sorted in descending order according to the average explicit importance score of average results (Figure 2).

**Figure 2.**
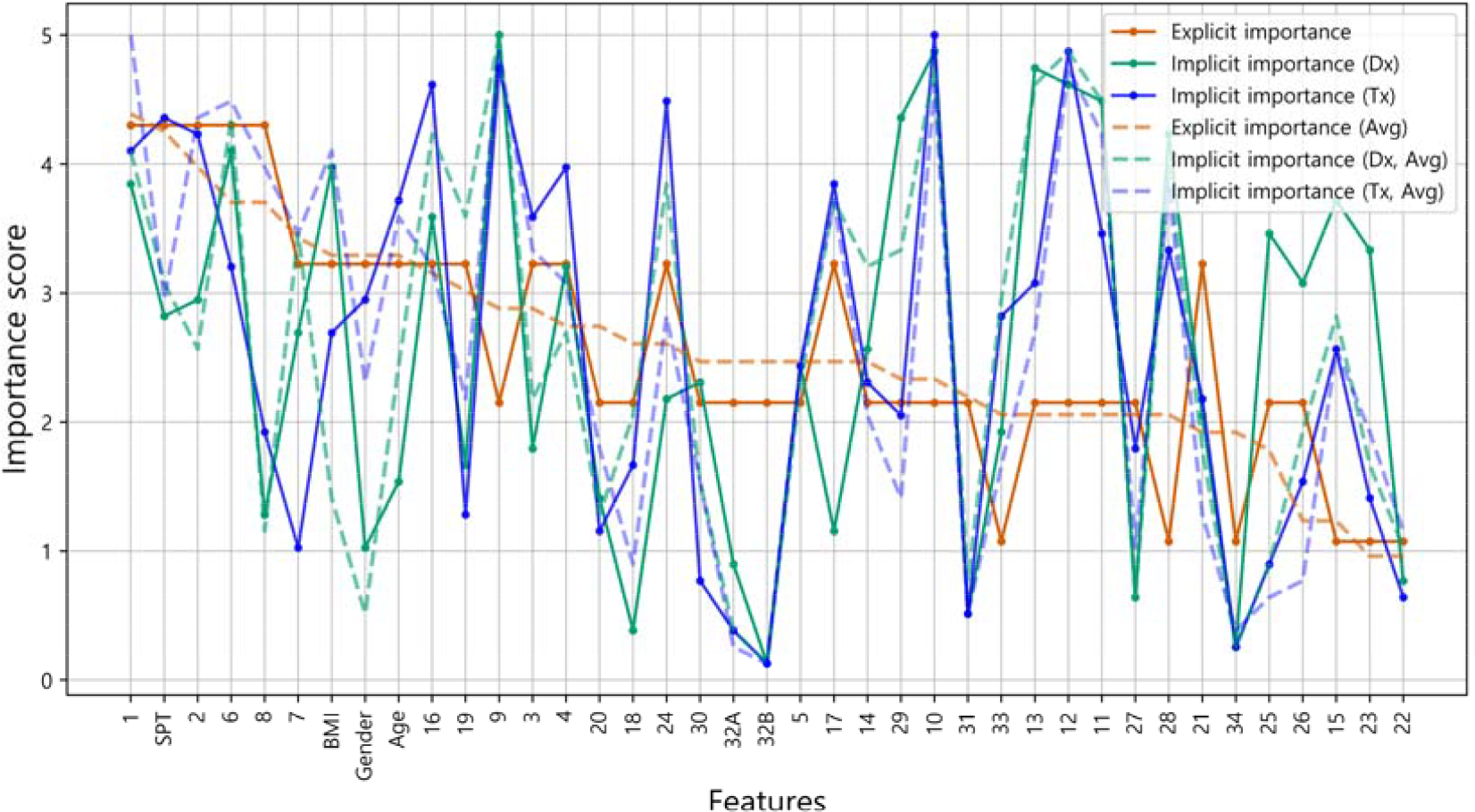
Comparison of the individual and average explicit and implicit importance scores (representative example). The explicit importance score, implicit importance score for diagnosis, and implicit importance score for prescription are visualized in orange, cyan, and blue, respectively. Individual results from each doctor and the average of all doctors’ results were expressed as a solid line and a dotted line, respectively. The graph was sorted in descending order according to the average explicit importance score. Dx, Diagnosis; Tx, prescription (treatment); Avg, Average.

To examine the overall correlation between the explicit and implicit knowledge of individuals, Main 2 (Supplementary material 1, M2) was presented as nodes and edges, which represented the types of importance scores (explicit importance score, implicit importance score for the diagnosis, and implicit importance score for the prescription) and the correlations between importance scores, respectively. The correlations were calculated using Spearman’s rank correlation coefficient (Figure 3) [25].

**Figure 3.**
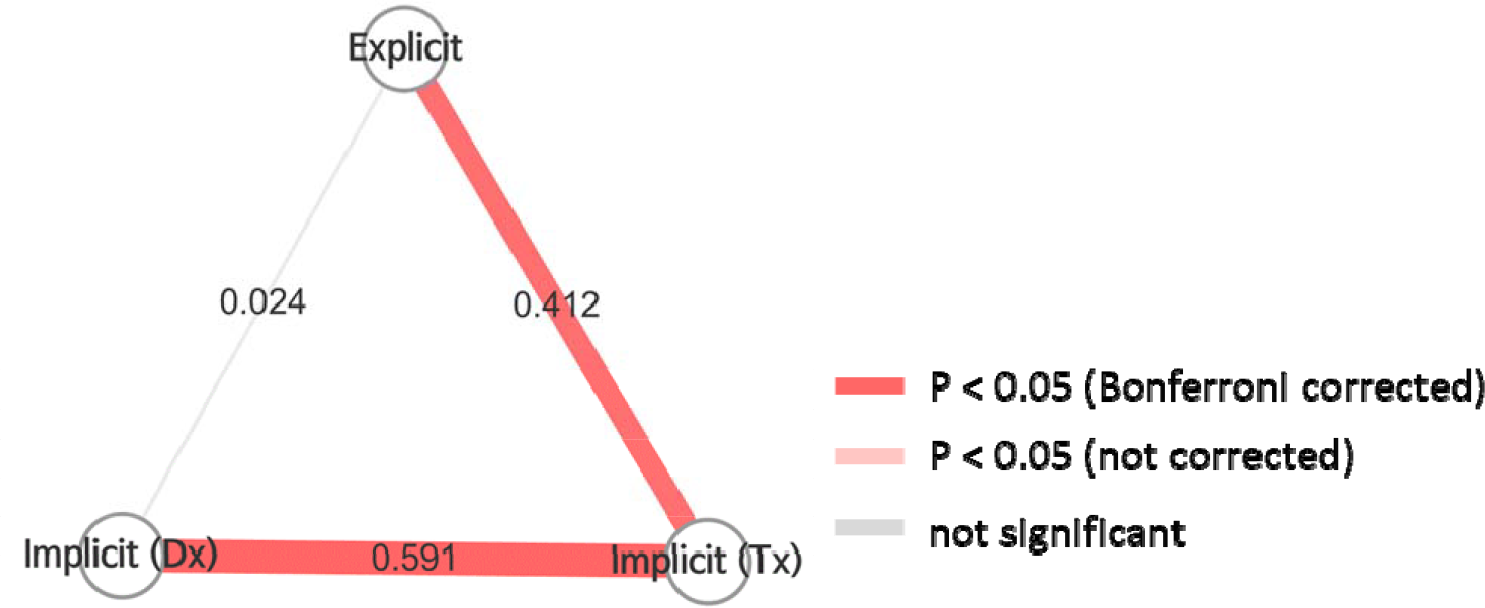
Intraindividual correlation of explicit and implicit importance scores (representative example). Nodes and edges represent the types of importance scores and the correlations between importance scores, respectively. The thickness of the edge indicates the Spearman’s coefficient (noted in the center of the edge). The color of the edge represents the significance of the correlation. Dx, diagnosis; Tx, prescription (treatment).

**Figure 4.**
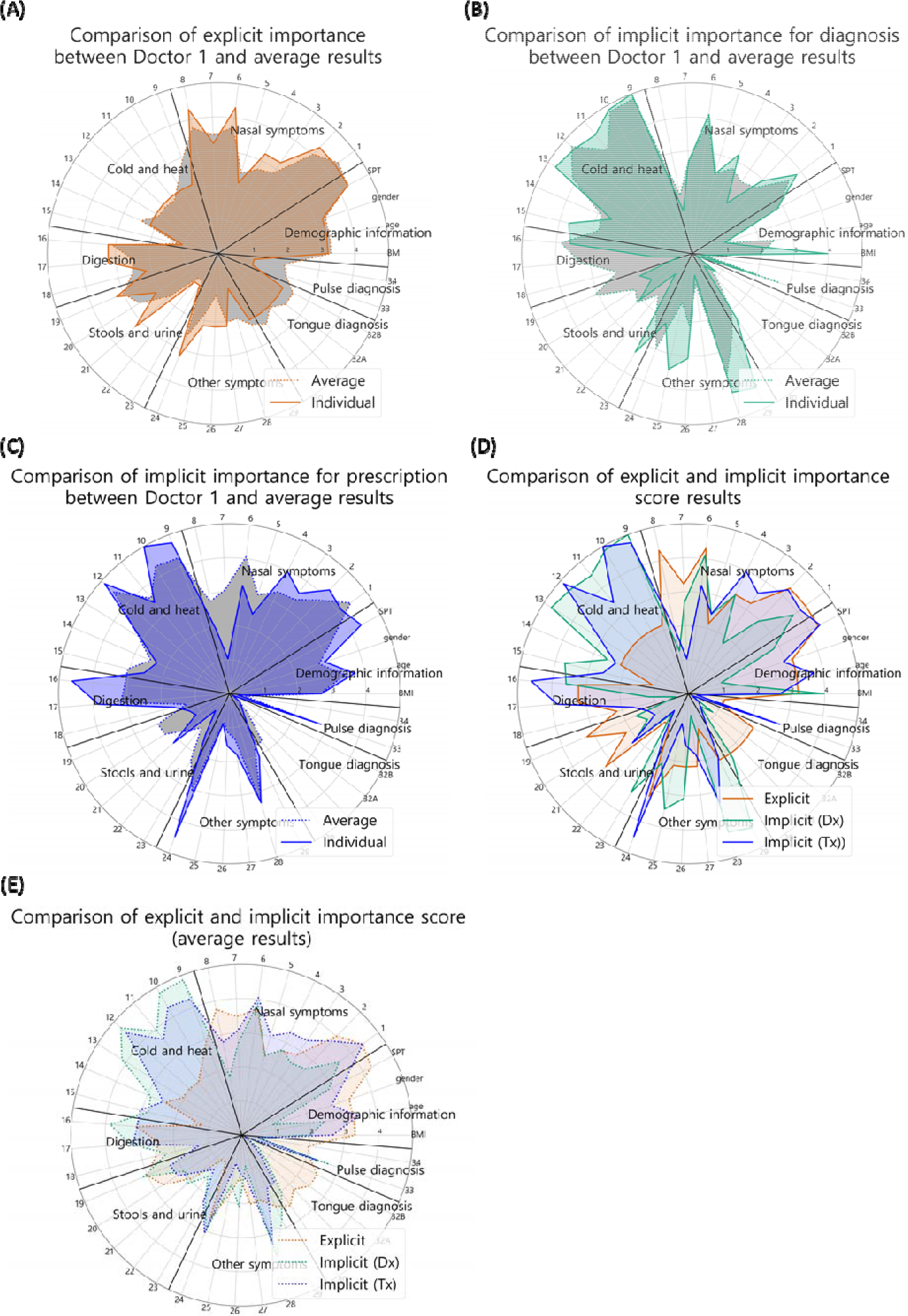
Radar chart analysis (representative example). Explicit importance score, implicit importance score for diagnosis, and implicit importance score for prescription were visualized in orange, cyan, and blue, respectively. Individual results from each doctors and average of all doctors’ results were represented as a solid line and a dotted line, respectively. The radar chart was composed of five graphs: (A) Comparison of explicit importance score between individual and average results; (B) Comparison of implicit importance score for diagnosis between individual doctor and average results; (C) Comparison of implicit importance score for prescription between individual and average results; (D) Comparison between explicit and implicit importance score results; (E) Comparison between explicit and implicit importance score (average results). Dx, diagnosis; Tx, treatment (prescription) In Main 3 (Supplementary material 1, M3), all the features used in the analysis were classified into eight categories and visualized as a radar chart to provide the information about the feature categories that the doctor referred to for treatment. The categories consisted of demographic features of the patient, nose symptoms, cold-hot, digestion, feces and urine, tongue diagnosis, pulse diagnosis and other symptoms (Figure 4).

In Supplementary material 1, Supplementary 1 and 2 (S1 and S2) were represented as scatter plots. The correlation statistics between the explicit and implicit scores were calculated using Spearman’s rank correlation coefficient test (Figure 5). Supplementary 3, 4 and 5 (S3, S4 and S5, respectively) were constructed to compare the average importance score of the doctors as a group with the individual doctor’s importance score (Figure 6). S3, S4 and S5 represented explicit importance scores, implicit importance scores for diagnosis and implicit scores for prescription, respectively. In the same way as the above analysis, the individual and average doctor scores were compared using Spearman’s rank correlation coefficient test.

**Figure 5.**
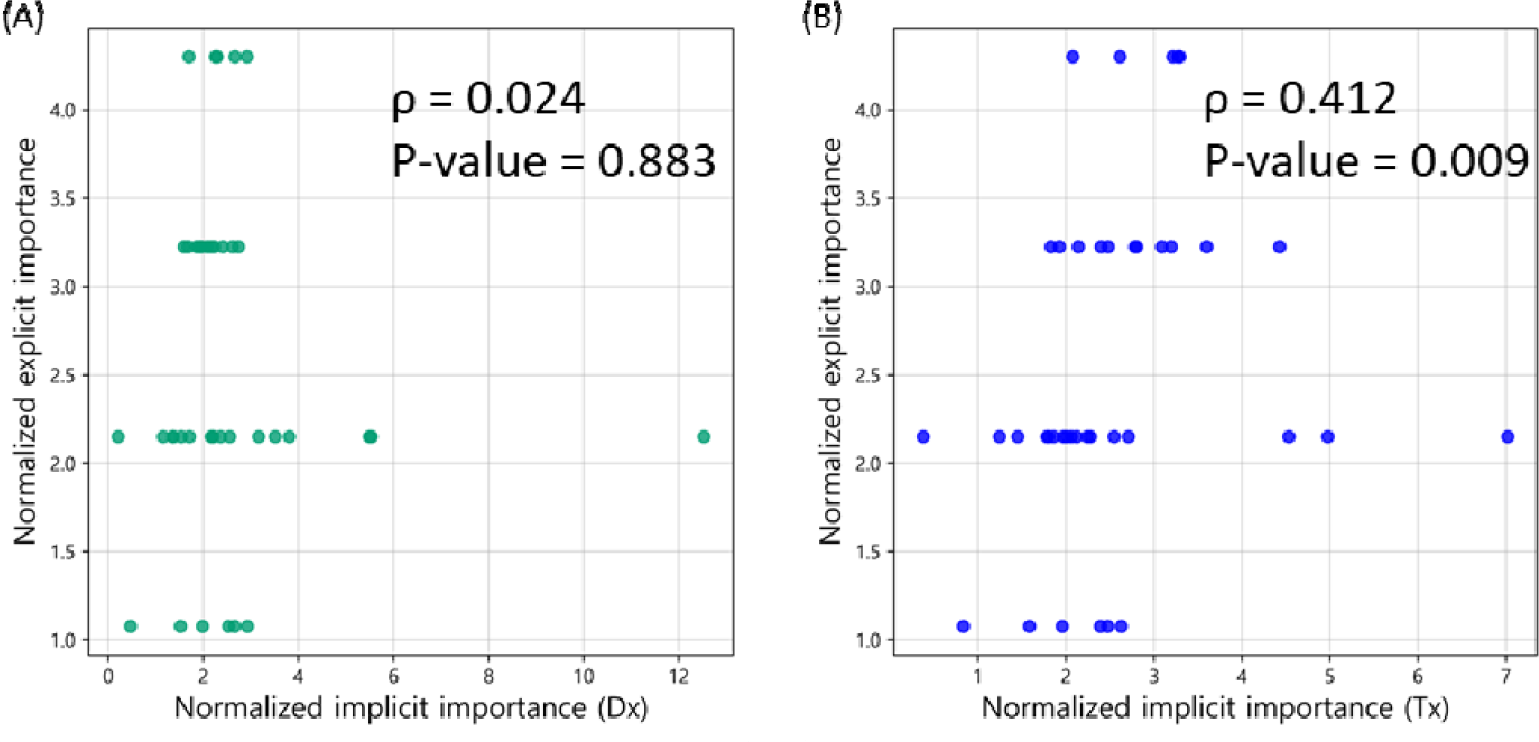
Intraindividual comparison of explicit and implicit importance scores (representative example). Implicit importance scores for diagnosis and prescription are visualized in cyan and blue, respectively. Both graphs are sorted and visualized in ascending order based on the implicit importance score. Dx, Diagnosis; Tx, Treatment (prescription)

**Figure 6.**
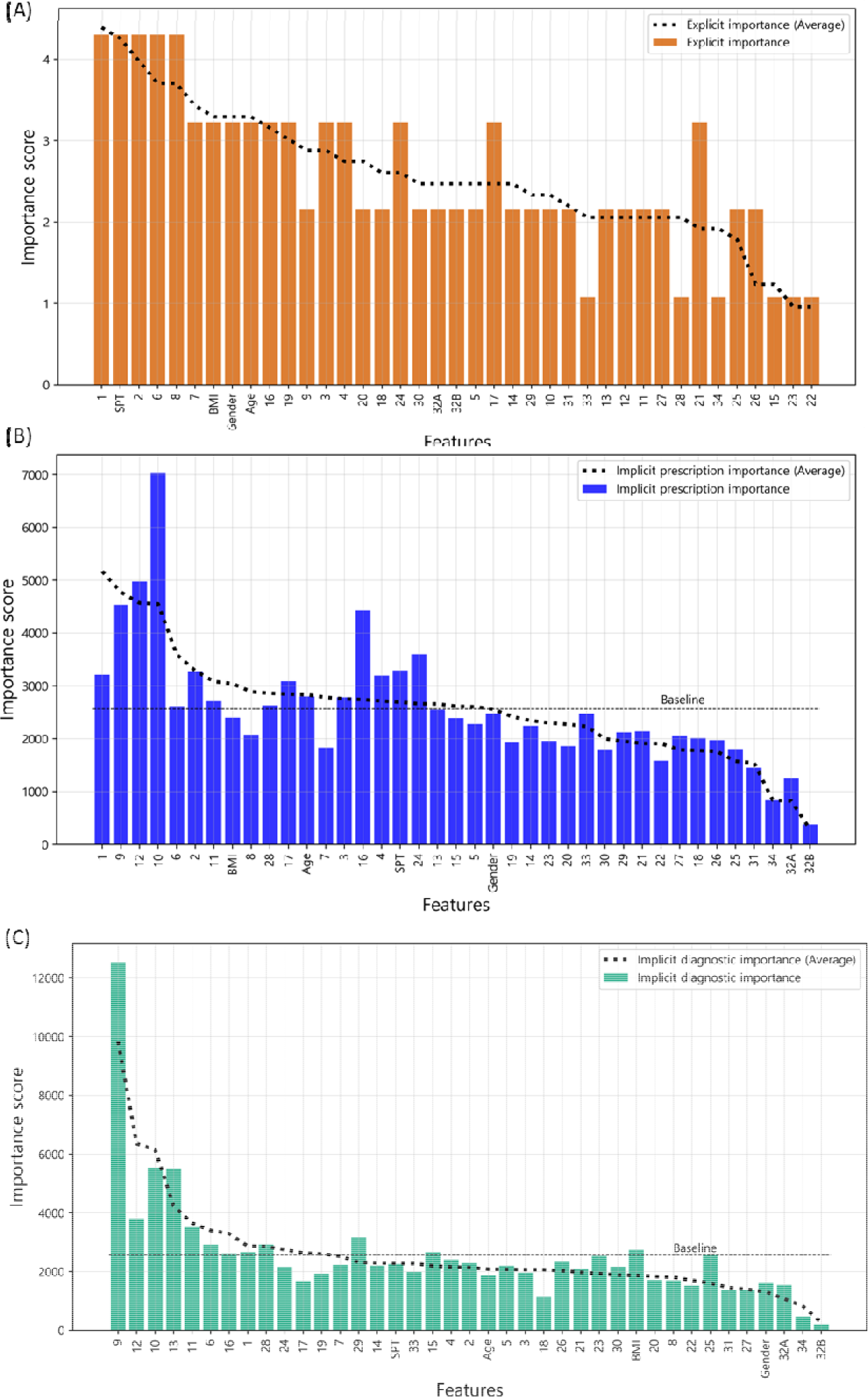
Comparison of importance score between individual and average results (representative example). Explicit importance score, implicit importance score for diagnosis, and implicit importance score for prescription were visualized in orange, cyan, and blue, respectively. Individual results from each doctors and average of all doctors’ results were expressed as a bar graph and a dotted line, respectively. The graphs were sorted in descending order according to the importance score of average results. Baseline is the average value of implicit importance score of individual results. (A) Comparison of explicit importance score between individual and average results; (B) Comparison of implicit importance score for diagnosis between individual and average results; (C) Comparison of implicit importance for prescription between individual and average results.

### 3.2. Pilot analysis using the developed assessment analysis tool

Next, we conducted a pilot analysis using the results of the tool for several otolaryngologists to ensure that our tool provides useful information. We found some interesting results from the analysis as follows.

#### 3.2.1. Comparison of doctors’ average explicit and implicit importance scores

We calculated explicit and implicit importance scores with the data obtained from the patient and physician surveys. The average importance scores of the eight doctors involved in the study were computed for each feature. (Figure 7). The tool showed that the average explicit importance score and the average implicit importance score for diagnosis were not correlated (ρ = 0.18, p-value = 0.272). On the other hand, the explicit importance score and implicit importance score for prescription were weakly correlated (ρ = 0.511, p-value = 0.001). There was a strong correlation between the two implicit importance scores (ρ = 0.732, p-value < 0.001). These results suggest that the explicit importance score can explain the process of prescription better than the process of diagnosis based on our proposed tool.

**Figure 7.**
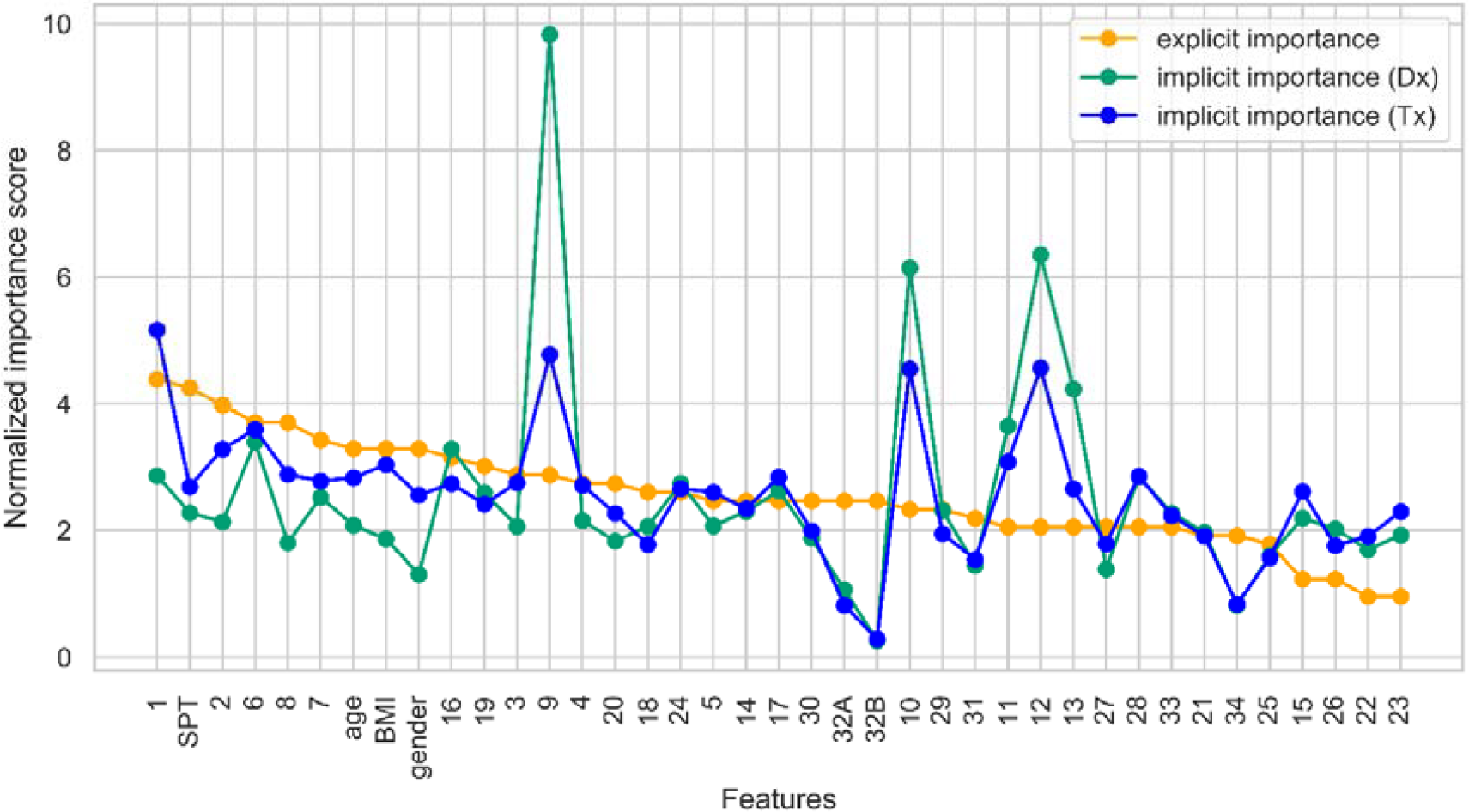
Comparison between doctors’ average explicit and implicit importance scores. The features are sorted in descending order of the explicit importance score. For an equal comparison of the importance scores, each importance score is normalized so that the total possible importance score was 100. Orange color indicates the explicit importance score, cyan color indicates the implicit importance score for diagnosis, and blue color indicates the implicit prescription importance. The features can be found in Supplementary table 2. Dx, diagnosis; Tx, prescription (treatment); SPT, skin prick test.

#### 3.2.2. Intra-individual correlation of explicit and implicit importance score

The correlations of the importance scores of the eight doctors involved in the study were compared and evaluated (Table 4). In the comparison between implicit importance scores for diagnosis and prescription, six doctors showed a significant correlation (p<0.05, Bonferroni corrected), and the other two also showed a correlation, but not at a significant level (p<0.05, uncorrected). There was no significant correlation between explicit and implicit importance scores for most of the doctors (7/8).

**Table 4.** Intra-individual correlation of explicit and implicit importance score

| Doctors | 1 | 2 | 3 | 4 | 5 | 6 | 7 | 8 |
| --- | --- | --- | --- | --- | --- | --- | --- | --- |
| Exp<sup>1</sup> & Imp<sup>2</sup> | 0.024 | 0.451 | -0.142 | -0.028 | 0.052 | 0.152 | 0.232 | 0.254 |
| (Dx<sup>3</sup>) | (0.883) | (0.004*) | (0.39) | (0.864) | (0.755) | (0.357) | (0.154) | (0.119) |
| Exp & Imp | 0.412 | 0.538 | -0.059 | 0.422 | 0.198 | 0.189 | 0.203 | 0.412 |
| (Tx<sup>4</sup>) | (0.009) | (<0.001*) | (0.723) | (0.007) | (0.228) | (0.250) | (0.215) | (0.009) |
| Imp (Dx) | 0.591 | 0.638 | 0.717 | 0.343 | 0.531 | 0.877 | 0.723 | 0.355 |
| & Imp (Tx) | (<0.001*) | (<0.001*) | (<0.001*) | (0.033) | (<0.001*) | (<0.001*) | (<0.001*) | (0.026) |

The numbers in parentheses indicate adjusted p-values. \*, statistically significant (Bonferroni-corrected $p < 0.00625$ ). <sup>1</sup> Explicit importance score; <sup>2</sup> Implicit importance score, <sup>3</sup> diagnosis; <sup>4</sup> prescription (treatment).

#### 3.2.3. Comparison between averaged explicit and implicit importance score of doctors

Radar chart analysis was used to determine how the explicit and implicit importance scores differed by category (Table 5). Through the calculation of the area of the doctors’ average radar chart (Supplementary material 1, Comparison of the explicit and implicit importance scores for diagnosis and the doctors’ average implicit importance score for prescription), the weights of the feature category were identified. We found that features with implicit importance scores had higher weights in the cold and heat categories than others, while they had lower weights in the tongue diagnosis category. In particular, the demographic information category and the nasal symptoms category showed low weights in the implicit importance score for diagnosis.

**Table 5.** Comparison between averaged explicit and implicit importance score of doctors by category

|  | #features | Explicit<br>Importance | Implicit (Dx<sup>1</sup>)<br>importance | Implicit (Tx<sup>2</sup>)<br>importance |
| --- | --- | --- | --- | --- |
| Demographic information | 8 | 14.22% | 8.29% ** | 12.28% |
| Nasal symptoms | 7 | 27.43% | 22.31% ** | 27.85% |
| Cold and heat | 3 | 15.03% | 28.19% * | 25.43% * |
| Digestion | 3 | 8.16% | 9.41% | 7.56% |
| Stools and urine | 5 | 9.49% | 10.93% | 9.66% |
| Other symptoms | 6 | 11.93% | 13.88% | 11.49% |
| Tongue diagnosis | 4 | 9.66% | 3.85% ** | 2.90% ** |
| Pulse diagnosis | 2 | 4.07% | 3.16% | 2.82% |

The average value of the feature in the radar chart was calculated and represented as a percentage. \*, Indicates categories with implicit importance scores more than 5% higher than the explicit importance score. \*\*, Indicates categories with implicit importance scores more than 5% lower than the explicit importance score. <sup>1</sup> diagnosis; <sup>2</sup> prescription (treatment).

### 3.3. Postquestionnaire study

Finally, to assess the usefulness of the tool, a postquestionnaire study was conducted with doctors who participated in the study. As shown in Figure 8 and Table 6, doctors provided positive responses regarding our proposed analytical tool overall. Doctors reported that the analysis results contained significant and valuable information (Question 6, Average score 4/5) and would be useful for the standardization of TAM (Question 11, Average score 3.75/5).

**Table 6.** Average score of the post-questionnaire question

| Questions | Average<br>score |
| --- | --- |
| 1. The analysis results are similar to those I expected. | 2.875 |
| 2. The questionnaire and diagnosis and prescription options used for the analysis were appropriate. | 3.375 |
| 3. I can interpret the terms and graphs used in the analysis well. | 3.5 |
| 4. I think that the results are biased. | 3.875 |
| 5.1. The results of the analysis could not represent my clinical practice well due to the limited PI diagnosis options. | 3.5 |
| 5.2. The results of the analysis could not represent my clinical practice well due to the limited herbal prescription options. | 3.5 |
| 5.3. The results of the analysis could not represent my clinical practice well due to not being able to add or remove herbs from the original prescription. | 3.5 |
| 5.4. The results of the analysis do not reflect my clinical practice well because the analysis was based on questionnaire, not based on face to face diagnosis. | 3.625 |
| 6. I think that these analysis results contain significant and valuable information for clinicians. | 4 |
| 7. The analysis results were worth the time and effort required to respond to the questionnaire. | 3.25 |
| 8. This analysis will be useful for the education of traditional Asian medical students. | 3.25 |
| 9. This analysis will be useful for training interns or residents. | 3.375 |
| 10. This analysis will be useful for my clinical practice. | 3.375 |
| 11. This analysis will be useful for the standardization of TAM<sup>1</sup>. | 3.75 |
| 12. I would like to recommend this analysis to other TAM doctors. | 3.375 |

<sup>1</sup> Traditional Asian medicine

**Figure 8.**
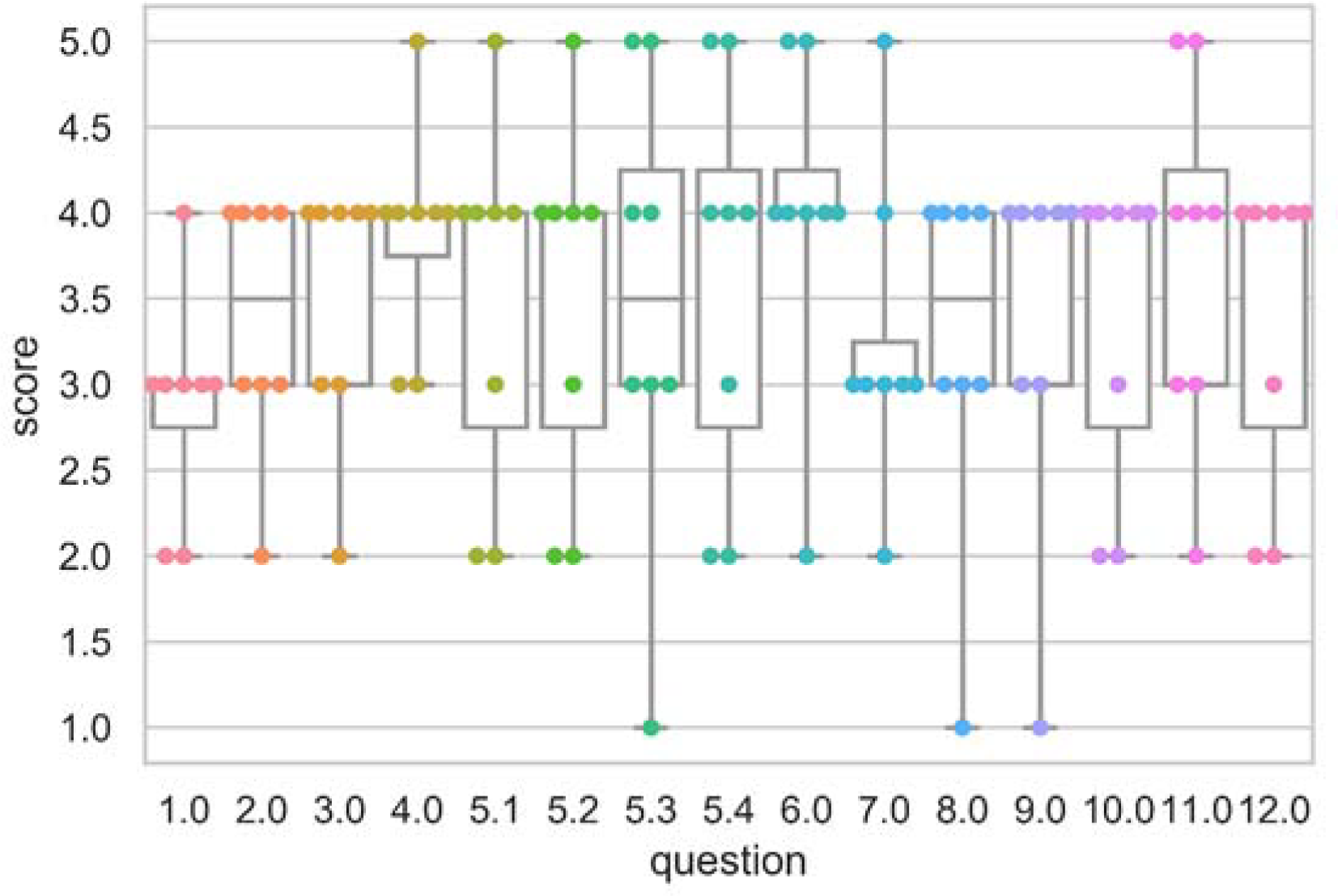
Scores on the postquestionnaire questions. The results of the postquestionnaire questions (multiple choice questions with a Likert scale) are presented as scatter plots and box plots. The questions can be found in Supplementary table 3.

## 4. Discussion

In this study, we operationally defined the explicit and implicit knowledge of PI based on the importance scores obtained based on TAM doctors’ scoring of the AR questionnaire and a supervised learning model that mimicked the doctors’ clinical decisions. The RF model successfully extracted doctors’ implicit knowledge, and using this information, we developed a tool to compare the implicit and explicit knowledge of doctors. We aimed to analyze doctors’ clinical decision-making processes using eight evaluation indicators. Finally, a postquestionnaire study was conducted to demonstrate the potential availability of the tool.

For the first time, we proposed a model to evaluate doctors’ clinical decision-making process and assess the degree of importance of each clinical feature. Several studies have employed machine learning techniques to analyze clinical data [26-29]; however, their research goal was the development of tools that mimic doctors’ clinical decisions. Here, our primary objective in employing a machine learning model was to extract doctors’ implicit knowledge. To minimize errors in labels, the supervised learning model selects each feature (clinical information) and assigns weights. Identifying the importance of features using supervised learning models can be a valuable approach for identifying the implicit clinical decision-making process through which doctors consciously or unconsciously derive their results.

We identified the advantages and potential for application of the tool through a postquestionnaire study. The doctors who participated in the pilot study reported that the results of our analysis tool provided significant and valuable information and that the tool shows potential for application to the standardization of TAM, clinical practice, education of medical students, and training of residents. They also suggested that if more diversified diagnosis and prescription options were provided in the questionnaire, the analysis results could better reflect real-world clinical practice.

Previous standardization studies of PI have classified patterns based on classical literature or the Delphi method. However, these approaches cannot derive knowledge of objective clinical decision-making processes because of their heavy reliance on subjective opinions. The proposed method will better standardize studies than conventional approaches because it derives results from clinical practice data in an objective manner. The participating doctors could compare their own decision-making processes (implicit and explicit) with those of other doctors. We expect that employing our tool could help solve the heterogeneity problem in the clinical decision-making process and be the key to standardizing and objectifying PI.

Different from guideline-based therapy, which offers the same therapy to most patients with the same disease, precision medicine (PM) is a new approach to patient care that takes into account individual variability [30]. PI in TAM is basically based on theme of holism and individualization [1]; therefore, it might provide useful ideas and tools for the development of PM, especially for chronic and complex diseases. However, despite being theoretically useful for PM, unstandardized PI is difficult to apply to PM because each doctor has different knowledge and makes different judgments. However, with the development of computational approaches such as machine learning, it is becoming possible to investigate the latent process of each doctor’s clinical decisions. The methodology and results of our study not only help to overcome the heterogeneity of PI by detecting implicit knowledge of clinicians but also contribute to the development of PM.

## 5. Conclusions

This study proposed a tool to evaluate and compare doctors’ decision-making processes in terms of their explicit and implicit knowledge. By comparing explicit and implicit importance scores, we identified the differences between doctors’ own explicit and implicit knowledge and the differences in these two types of knowledge among fellow doctors. The proposed tool to identify doctors’ decision-making processes is expected to be used in various ways in education, clinical and institutional contexts.

## Author Contributions

IC and CE designed the project. MHK, SYP and MK carried out the study. MP and CE developed the analytical methods. MHK and MP analyzed the data, interpreted the results and wrote the manuscript. IC and CE supervised the project. All authors have read and approved the final manuscript.

## Data Availability

All data produced in the present study are available upon reasonable request to the authors

## Acknowledgements

This work was supported by the Gachon University research fund of 2018 (GCU-2018-0296). This research was supported by the Korea Institute of Oriental Medicine (KSN2012110)

## Summary table

|  |
| --- |
| What was already known on the topic? |
| - Implicit knowledge is important for the doctor's clinical decision-making process.<br>- The pattern identification of traditional Asian medicine in clinical decision-making process is hard to objectify.<br>- Since machine learning is used for pattern recognition, it can be used to extract the implicit knowledge in the decision-making process of a doctor. |
| What this study added to our knowledge? |
| - We proposed an identification tool that explains and compare doctors' explicit and implicit knowledge.<br>- This tool can extract the doctor's implicit knowledge based a supervised machine learning technique, and compare the doctor's clinical making-decision process based on the extracted knowledge.<br>- Our tool will be helpful for the clinical standardization of traditional Asian medicine, doctors' own clinical practice, and intern/resident training. |

## Appendix A. Supplementary Materials

Material S1: The results of the analysis tool, Table S1. Pattern identification questionnaire of allergic rhinitis, Table S2. Features used in the evaluation of allergic rhinitis symptoms. Table S3. Postquestionnaire

