## Supplementary Table 1 for "Development of an explicit and implicit knowledge identification tool for the analysis of the decision-making process of traditional Asian medicine doctors"

Supplementary Table 1. Pattern identification questionnaire of allergic rhinitis

| **General characteristics** | | | | |
| --- | --- | --- | --- | --- |
| Age ( years) Gender ( M / F ) Body mass index ( kg/m^2^) | | | | |
| **Nasal symptoms** | | | | |
| These questions are about nasal symptoms. Please answer all of the questions 1-6 by marking the answer that most closely describes your symptoms over the last 2 days (0=None, 1=Mild, 2=Moderate, or 3=Severe). Clinician checks the score of question 7-8 after enquiring about the color and viscosity of rhinorrhea | | | | |
| **Subjective nasal symptoms** | 0 | 1 | 2 | 3 |
| 1. Rhinorrhea |  |  |  |  |
| 2. Nasal congestion |  |  |  |  |
| 3. Nasal itching |  |  |  |  |
| 4. Sneezing |  |  |  |  |
| 5. Nasal dryness |  |  |  |  |
| 6. The color and viscosity of rhinorrhea | □ 0. Totally watery and transparent  □ 1. Slightly watery and transparent  □ 2. Slightly sticky and yellowish  □ 3. Totally sticky and yellowish | | | |
| **Objective nasal symptoms** | | | | |
| 7. The nasal membrane color | □ 0. Severe pale  □ 1. Mild pale  □ 2. Normal  □ 3. Mild hyperemia  □ 4. Severe hyperemia | | | |
| 8. Inferior turbinate hypertrophy | □ 0. Normal  □ 1. Mild  □ 2. Severe | | | |
| **General conditions** | | | | |
| Please answer all of the questions by marking the answer that most closely describes your symptoms over the last 2 weeks (0=None, 1=Mild, 2=Moderate, or 3=Severe). | | | | |
| **Cold and heat** | 0 | 1 | 2 | 3 |
| 9. Aversion to cold |  |  |  |  |
| 10. Aversion to heat |  |  |  |  |
| 11. Heat in the upper body |  |  |  |  |
| 12. Preference for cold water |  |  |  |  |
| 13. Preference for hot water |  |  |  |  |
| 14. Dry mouth and thirst |  |  |  |  |
| 15. Dry eye |  |  |  |  |
| **Digestion (0=None, 1=Mild, 2=Moderate, or 3=Severe)** | | | | |
| 16. I have a feeling of fullness in my stomach after eating |  |  |  |  |
| 17. I feel abdominal gas |  |  |  |  |
| 18. I have a poor appetite and eat just a little food |  |  |  |  |
| **Stools** |  |  |  |  |
| 19. Soft stool | □ 0. Rarely  □ 1. Sometimes  □ 2. Often  □ 3. Always | | | |
| 20. Dry stool | □ 0. Rarely  □ 1. Sometimes  □ 2. Often  □ 3. Always | | | |
| 21. Stool frequency | □ 0. One or 2 times per week  □ 1. Once every two days  □ 2. Daily  □ 3. >2 times per day | | | |
| **Urine** |  |  |  |  |
| 22. Transparent with no yellow color |  |  |  |  |
| 23. Yellow color |  |  |  |  |
| **General** |  |  |  |  |
| 24. I usually feel tired or languid |  |  |  |  |
| **Facial complexion** |  |  |  |  |
| 25. Pale white complexion |  |  |  |  |
| 26. Yellow complexion |  |  |  |  |
| 27. Reddened complexion |  |  |  |  |
| **Sweat** |  |  |  |  |
| 28. Excess sweating |  |  |  |  |
| **Sleep** |  |  |  |  |
| 29. Sleep quality | □ 0. Very good  □ 1. Good  □ 2. Fair  □ 3. Poor | | | |
| **Tongue diagnosis** |  |  |  |  |
| 30. Tongue color | □ Pale □ Red □ Purple □ Red in tip of the tongue | | | |
| 31. Tongue body | □ Very dry □ Little dry □ Little moist □ Very moist | | | |
| 32A. Tongue fur (coating)  32B. Tongue fur (color) | □ Thin □ Thick □ Slimy  □ White □ Moderate □ Yellow | | | |
| **Pulse diagnosis** |  |  |  |  |
| 33. Week/forceful | □ Very weak □ Weak □ Forceful □ Very forceful | | | |
| 34. Slow/rapid | Per minute | | | |
