## Supplementary Table 2 for "Development of an explicit and implicit knowledge identification tool for the analysis of the decision-making process of traditional Asian medicine doctors"

Supplementary Table 2. Features used in the evaluation of allergic rhinitis symptoms

| 1 | Rhinorrhea | 14 | Dry mouth and thirst | 27 | Reddened complexion |
| --- | --- | --- | --- | --- | --- |
| 2 | Nasal congestion | 15 | Dry eye | 28 | Excess sweating |
| 3 | Nasal itching | 16 | Feeling of fullness in my stomach after eating | 29 | Sleep quality |
| 4 | Sneezing | 17 | abdominal gas | 30 | Tongue color |
| 5 | Nasal dryness | 18 | poor appetite and eat just a little food | 31 | Tongue body |
| 6 | Color and viscosity of rhinorrhea | 19 | Soft stool | 32A | Tongue fur (coating) |
| 7 | The nasal membrane color | 20 | Dry stool | 32B | Tongue fur (color) |
| 8 | Inferior turbinate hypertrophy | 21 | Stool frequency | 33 | Pulse week/forceful |
| 9 | Aversion to cold | 22 | Transparent with no yellow color | 34 | Pulse slow/rapid |
| 10 | Aversion to heat | 23 | Yellow color | SPT | Score of skin prick test |
| 11 | Heat in the upper body | 24 | feel tired or languid | Age | Age |
| 12 | Preference for cold water | 25 | Pale white complexion | Gender | Gender |
| 13 | Preference for hot water | 26 | Yellow complexion | BMI | Body Mass Index (kg/$m^{2})$ |
