## Supplementary Table 3 for "Development of an explicit and implicit knowledge identification tool for the analysis of the decision-making process of traditional Asian medicine doctors"

Supplementary Table 3. Post-questionnaire

| Thank you for participating in this study developing an assessment tool for definition of the clinical decision-making process of traditional Asian medicine (TAM) clinicians.  Based on your response in questionnaire, we will provide ‘explicit and implicit Importance’ and the data of comparative analysis with other TAM doctors for diagnosis and treatment of their allergic rhinitis. Please read each one carefully, check one each line. | | | | | | |
| --- | --- | --- | --- | --- | --- | --- |
|  | | Strongly disagree | Tend to disagree | Neither agree nor disagree | Tend to agree | Strongly agree |
|  |  | 1 | 2 | 3 | 4 | 5 |
| 1. The analysis results are similar to those I expected. | |  |  |  |  |  |
| 2. The questionnaire* and diagnosis and prescription options used for the analysis were appropriate.  * scoring the importance of each symptoms in the clinical dataset filled out by patients and choosing the pattern identification (PI) and herbal prescription for each patient | |  |  |  |  |  |
|  | 2.1 If not, what was particularly inappropriate? |  | | | | |
| 3. I can interpret the terms and graphs used in the analysis well. | |  |  |  |  |  |
|  | 3.1 If not, what was not particularly well understood? |  | | | | |
| 4. I think that the results are biased. | |  |  |  |  |  |
| 5. The following questions are about potential source of bias if you think clinical dataset-based analysis bias the results. | | | | | | |
|  | 5.1 The results of the analysis could not represent my clinical practice well due to the limited PI diagnosis options*.  * lung-cold, lung-heat or spleen qi deficiency |  |  |  |  |  |
|  | 5.2 The results of the analysis could not represent my clinical practice well due to the limited herbal prescription options*.  * Galgeun-tang, Socheongryong-Tang, Hyeonggaeyeongyo-tang, Yeotaektonggi-tang, Bojungikgi-tang |  |  |  |  |  |
|  | 5.3 The results of the analysis could not represent my clinical practice well due to not being able to add or remove herbs from the original prescription. |  |  |  |  |  |
|  | 5.4 The results of the analysis do not reflect my clinical practice well because the analysis was based on questionnaire, not based on face to face diagnosis. |  |  |  |  |  |
|  | 5.5 What else do you think is the reason why your explicit and implicit importance is different? |  | | | | |
| 6. I think that these analysis results contain significant and valuable information for clinicians. | |  |  |  |  |  |
|  | 6.1 If you think analysis results are valuable, which graphs are thought to contain particularly significant information? |  | | | | |
| 7. The analysis results were worth the time and effort required to respond to the questionnaire*.  * scoring the importance of each symptoms in the clinical dataset filled out by patients and choosing the pattern identification (PI) and herbal prescription for each patient | |  |  |  |  |  |
| 8. This analysis will be useful for the education of traditional Asian medical students.  (Even if you are not educating someone, answer from the position in charge of education) | |  |  |  |  |  |
| 9. This analysis will be useful for training interns or residents.  (Even if you are not educating someone, answer from the position in charge of education) | |  |  |  |  |  |
| 10. This analysis will be useful for my clinical practice. | |  |  |  |  |  |
| 11. This analysis will be useful for the standardization of TAM. | |  |  |  |  |  |
| 12. I would like to recommend this analysis to other TAM doctors. | |  |  |  |  |  |
| * Please write down any comments or suggestions for this study. | | | | | | |
