## Supplementary material 1 for "Development of an explicit and implicit knowledge identification tool for the analysis of the decision-making process of traditional Asian medicine doctors"

### Allergic Rhinitis Diagnostic Analysis Assessment Tool

- This sheet evaluates the knowledge used in the clinical decision process using a tool developed to objectify.
- The doctor who participated in the assessment can check the hidden knowledge at the process of diagnosis or prescription and compare with the average of doctors who participated in the assessment.
- Note that since this paper has been developed to help doctor's clinical decision-making, there is no correct answer for this assessment.

❖ The features used for analysis are as follows:

| SPT | Score of skin prick test | | Age | | Age |
|---|---|---|---|---|---|
| Gender | Gender | | BMI | | BMI (kg/$m^2$) |
| 1 | Rhinorrhea | 13 | Preference for hot water | 25 | Pale white complexion |
| 2 | Nasal congestion | 14 | Dry mouth and thirst | 26 | Yellow complexion |
| 3 | Nasal itching | 15 | Dry eye | 27 | Reddened complexion |
| 4 | Sneezing | 16 | Feeling of fullness in my stomach after eating | 28 | Excess sweating |
| 5 | Nasal dryness | 17 | abdominal gas | 29 | Sleep quality |
| 6 | Color and viscosity of rhinorrhea | 18 | poor appetite and eat just a little food | 30 | Tongue color |
| 7 | The nasal membrane color | 19 | Soft stool | 31 | Tongue body |
| 8 | Inferior turbinate hypertrophy | 20 | Dry stool | 32A | Tongue fur (coating) |
| 9 | Aversion to cold | 21 | Stool frequency | 32B | Tongue fur (color) |
| 10 | Aversion to heat | 22 | Transparent with no yellow color | 33 | Pulse week/forceful |
| 11 | Heat in the upper body | 23 | Yellow color | 34 | Pulse slow/rapid |
| 12 | Preference for cold water | 24 | feel tired or languid | | |

### Doctor #1

#### Glossary & Tips

1. Explicit knowledge & implicit knowledge
- Knowledge can be classified into two different categories: explicit and implicit knowledge. Explicit knowledge refers to knowledge that can be expressed in words and in a form that can be codified. In contrast, implicit knowledge refers to knowledge that an individual obtains through experience but does not convey to others.
- In this analysis, the explicit importance score was defined for the features obtained from doctors' label indicating the degree of importance of each symptom for diagnosing and treating AR patients. Implicit importance was defined based on the weights of the features learned by the machine learning algorithm when the learning algorithm effectively reproduced the doctor's decision-making process.

2. Baseline
- The baseline is the average value of the implicit importance score of individual results.

3. The explicit importance score, implicit importance score for diagnosis, and implicit importance score for prescription are visualized in orange, cyan, and blue, respectively. Individual results from each doctor and the average of all doctors' results are expressed as a bar graph and a dotted line, respectively

4. The statistic analysis was calculated by Spearman's rank correlation coefficient (p-value < 0.05).

(M1) Comparison of the individual and average explicit and implicit importance scores

To provide an overview of the differences in all importance scores, M1 was visualized with line graphs that consisted of all six scores. Scores were sorted in descending order according to the average explicit importance score of average results

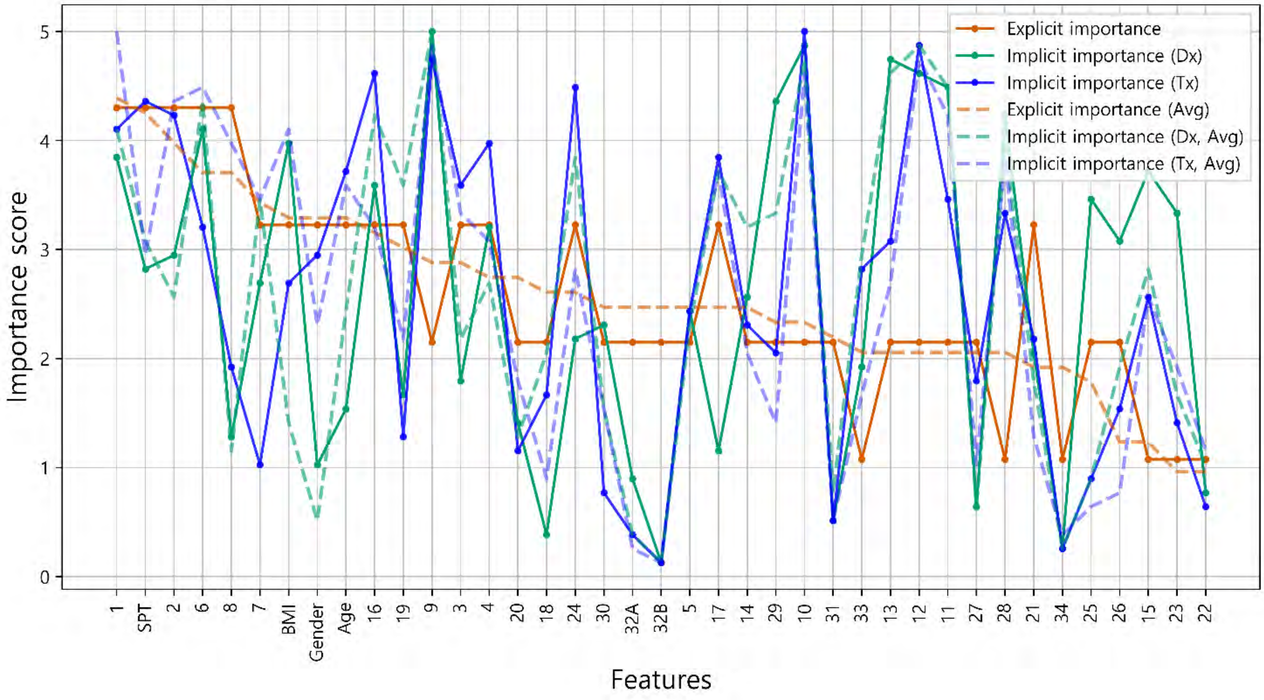


(M2) Intra-individual correlation of explicit and implicit importance

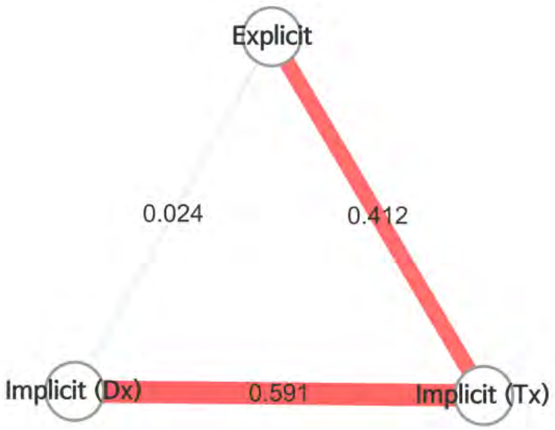


- To examine the overall correlation between the explicit and implicit knowledge of individuals, M2 was presented as nodes and edges, which represented the types of importance scores and the correlations between importance scores, respectively.
- Color of edge : the results of the Bonferroni post hoc test
  - Dark red: statistically significant
  - light red: not significant, but p-value<0.05
  - grey : p-value>0.05
- Thickness of edge (statistic): the thicker the edge, the greater the correlation.

- It is analyzed that there is no correlation between the explicit importance and implicit diagnostic importance (ρ = 0.024, p-value = 0.883). Explicit importance and implicit prescription importance have been identified as having a weak correlation (ρ = 0.412, p-value = 0.009).

**Doctor #1** (M3) Radar chart analysis

All the features used in the analysis were classified into eight categories and visualized as a radar chart to provide the information about the feature categories that the doctor referred to for treatment. The categories consisted of demographic features of the patient, nose symptoms, cold-hot, digestion, feces and urine, tongue diagnosis, pulse diagnosis and other symptoms

- The explicit importance score, implicit importance score for diagnosis, and implicit importance score for prescription are visualized in orange, cyan, and blue, respectively.
- Individual results from each doctor and the average of all doctors' results are represented as a solid line and a dotted line, respectively.

Comparison of Doctor 1 and average explicit importance scores

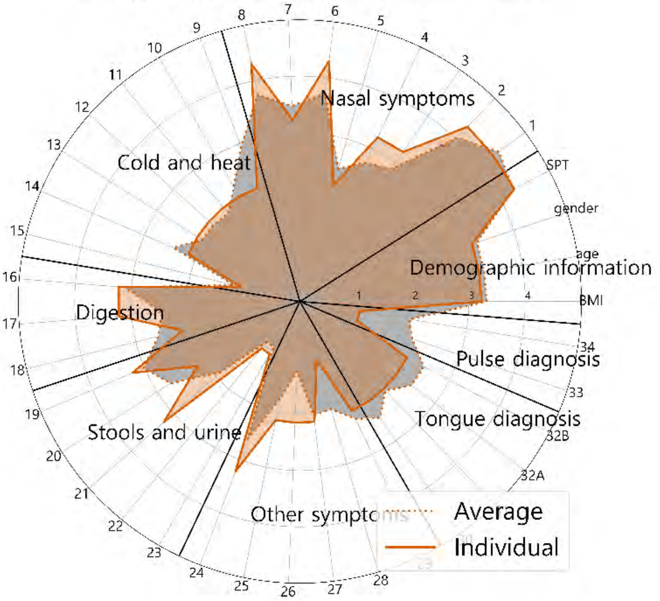


Comparison of Doctor 1 and average implicit importance scores for diagnosis

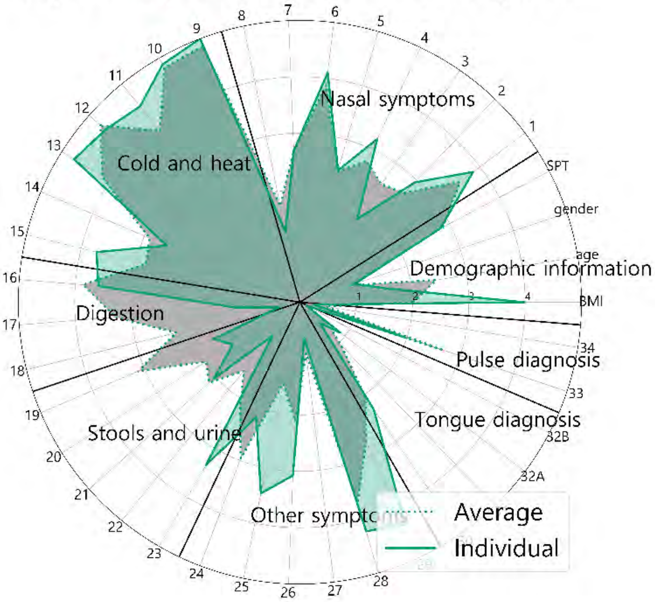


Comparison of Doctor 1 and average implicit importance scores for prescription

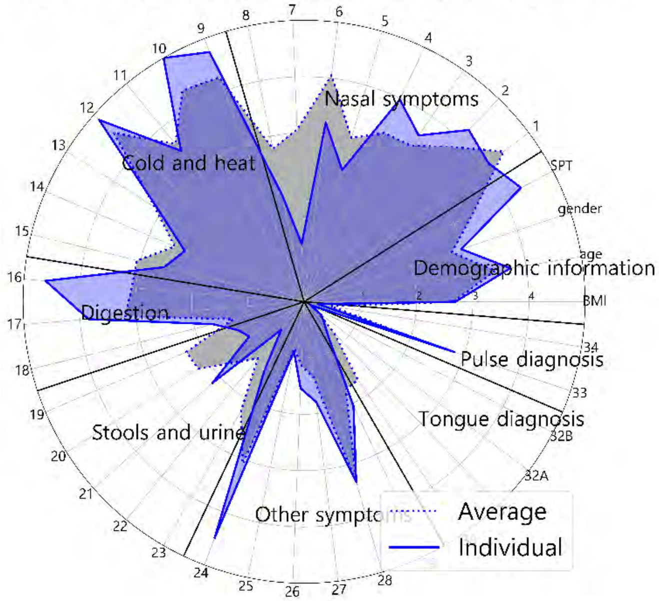


Comparison of explicit and implicit importance scores

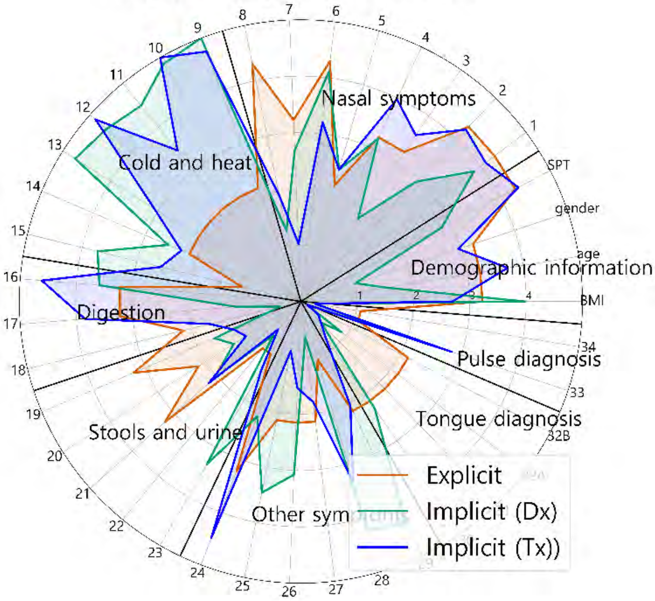


Comparison of explicit and implicit importance scores (average results)

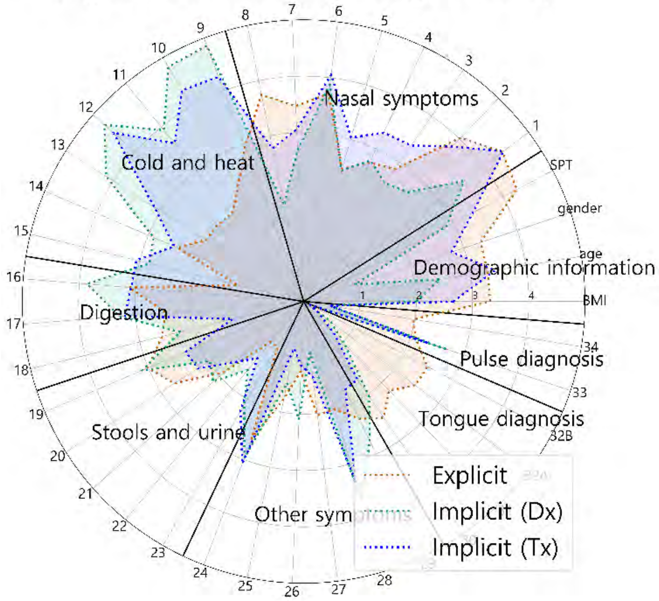

### Doctor #1

### Supplementary sheet

(S1) Intra-individual comparison of explicit importance score and implicit importance score for diagnosis

(S2) Intra-individual comparison of explicit importance score and implicit importance score for prescription

S1 and S2 were represented as scatter plots. The correlation statistics between the explicit and implicit scores were calculated using Spearman's rank correlation coefficient test. The X-axis represents the normalized value of the implicit importance and the Y-axis represents the normalized value of the explicit importance.

| Result of Spearman test | statistic | 0.024 |
|---|---|---|
| | p-value | 0.883 |

| Result of Spearman test | statistic | 0.412 |
|---|---|---|
| | p-value | 0.009 |

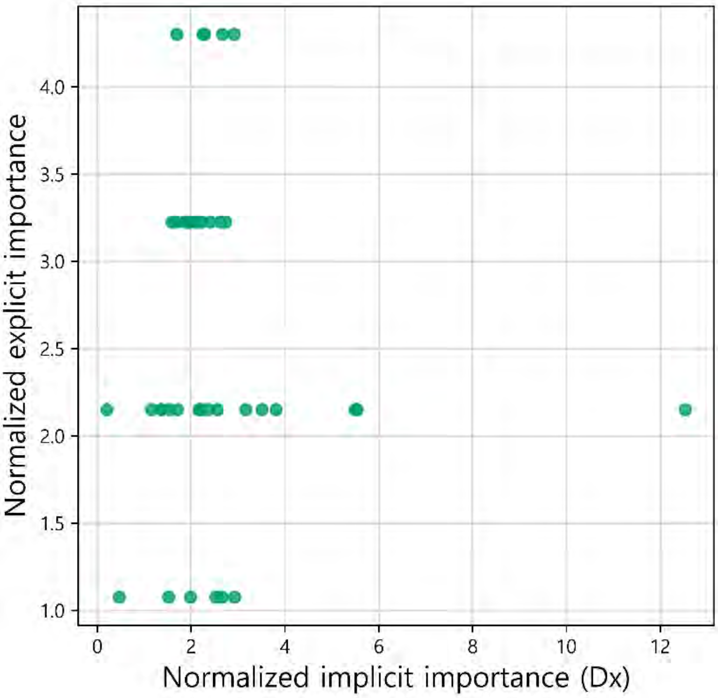


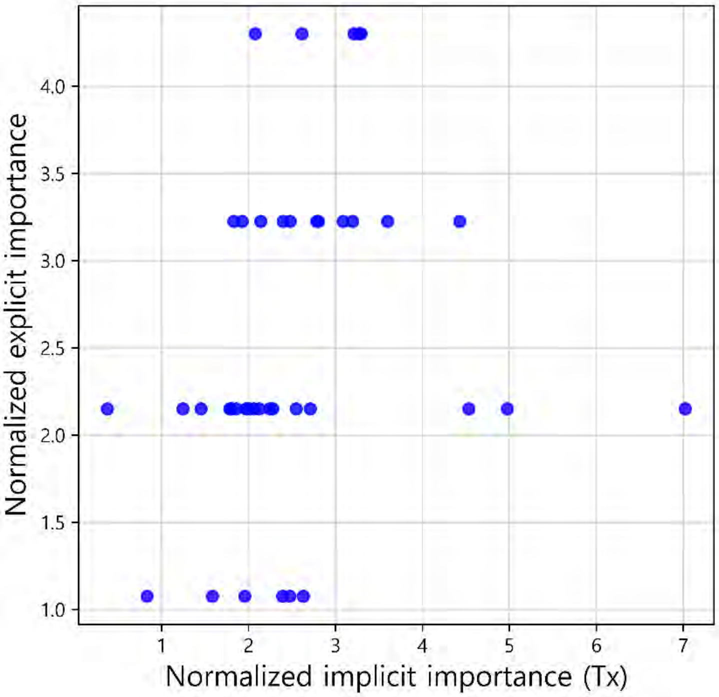


(S3) Comparison of the individual and average explicit importance scores

S3 was constructed to compare the average importance score of the doctors as a group with the individual doctor's importance score. Individual results from each doctor and the average of all doctors' results are expressed as a bar graph and a dotted line, respectively. The graphs are sorted in descending order according to the importance score of the average results.

| Result of Spearman test | |
|---|---|
| statistic | 0.832 |
| p-value | < 0.001 |

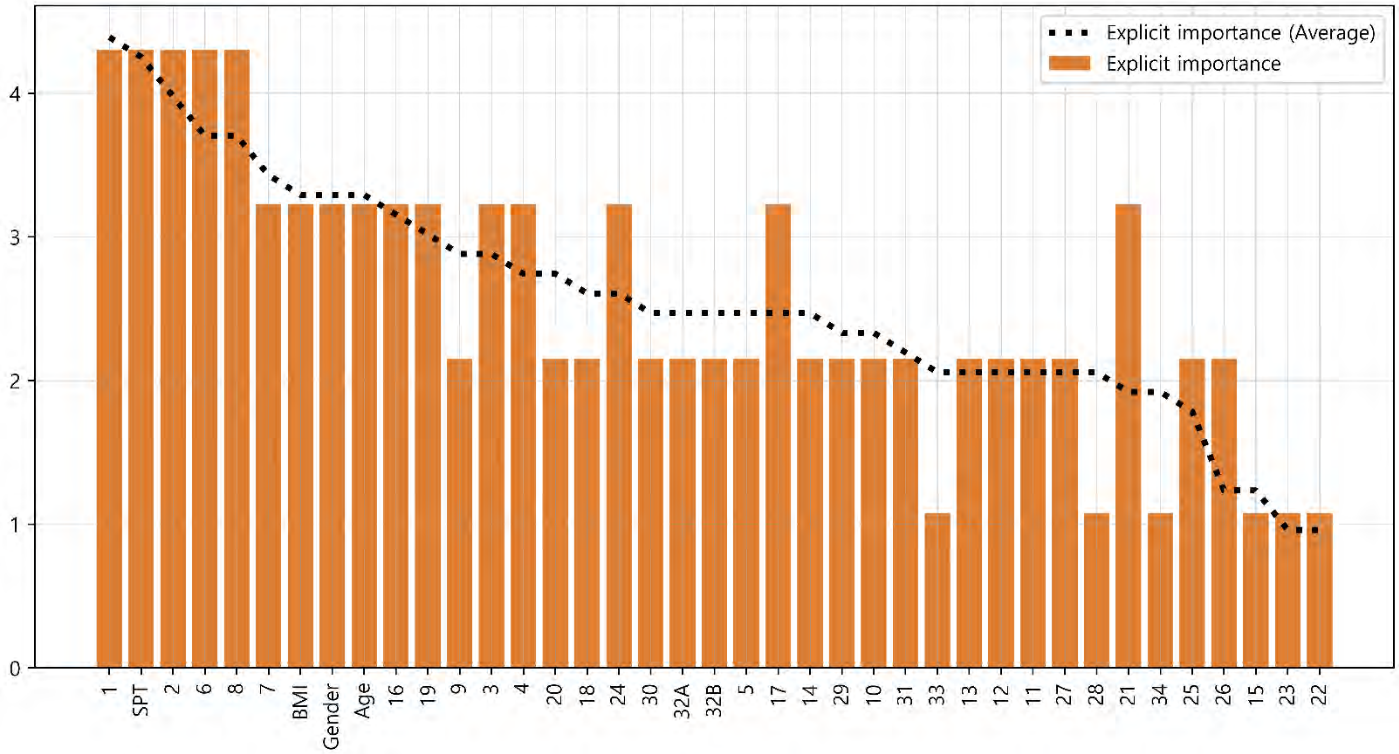

#### (S4) Comparison of the individual and average implicit importance scores for diagnosis

S4 was constructed to compare the average importance score of the doctors as a group with the individual doctor's importance score. Individual results from each doctor and the average of all doctors' results are expressed as a bar graph and a dotted line, respectively. The graphs are sorted in descending order according to the importance score of the average results.

| Result of Spearman test | |
|---|---|
| statistic | 0.751 |
| p-value | < 0.001 |

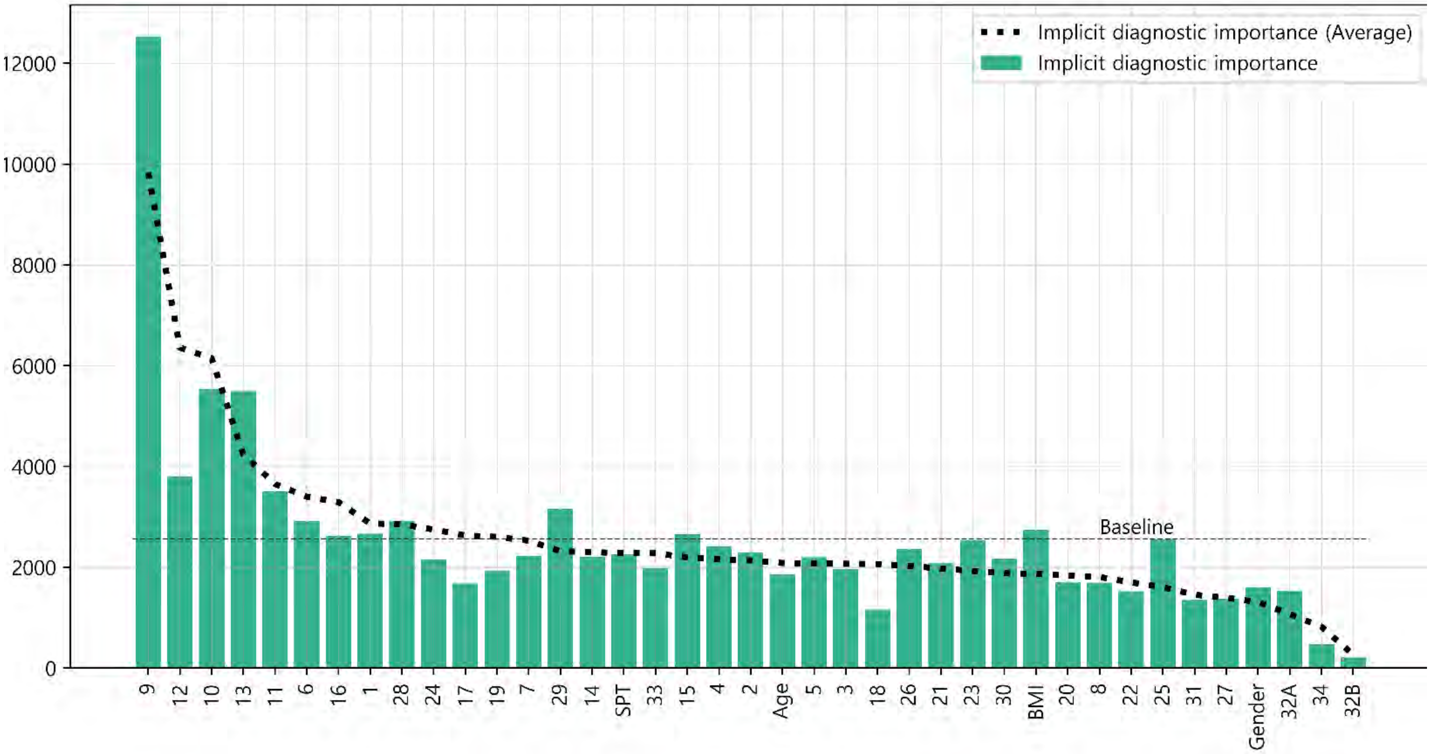


#### (S5) Comparison of the individual and average implicit importance scores for prescription

S5 was constructed to compare the average importance score of the doctors as a group with the individual doctor's importance score. Individual results from each doctor and the average of all doctors' results are expressed as a bar graph and a dotted line, respectively. The graphs are sorted in descending order according to the importance score of the average results.

| Result of Spearman test | |
|---|---|
| statistic | 0.811 |
| p-value | < 0.001 |

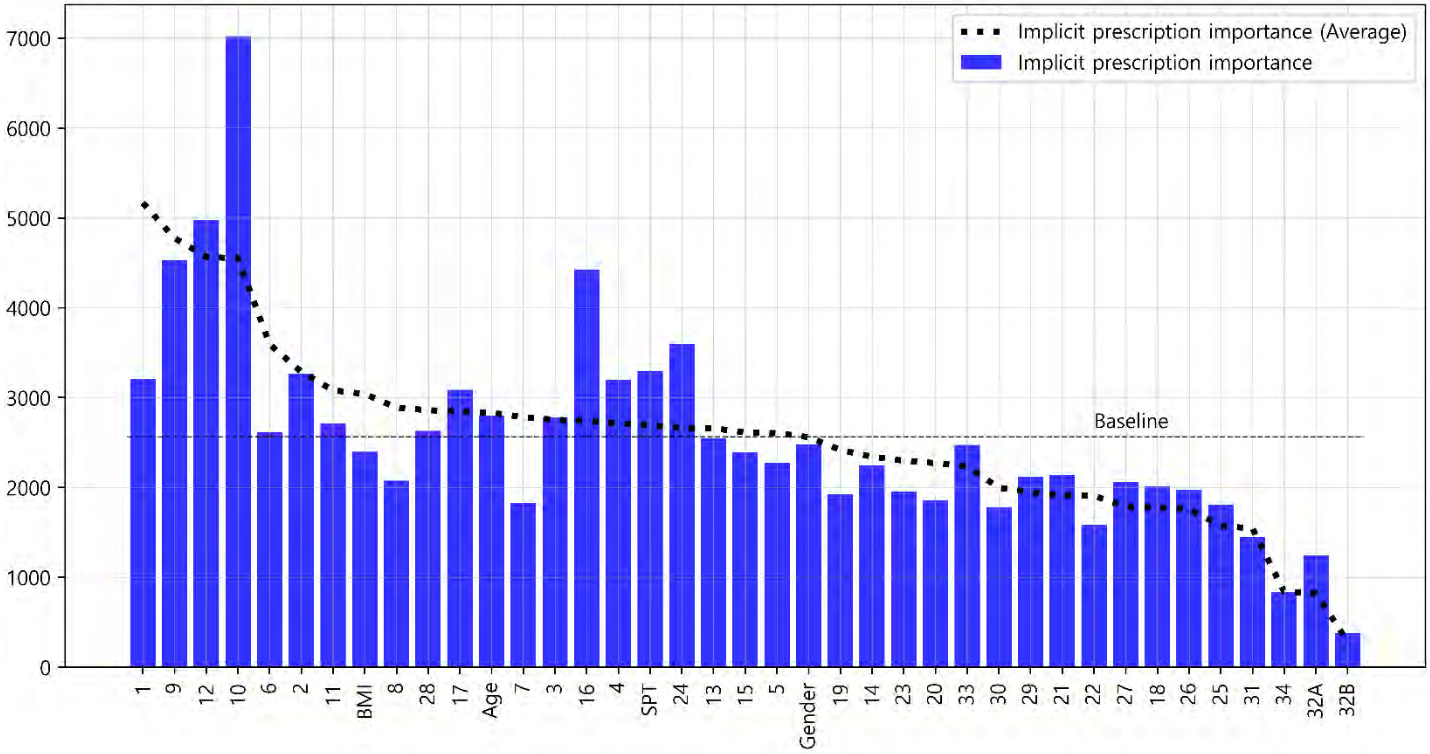

### Doctor #2

#### Glossary & Tips

1. Explicit knowledge & implicit knowledge

- Knowledge can be classified into two different categories: explicit and implicit knowledge. Explicit knowledge refers to knowledge that can be expressed in words and in a form that can be codified. In contrast, implicit knowledge refers to knowledge that an individual obtains through experience but does not convey to others.
- In this analysis, the explicit importance score was defined for the features obtained from doctors' label indicating the degree of importance of each symptom for diagnosing and treating AR patients. Implicit importance was defined based on the weights of the features learned by the machine learning algorithm when the learning algorithm effectively reproduced the doctor's decision-making process.

2. Baseline

- The baseline is the average value of the implicit importance score of individual results.

3. The explicit importance score, implicit importance score for diagnosis, and implicit importance score for prescription are visualized in orange, cyan, and blue, respectively. Individual results from each doctor and the average of all doctors' results are expressed as a bar graph and a dotted line, respectively

4. The statistic analysis was calculated by Spearman's rank correlation coefficient (p-value < 0.05).

(M1) Comparison of the individual and average explicit and implicit importance scores

To provide an overview of the differences in all importance scores, M1 was visualized with line graphs that consisted of all six scores. Scores were sorted in descending order according to the average explicit importance score of average results

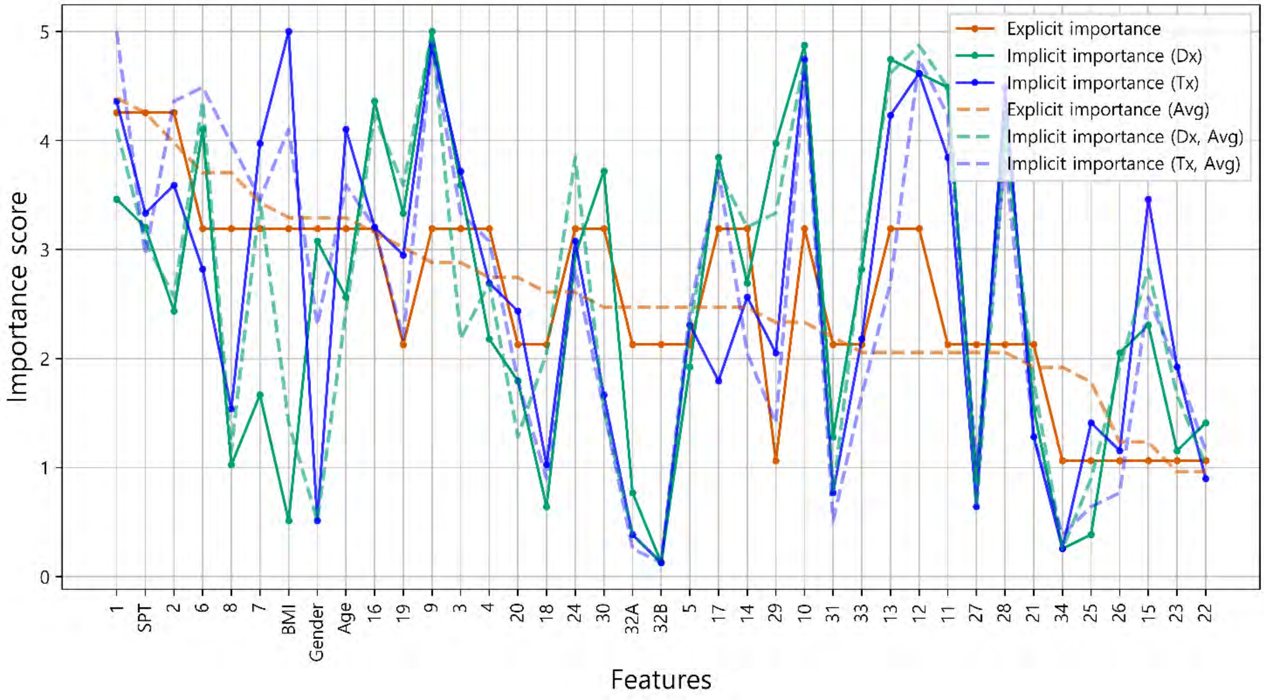


(M2) Intra-individual correlation of explicit and implicit importance

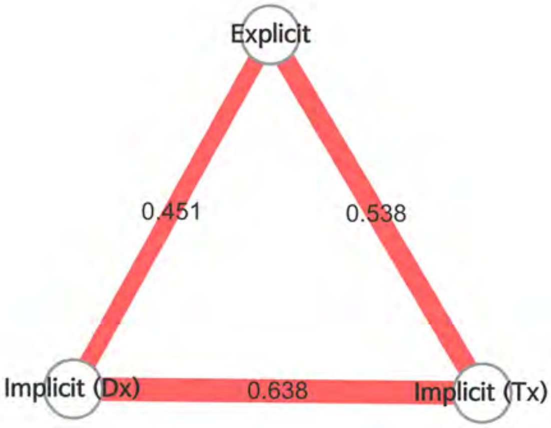


- To examine the overall correlation between the explicit and implicit knowledge of individuals, M2 was presented as nodes and edges, which represented the types of importance scores and the correlations between importance scores, respectively.
- Color of edge : the results of the Bonferroni post hoc test
  - Dark red: statistically significant
  - light red: not significant, but p-value<0.05
  - grey : p-value>0.05
- Thickness of edge (statistic): the thicker the edge, the greater the correlation.

- It is analyzed that there is a weak correlation between the explicit importance and implicit diagnostic importance (ρ = 0.451, p-value = 0.004). Explicit importance and implicit prescription importance have been identified as having a weak correlation (ρ = 0.538, p-value < 0.001).

**Doctor #2** (M3) Radar chart analysis

All the features used in the analysis were classified into eight categories and visualized as a radar chart to provide the information about the feature categories that the doctor referred to for treatment. The categories consisted of demographic features of the patient, nose symptoms, cold-hot, digestion, feces and urine, tongue diagnosis, pulse diagnosis and other symptoms

- The explicit importance score, implicit importance score for diagnosis, and implicit importance score for prescription are visualized in orange, cyan, and blue, respectively.
- Individual results from each doctor and the average of all doctors' results are represented as a solid line and a dotted line, respectively.

Comparison of Doctor 2 and average explicit importance scores

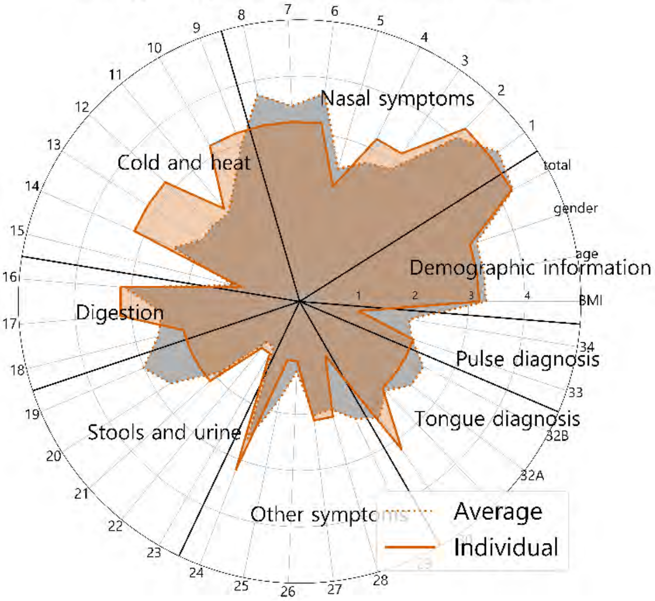


Comparison of Doctor 2 and average implicit importance scores for diagnosis

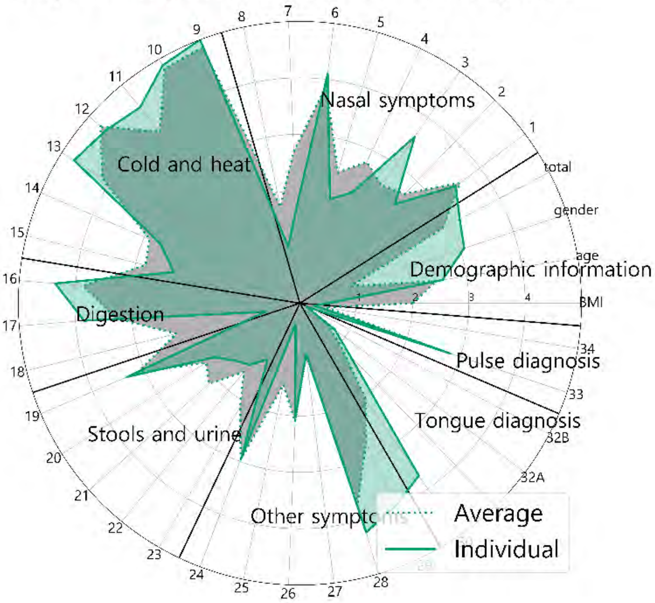


Comparison of Doctor 2 and average implicit importance scores for prescription

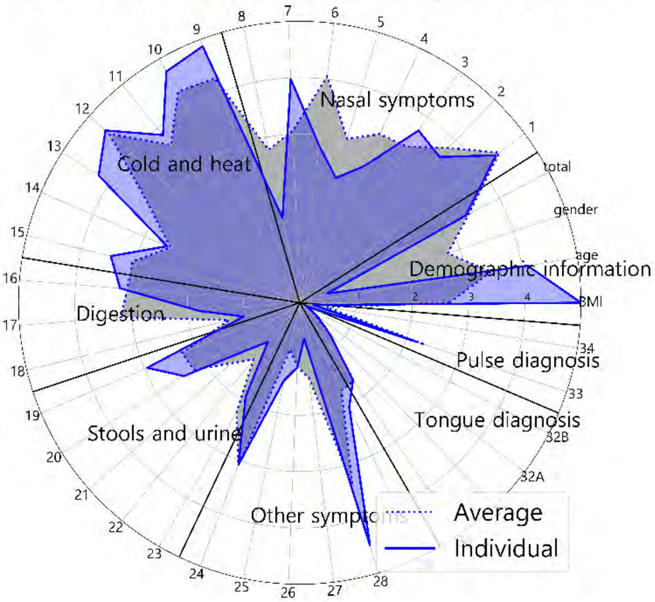


Comparison of explicit and implicit importance scores

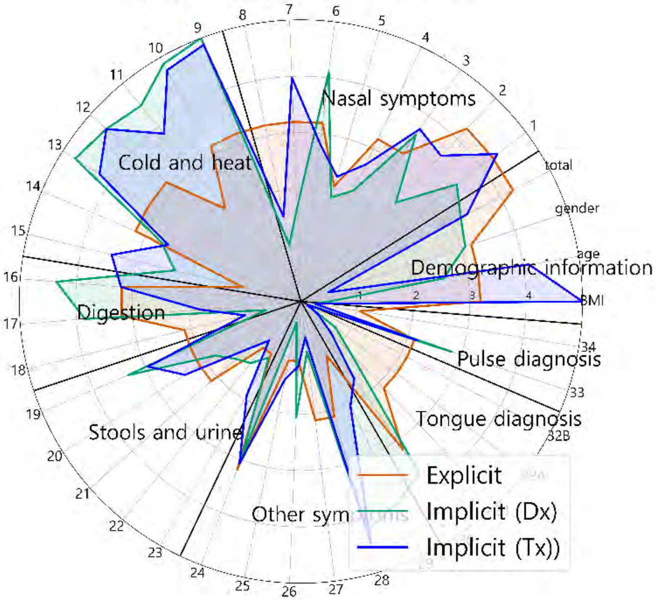


Comparison of explicit and implicit importance scores (average results)

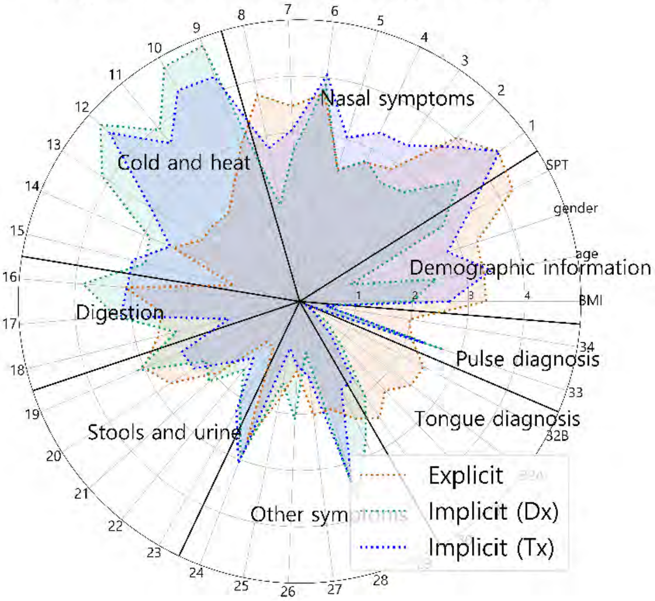

### Doctor #2

### Supplementary sheet

(S1) Intra-individual comparison of explicit importance score and implicit importance score for diagnosis

(S2) Intra-individual comparison of explicit importance score and implicit importance score for prescription

S1 and S2 were represented as scatter plots. The correlation statistics between the explicit and implicit scores were calculated using Spearman's rank correlation coefficient test. The X-axis represents the normalized value of the implicit importance and the Y-axis represents the normalized value of the explicit importance.

| Result of Spearman test | statistic | 0.451 |
|---|---|---|
| | p-value | 0.004 |

| Result of Spearman test | statistic | 0.538 |
|---|---|---|
| | p-value | < 0.001 |

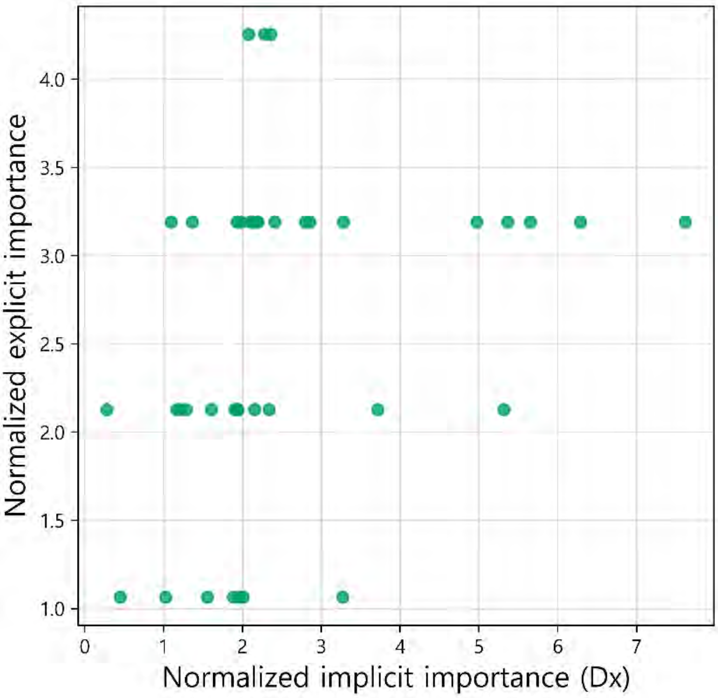


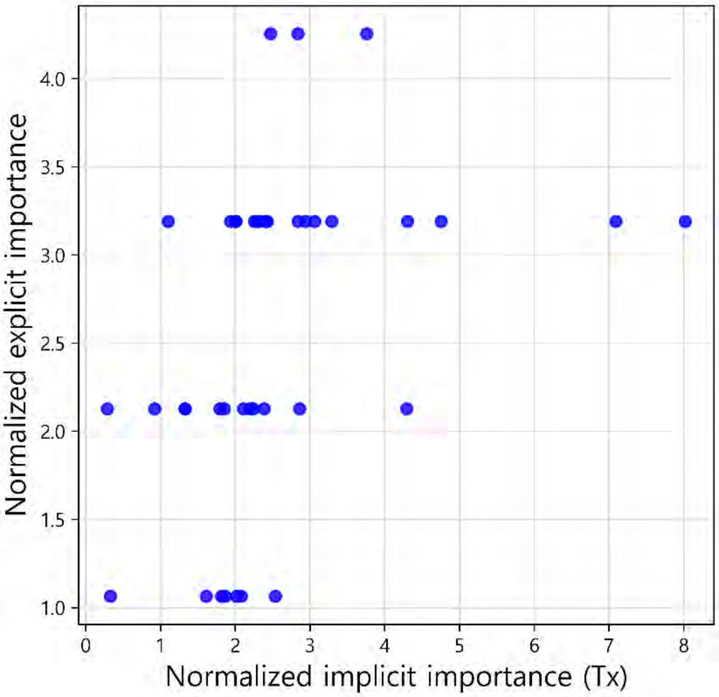


(S3) Comparison of the individual and average explicit importance scores

S3 was constructed to compare the average importance score of the doctors as a group with the individual doctor's importance score. Individual results from each doctor and the average of all doctors' results are expressed as a bar graph and a dotted line, respectively. The graphs are sorted in descending order according to the importance score of the average results.

| Result of Spearman test | |
|---|---|
| statistic | 0.780 |
| p-value | < 0.001 |

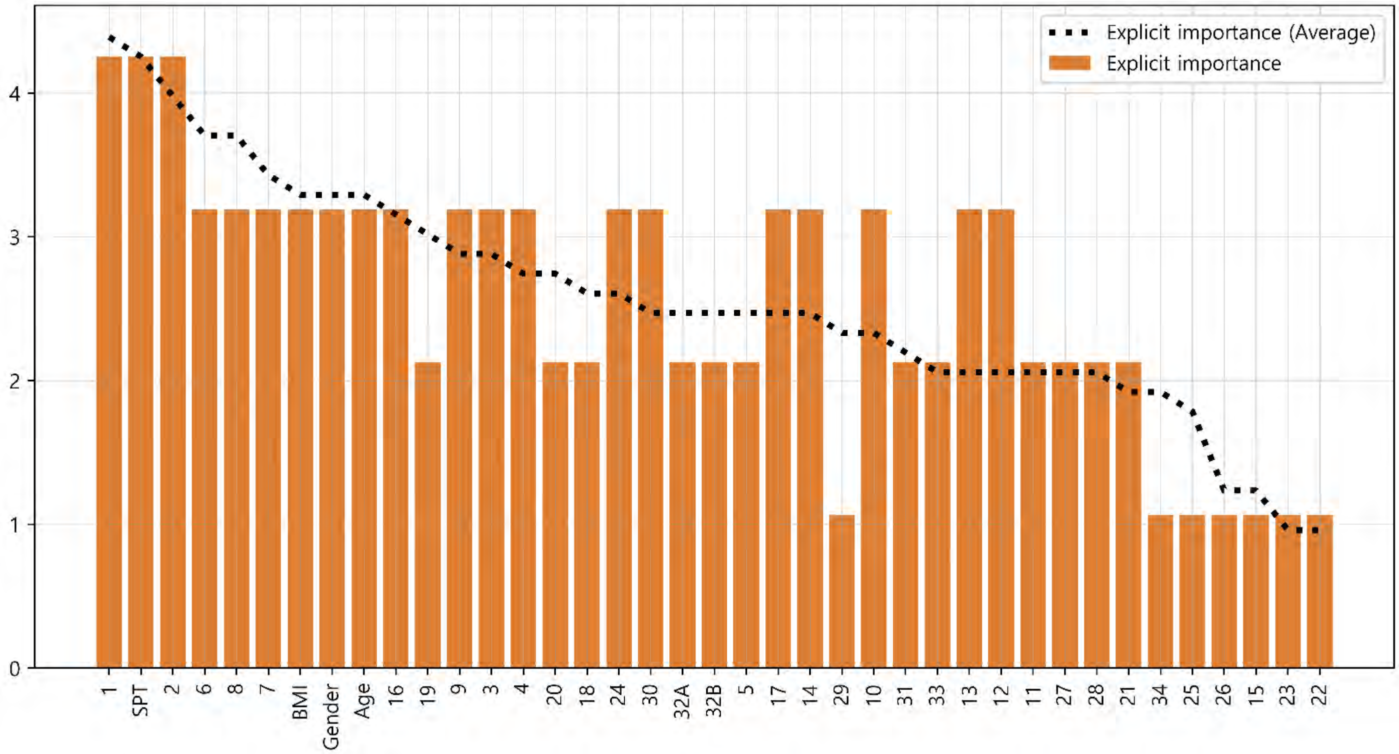

### Doctor #2

### Supplementary sheet

(S4) Comparison of the individual and average implicit importance scores for diagnosis

S4 was constructed to compare the average importance score of the doctors as a group with the individual doctor's importance score. Individual results from each doctor and the average of all doctors' results are expressed as a bar graph and a dotted line, respectively. The graphs are sorted in descending order according to the importance score of the average results.

| Result of Spearman test | |
|---|---|
| statistic | 0.853 |
| p-value | < 0.001 |

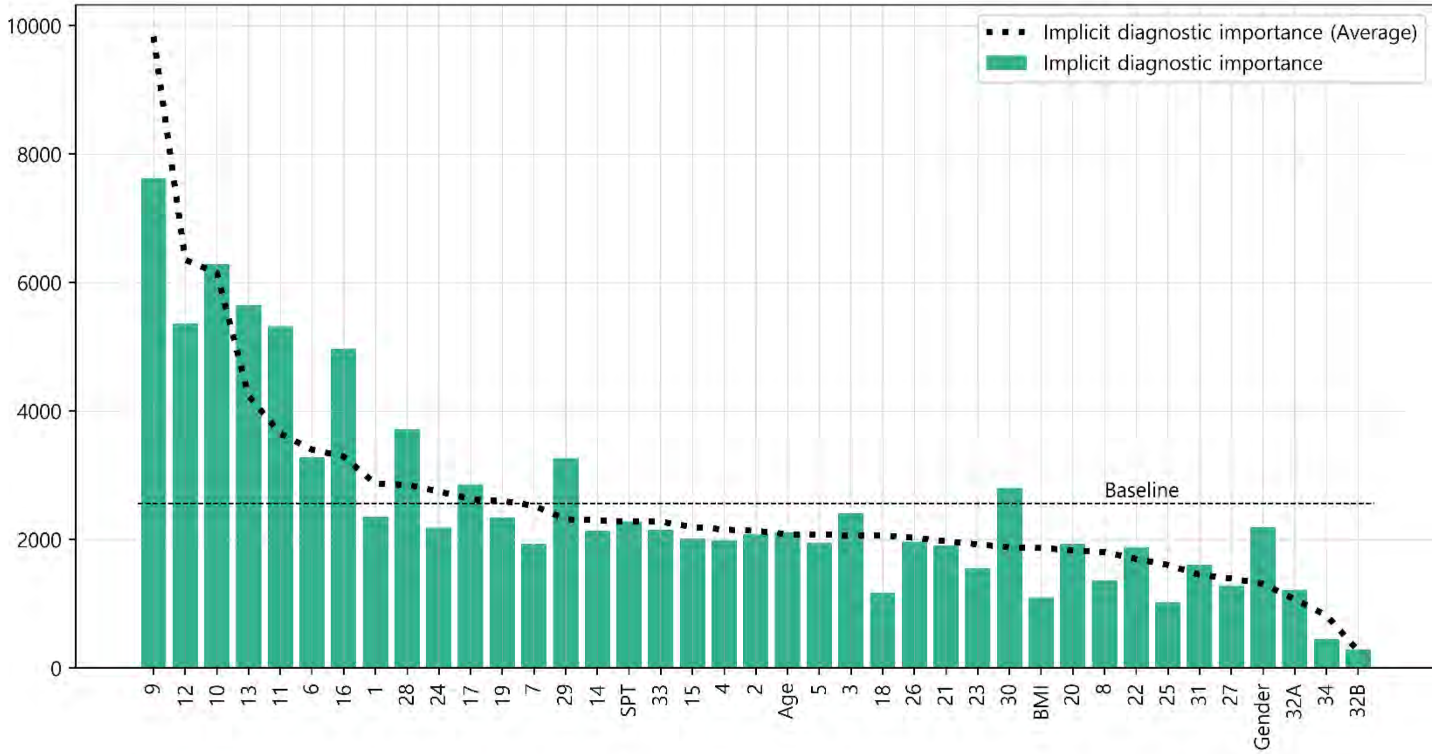


(S5) Comparison of the individual and average implicit importance scores for prescription

S5 was constructed to compare the average importance score of the doctors as a group with the individual doctor's importance score. Individual results from each doctor and the average of all doctors' results are expressed as a bar graph and a dotted line, respectively. The graphs are sorted in descending order according to the importance score of the average results.

| Result of Spearman test | |
|---|---|
| statistic | 0.844 |
| p-value | < 0.001 |

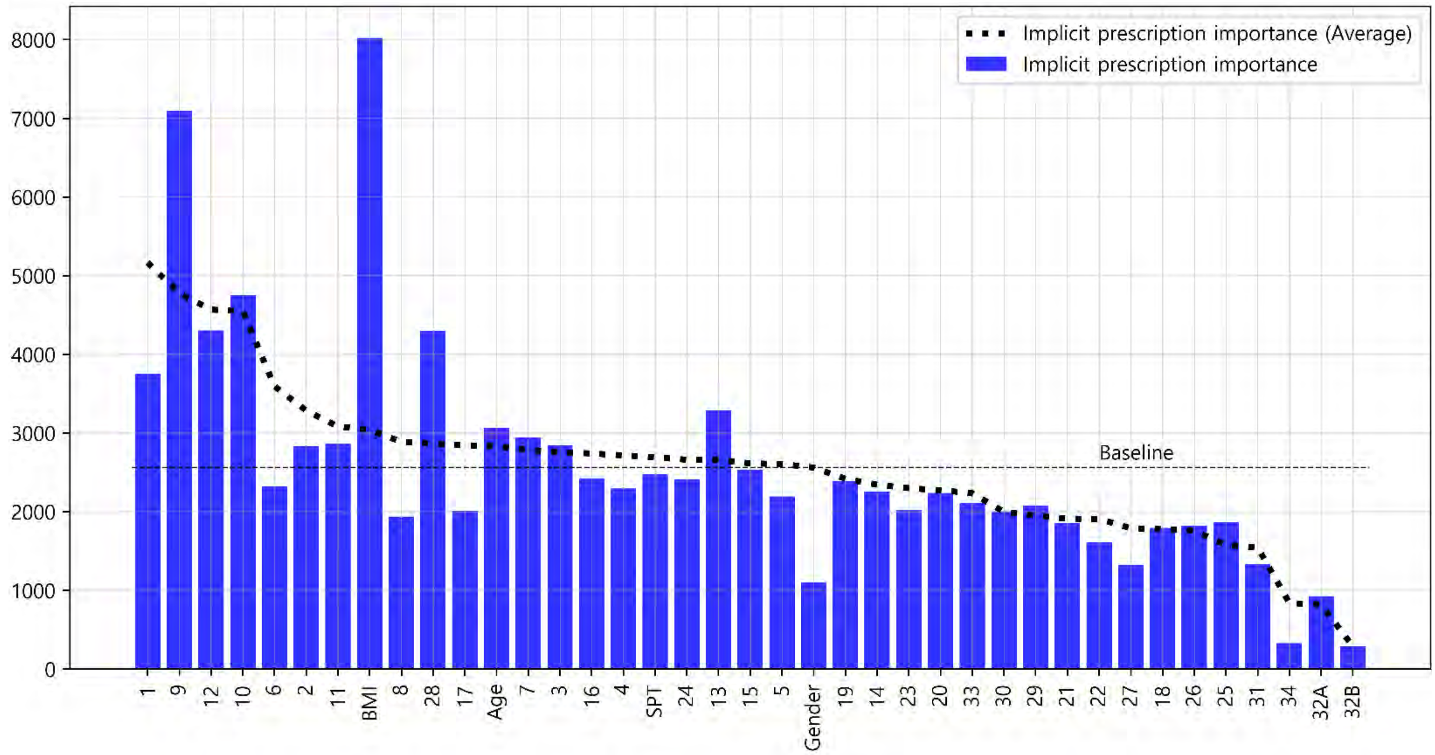

### Doctor #3

#### Glossary & Tips

1. Explicit knowledge & implicit knowledge
- Knowledge can be classified into two different categories: explicit and implicit knowledge. Explicit knowledge refers to knowledge that can be expressed in words and in a form that can be codified. In contrast, implicit knowledge refers to knowledge that an individual obtains through experience but does not convey to others.
- In this analysis, the explicit importance score was defined for the features obtained from doctors' label indicating the degree of importance of each symptom for diagnosing and treating AR patients. Implicit importance was defined based on the weights of the features learned by the machine learning algorithm when the learning algorithm effectively reproduced the doctor's decision-making process.

2. Baseline
- The baseline is the average value of the implicit importance score of individual results.

3. The explicit importance score, implicit importance score for diagnosis, and implicit importance score for prescription are visualized in orange, cyan, and blue, respectively. Individual results from each doctor and the average of all doctors' results are expressed as a bar graph and a dotted line, respectively

4. The statistic analysis was calculated by Spearman's rank correlation coefficient (p-value < 0.05).

(M1) Comparison of the individual and average explicit and implicit importance scores

To provide an overview of the differences in all importance scores, M1 was visualized with line graphs that consisted of all six scores. Scores were sorted in descending order according to the average explicit importance score of average results

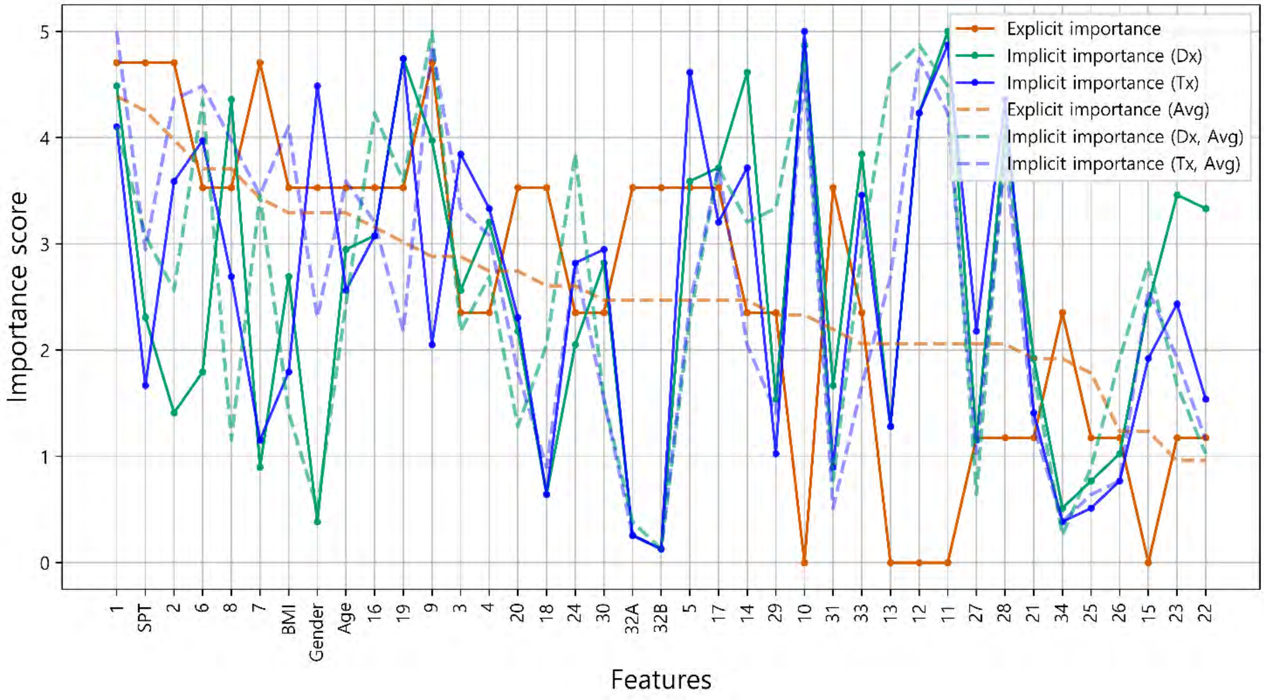


(M2) Intra-individual correlation of explicit and implicit importance

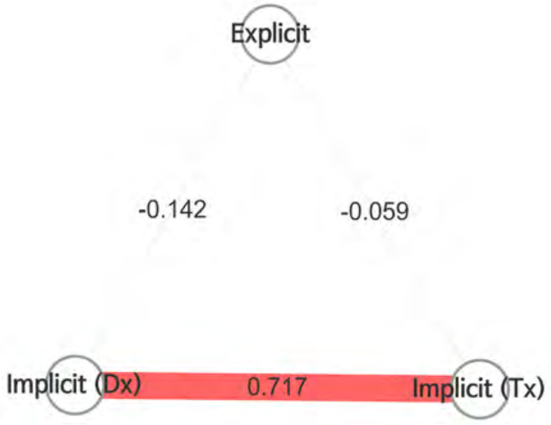


- To examine the overall correlation between the explicit and implicit knowledge of individuals, M2 was presented as nodes and edges, which represented the types of importance scores and the correlations between importance scores, respectively.
- Color of edge : the results of the Bonferroni post hoc test
  - Dark red: statistically significant
  - light red: not significant, but p-value<0.05
  - grey : p-value>0.05
- Thickness of edge (statistic): the thicker the edge, the greater the correlation.

- It is analyzed that there is no correlation between the explicit importance and implicit diagnostic importance (ρ = -0.142, p-value = 0.390). Explicit importance and implicit prescription importance have been identified as having no correlation (ρ = -0.059, p-value = 0.723).

### Doctor #3

(M3) Radar chart analysis

All the features used in the analysis were classified into eight categories and visualized as a radar chart to provide the information about the feature categories that the doctor referred to for treatment. The categories consisted of demographic features of the patient, nose symptoms, cold-hot, digestion, feces and urine, tongue diagnosis, pulse diagnosis and other symptoms

- The explicit importance score, implicit importance score for diagnosis, and implicit importance score for prescription are visualized in orange, cyan, and blue, respectively.
- Individual results from each doctor and the average of all doctors' results are represented as a solid line and a dotted line, respectively.

Comparison of Doctor 3 and average explicit importance scores

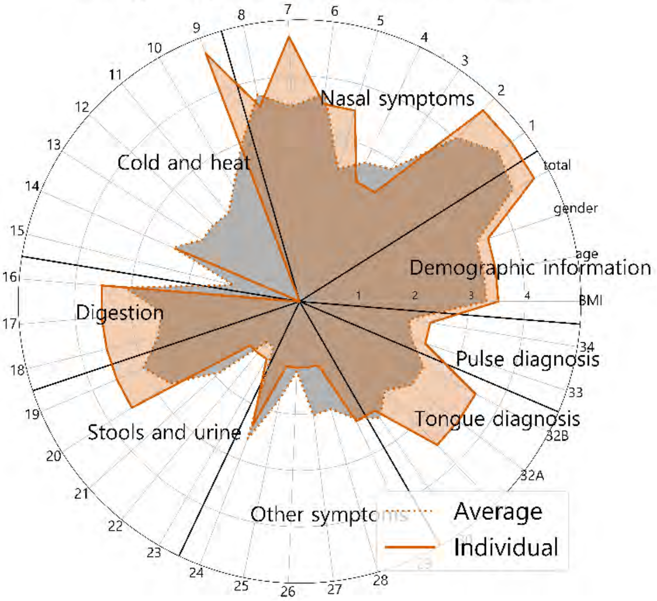


Comparison of Doctor 3 and average implicit importance scores for diagnosis

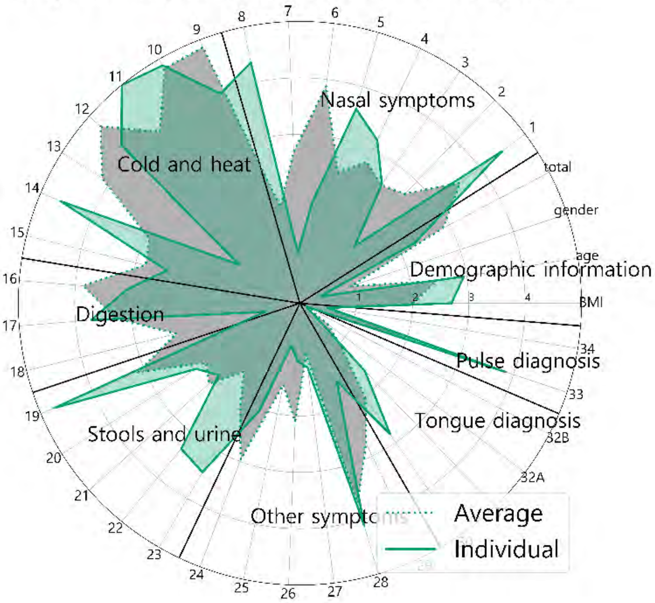


Comparison of Doctor 3 and average implicit importance scores for prescription

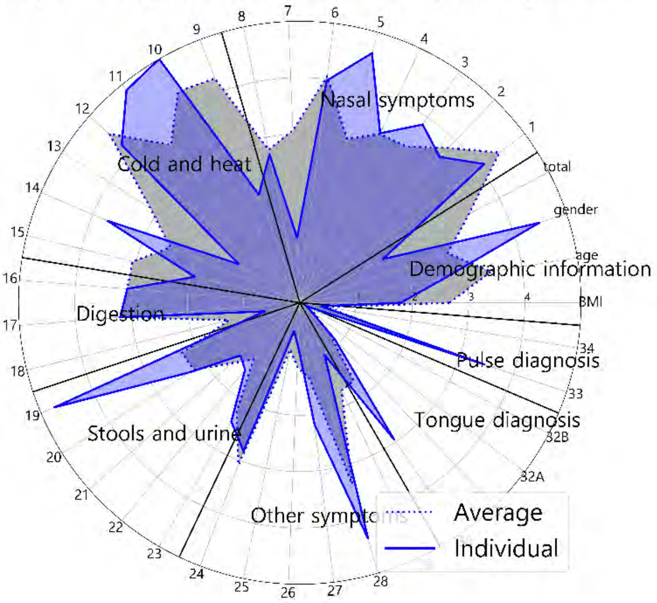


Comparison of explicit and implicit importance scores

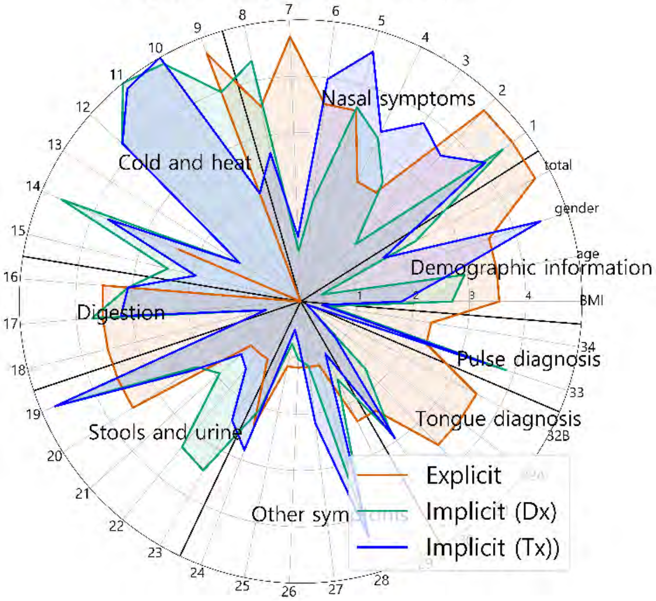


Comparison of explicit and implicit importance scores (average results)

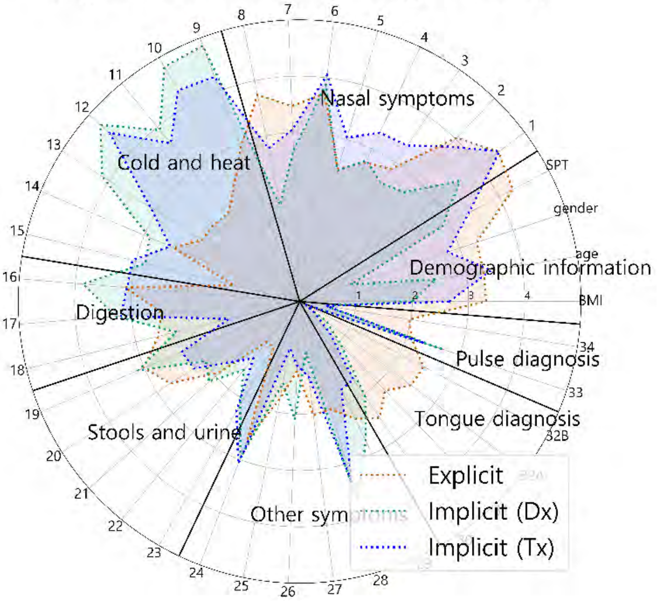

### Doctor #3

### Supplementary sheet

(S1) Intra-individual comparison of explicit importance score and implicit importance score for diagnosis

(S2) Intra-individual comparison of explicit importance score and implicit importance score for prescription

S1 and S2 were represented as scatter plots. The correlation statistics between the explicit and implicit scores were calculated using Spearman's rank correlation coefficient test. The X-axis represents the normalized value of the implicit importance and the Y-axis represents the normalized value of the explicit importance.

| Result of Spearman test | statistic | -0,142 |
|---|---|---|
| | p-value | 0.390 |

| Result of Spearman test | statistic | -0.059 |
|---|---|---|
| | p-value | 0.723 |

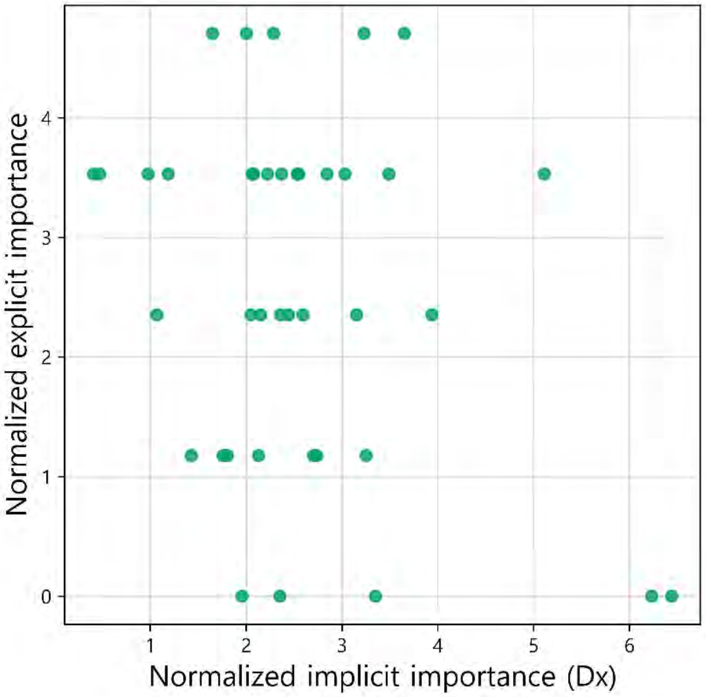


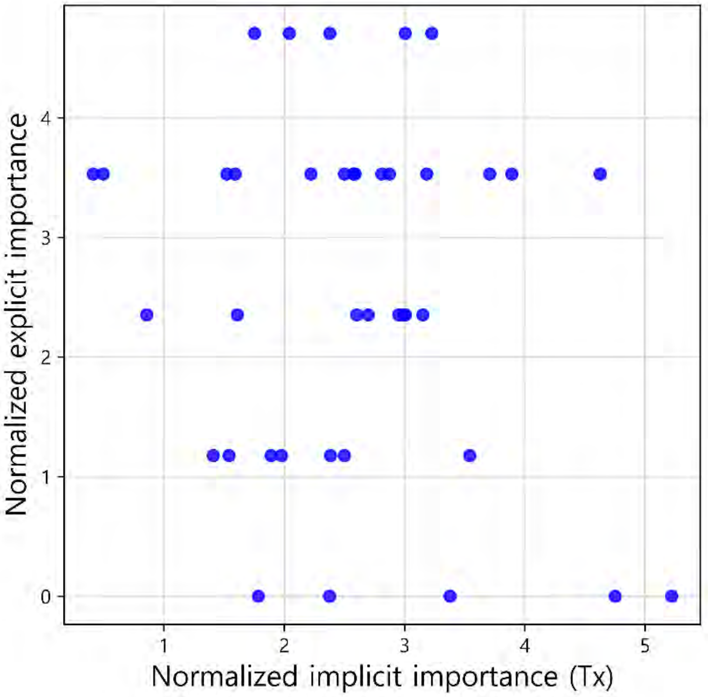


(S3) Comparison of the individual and average explicit importance scores

S3 was constructed to compare the average importance score of the doctors as a group with the individual doctor's importance score. Individual results from each doctor and the average of all doctors' results are expressed as a bar graph and a dotted line, respectively. The graphs are sorted in descending order according to the importance score of the average results.

| Result of Spearman test | |
|---|---|
| statistic | 0.826 |
| p-value | < 0.001 |

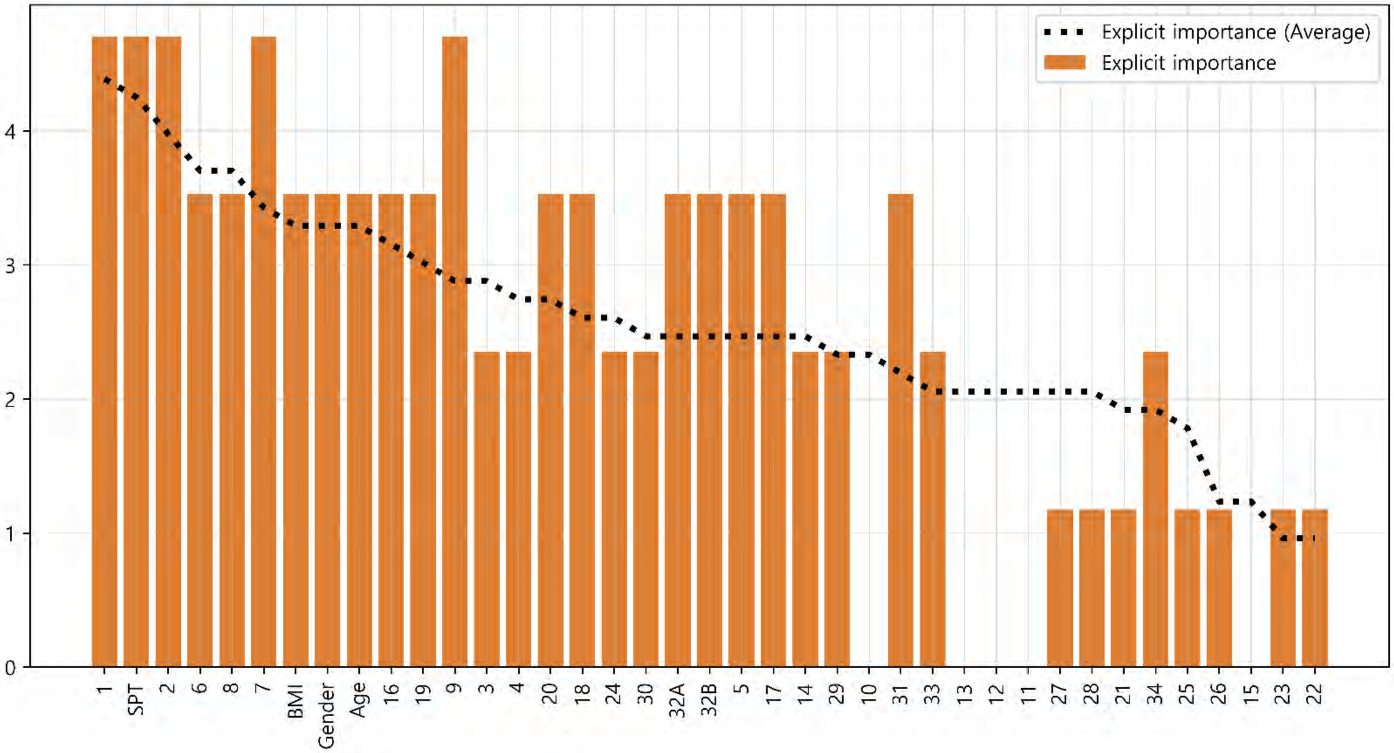

### Doctor #3

### Supplementary sheet

(S4) Comparison of the individual and average implicit importance scores for diagnosis

S4 was constructed to compare the average importance score of the doctors as a group with the individual doctor's importance score. Individual results from each doctor and the average of all doctors' results are expressed as a bar graph and a dotted line, respectively. The graphs are sorted in descending order according to the importance score of the average results.

| Result of Spearman test | |
|---|---|
| statistic | 0.573 |
| p-value | < 0.001 |

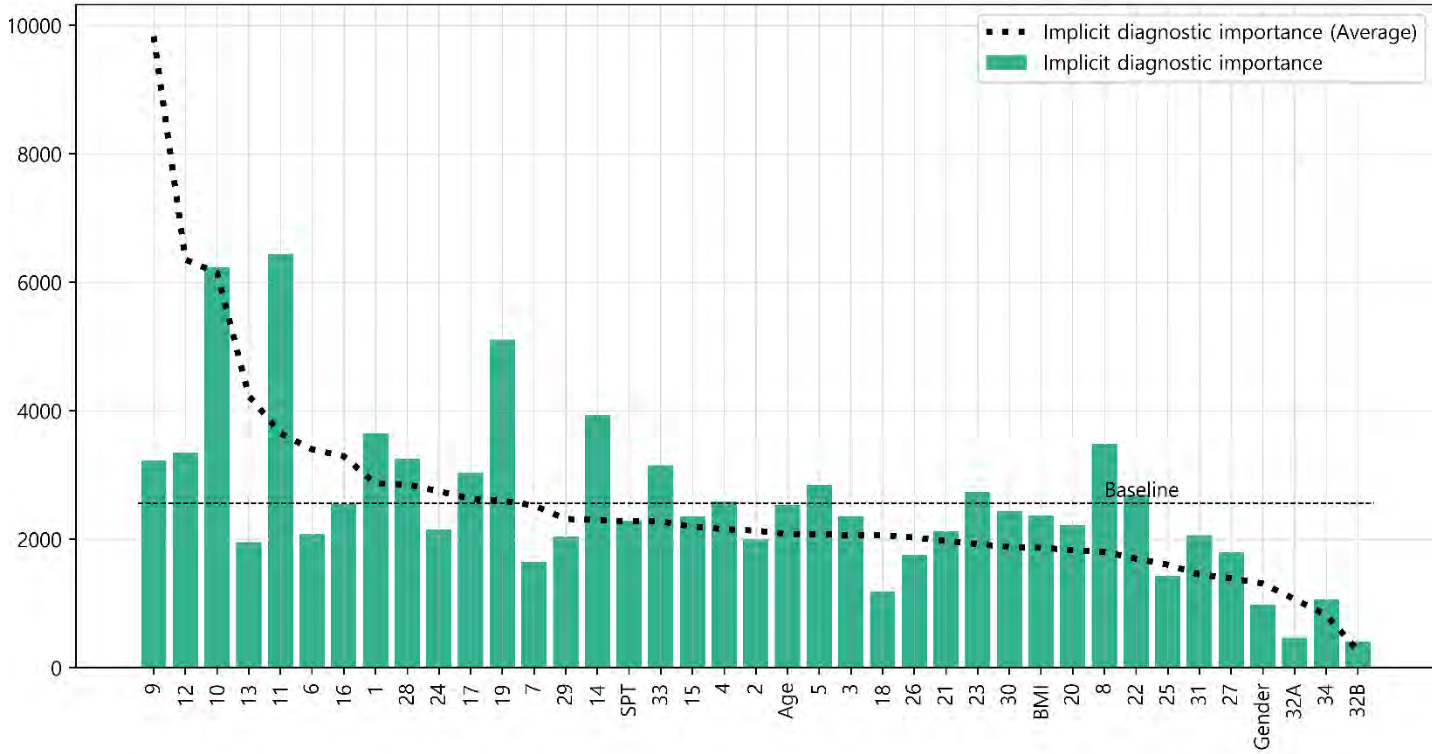


(S5) Comparison of the individual and average implicit importance scores for prescription

S5 was constructed to compare the average importance score of the doctors as a group with the individual doctor's importance score. Individual results from each doctor and the average of all doctors' results are expressed as a bar graph and a dotted line, respectively. The graphs are sorted in descending order according to the importance score of the average results.

| Result of Spearman test | |
|---|---|
| statistic | 0.660 |
| p-value | < 0.001 |

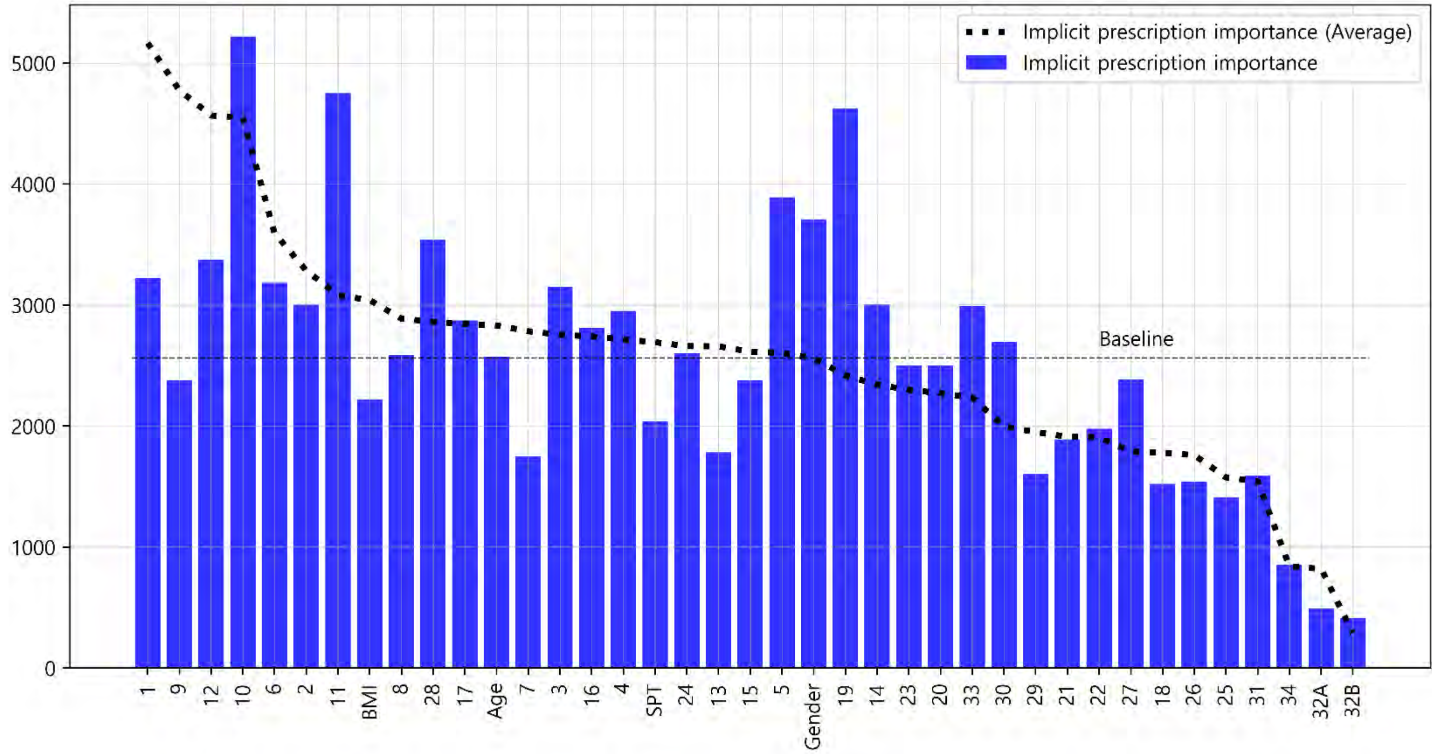

### Doctor #4

#### Glossary & Tips

1. Explicit knowledge & implicit knowledge

- Knowledge can be classified into two different categories: explicit and implicit knowledge. Explicit knowledge refers to knowledge that can be expressed in words and in a form that can be codified. In contrast, implicit knowledge refers to knowledge that an individual obtains through experience but does not convey to others.
- In this analysis, the explicit importance score was defined for the features obtained from doctors' label indicating the degree of importance of each symptom for diagnosing and treating AR patients. Implicit importance was defined based on the weights of the features learned by the machine learning algorithm when the learning algorithm effectively reproduced the doctor's decision-making process.

2. Baseline

- The baseline is the average value of the implicit importance score of individual results.

3. The explicit importance score, implicit importance score for diagnosis, and implicit importance score for prescription are visualized in orange, cyan, and blue, respectively. Individual results from each doctor and the average of all doctors' results are expressed as a bar graph and a dotted line, respectively

4. The statistic analysis was calculated by Spearman's rank correlation coefficient (p-value < 0.05).

(M1) Comparison of the individual and average explicit and implicit importance scores

To provide an overview of the differences in all importance scores, M1 was visualized with line graphs that consisted of all six scores. Scores were sorted in descending order according to the average explicit importance score of average results

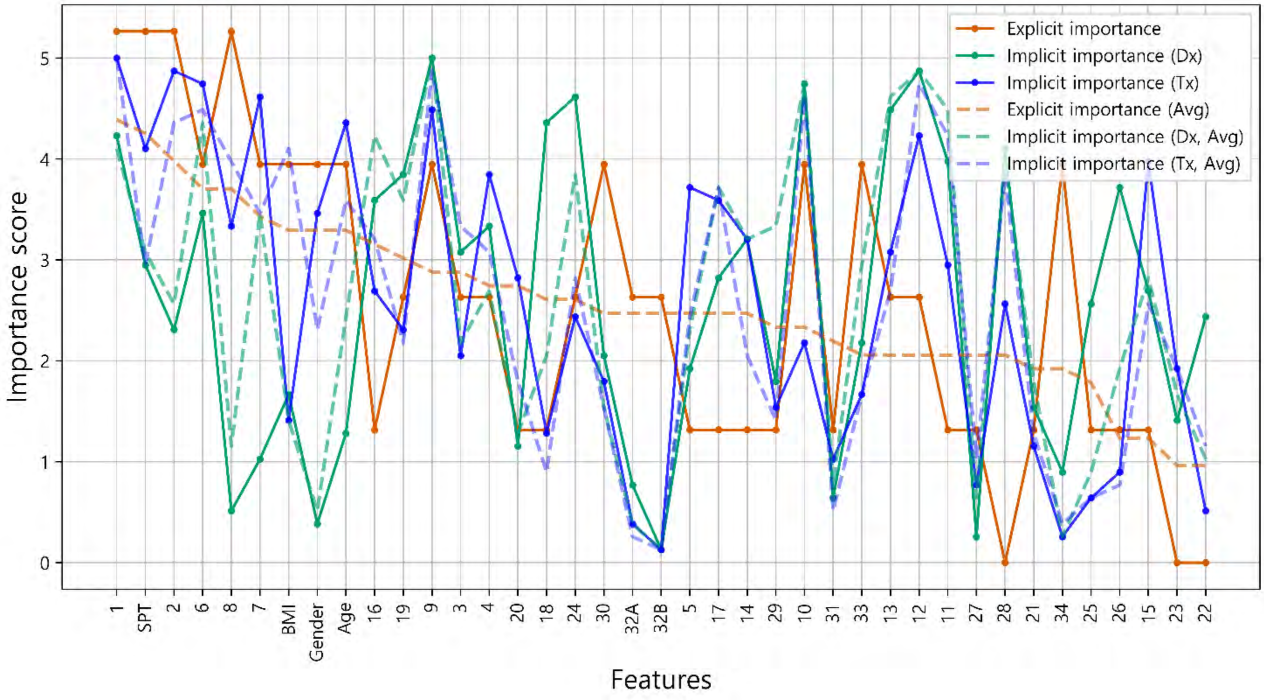


(M2) Intra-individual correlation of explicit and implicit importance

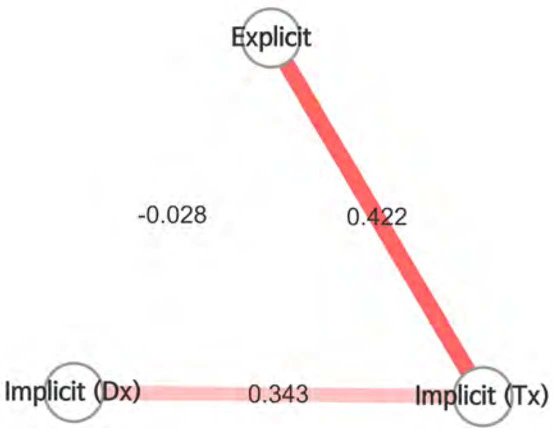


- To examine the overall correlation between the explicit and implicit knowledge of individuals, M2 was presented as nodes and edges, which represented the types of importance scores and the correlations between importance scores, respectively.
- Color of edge : the results of the Bonferroni post hoc test
  - Dark red: statistically significant
  - light red: not significant, but p-value<0.05
  - grey : p-value>0.05
- Thickness of edge (statistic): the thicker the edge, the greater the correlation.

- It is analyzed that there is no correlation between the explicit importance and implicit diagnostic importance (ρ = -0.028, p-value = 0.864). Explicit importance and implicit prescription importance have been identified as having a weak correlation (ρ = 0.422, p-value = 0.007).

### Doctor #4

#### (M3) Radar chart analysis

All the features used in the analysis were classified into eight categories and visualized as a radar chart to provide the information about the feature categories that the doctor referred to for treatment. The categories consisted of demographic features of the patient, nose symptoms, cold-hot, digestion, feces and urine, tongue diagnosis, pulse diagnosis and other symptoms

- The explicit importance score, implicit importance score for diagnosis, and implicit importance score for prescription are visualized in orange, cyan, and blue, respectively.
- Individual results from each doctor and the average of all doctors' results are represented as a solid line and a dotted line, respectively.

Comparison of Doctor 4 and average explicit importance scores

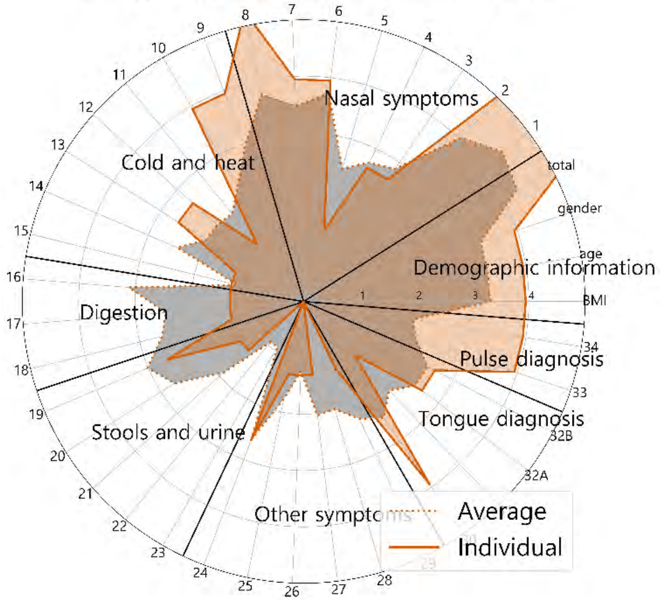


Comparison of Doctor 4 and average implicit importance scores for diagnosis

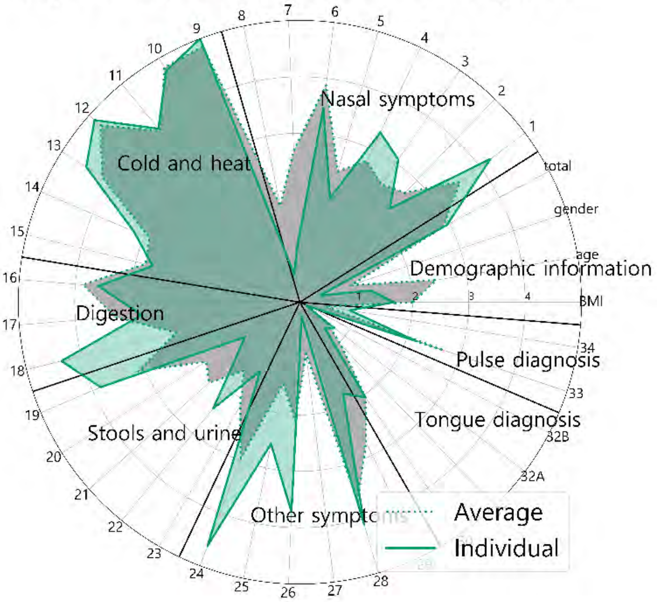


Comparison of Doctor 4 and average implicit importance scores for prescription

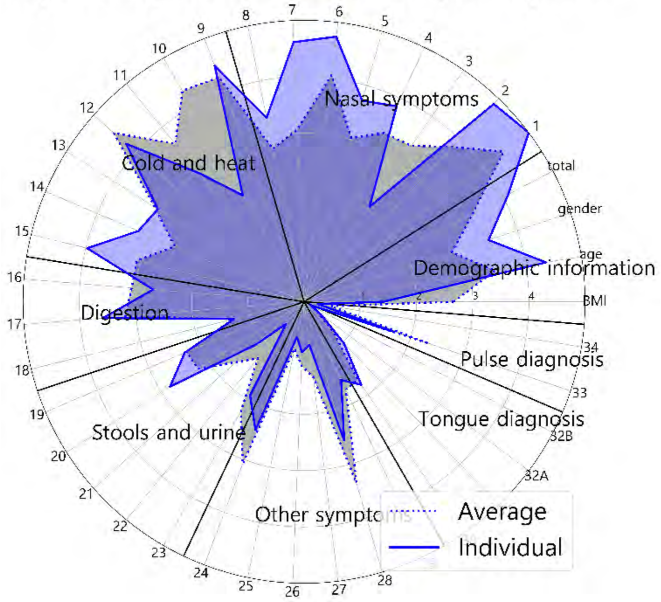


Comparison of explicit and implicit importance scores

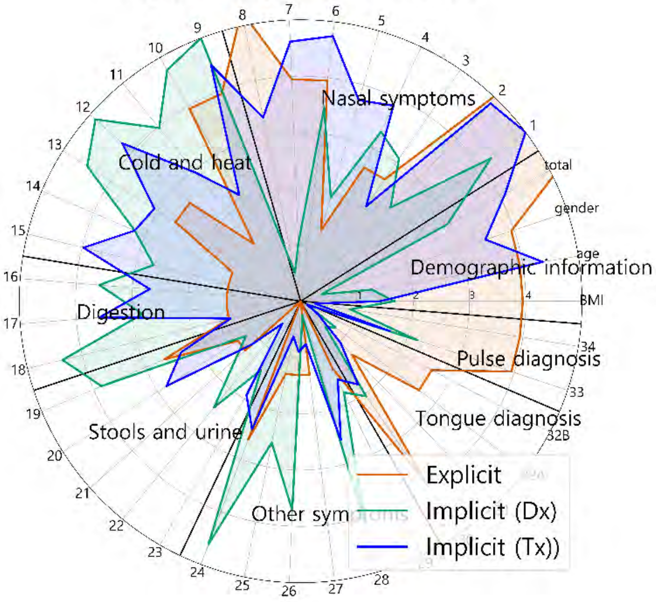


Comparison of explicit and implicit importance scores (average results)

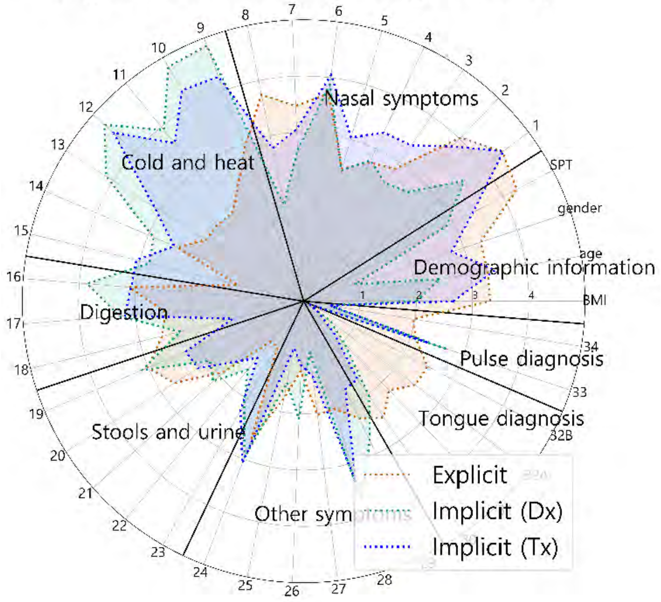

### Doctor #4

#### Supplementary sheet

(S1) Intra-individual comparison of explicit importance score and implicit importance score for diagnosis

(S2) Intra-individual comparison of explicit importance score and implicit importance score for prescription

S1 and S2 were represented as scatter plots. The correlation statistics between the explicit and implicit scores were calculated using Spearman's rank correlation coefficient test. The X-axis represents the normalized value of the implicit importance and the Y-axis represents the normalized value of the explicit importance.

| Result of Spearman test | | |
|---|---|---|
| | statistic | -0.028 |
| | p-value | 0.864 |

| Result of Spearman test | | |
|---|---|---|
| | statistic | 0.422 |
| | p-value | 0.007 |

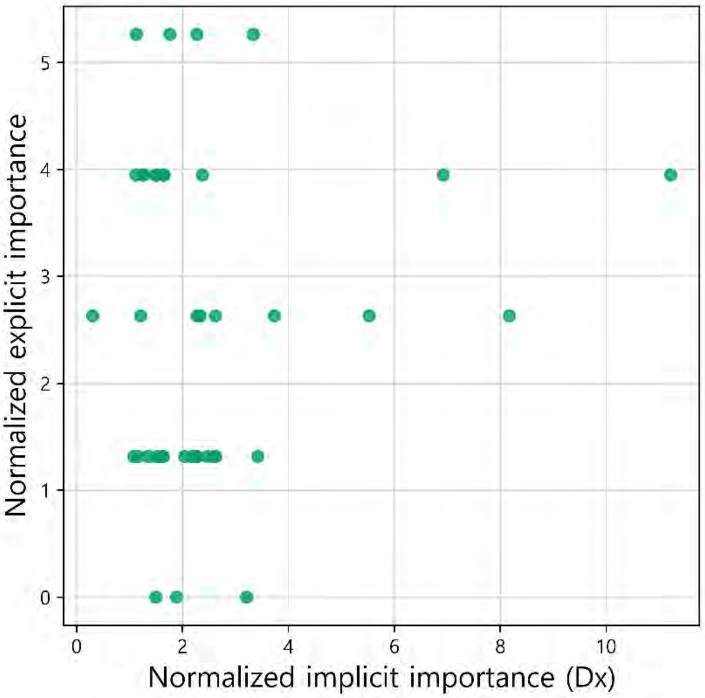


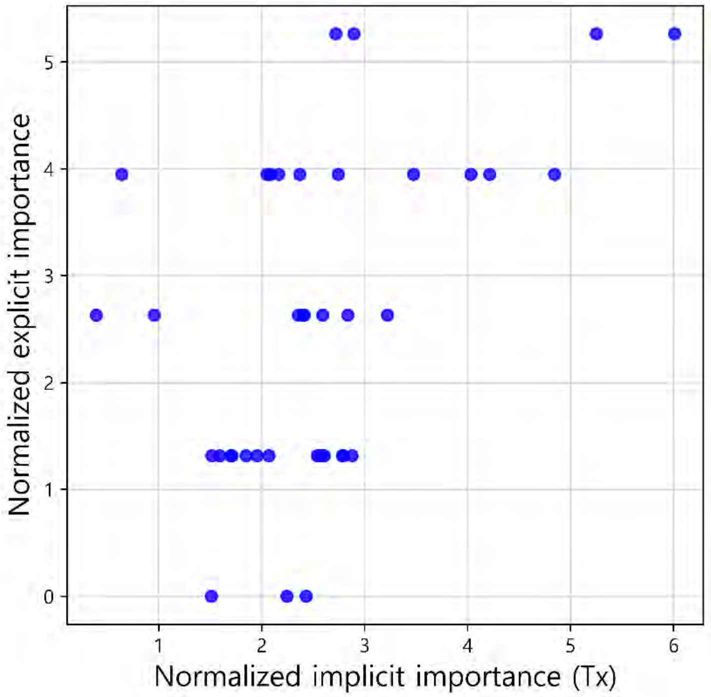


(S3) Comparison of the individual and average explicit importance scores

S3 was constructed to compare the average importance score of the doctors as a group with the individual doctor's importance score. Individual results from each doctor and the average of all doctors' results are expressed as a bar graph and a dotted line, respectively. The graphs are sorted in descending order according to the importance score of the average results.

| Result of Spearman test | |
|---|---|
| statistic | 0.682 |
| p-value | < 0.001 |

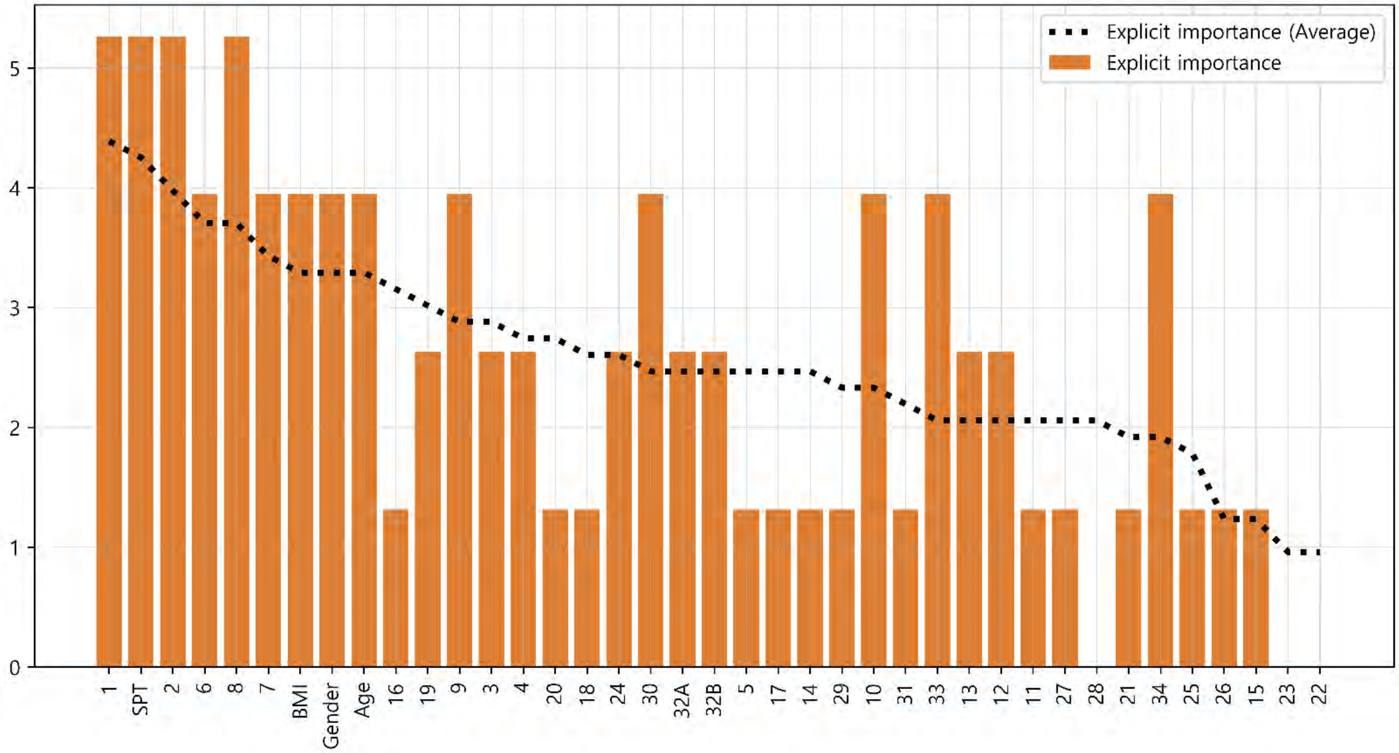

(S4) Comparison of the individual and average implicit importance scores for diagnosis

S4 was constructed to compare the average importance score of the doctors as a group with the individual doctor's importance score. Individual results from each doctor and the average of all doctors' results are expressed as a bar graph and a dotted line, respectively. The graphs are sorted in descending order according to the importance score of the average results.

| Result of Spearman test | |
|---|---|
| statistic | 0.818 |
| p-value | < 0.001 |

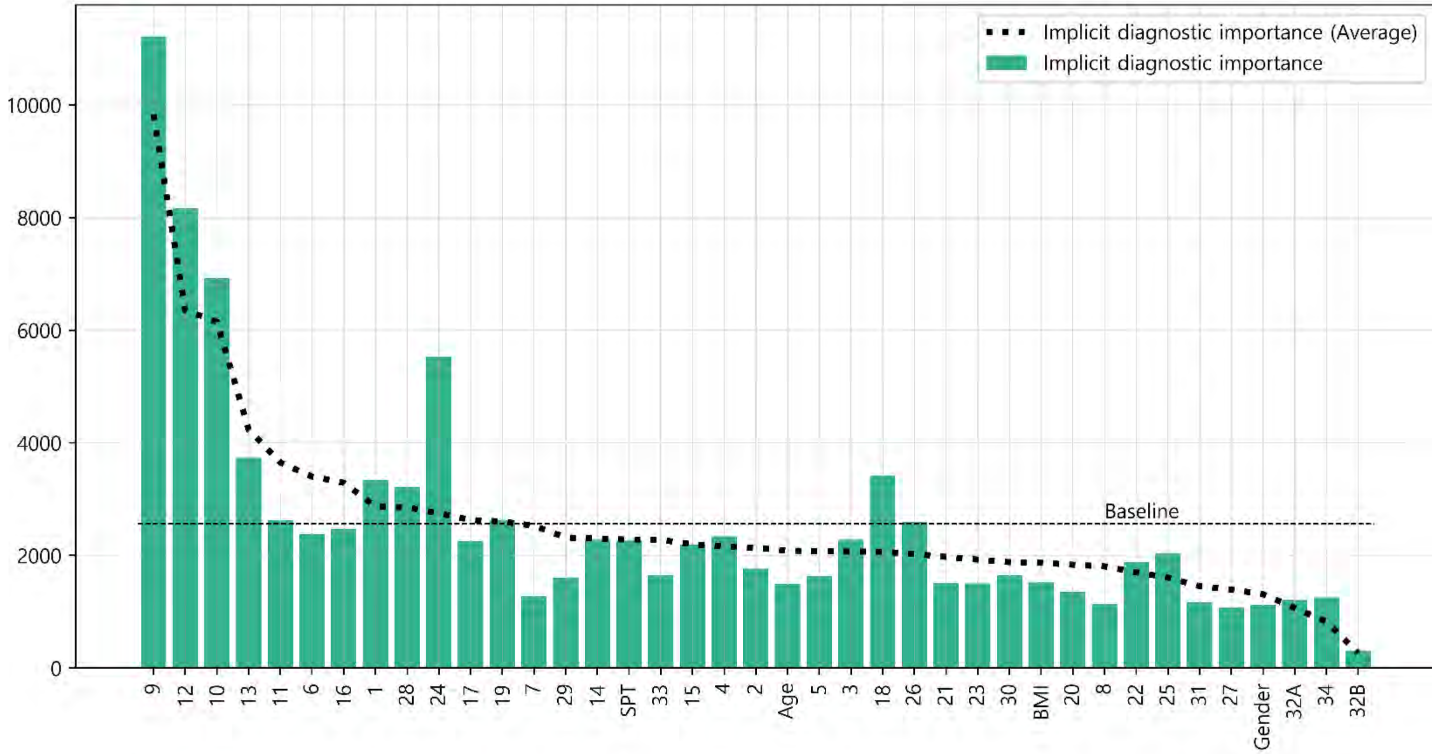


(S5) Comparison of the individual and average implicit importance scores for prescription

S5 was constructed to compare the average importance score of the doctors as a group with the individual doctor's importance score. Individual results from each doctor and the average of all doctors' results are expressed as a bar graph and a dotted line, respectively. The graphs are sorted in descending order according to the importance score of the average results.

| Result of Spearman test | |
|---|---|
| statistic | 0.802 |
| p-value | < 0.001 |

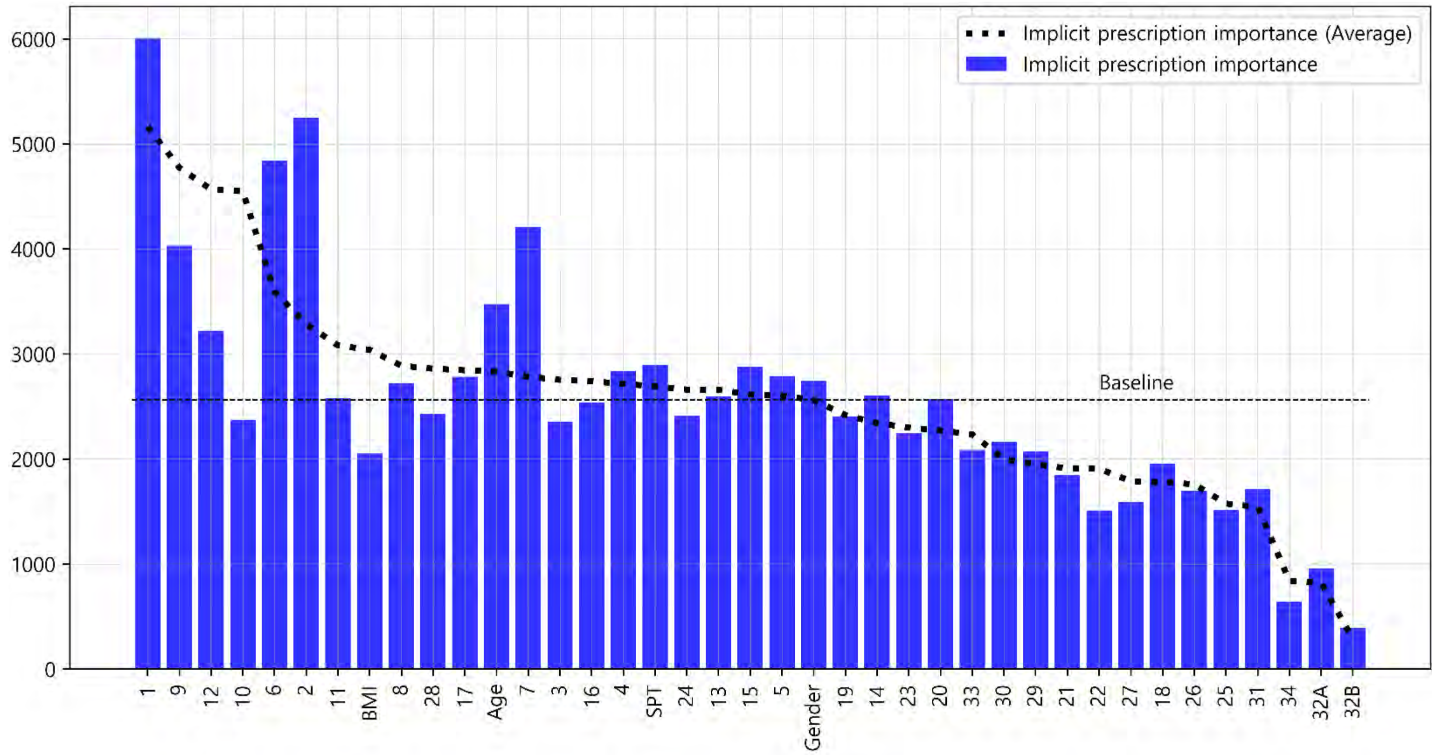

### Doctor #5

#### Glossary & Tips

1. Explicit knowledge & implicit knowledge

- Knowledge can be classified into two different categories: explicit and implicit knowledge. Explicit knowledge refers to knowledge that can be expressed in words and in a form that can be codified. In contrast, implicit knowledge refers to knowledge that an individual obtains through experience but does not convey to others.
- In this analysis, the explicit importance score was defined for the features obtained from doctors' label indicating the degree of importance of each symptom for diagnosing and treating AR patients. Implicit importance was defined based on the weights of the features learned by the machine learning algorithm when the learning algorithm effectively reproduced the doctor's decision-making process.

2. Baseline

- The baseline is the average value of the implicit importance score of individual results.

3. The explicit importance score, implicit importance score for diagnosis, and implicit importance score for prescription are visualized in orange, cyan, and blue, respectively. Individual results from each doctor and the average of all doctors' results are expressed as a bar graph and a dotted line, respectively

4. The statistic analysis was calculated by Spearman's rank correlation coefficient (p-value < 0.05).

(M1) Comparison of the individual and average explicit and implicit importance scores

To provide an overview of the differences in all importance scores, M1 was visualized with line graphs that consisted of all six scores. Scores were sorted in descending order according to the average explicit importance score of average results

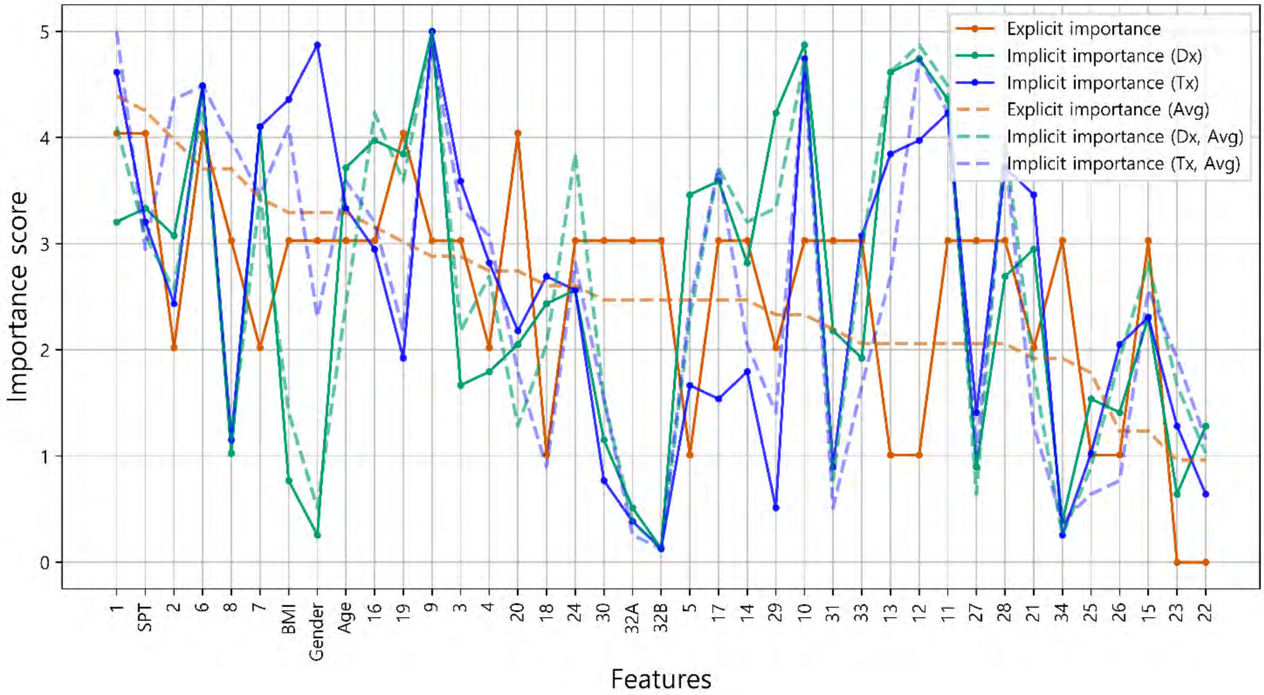


(M2) Intra-individual correlation of explicit and implicit importance

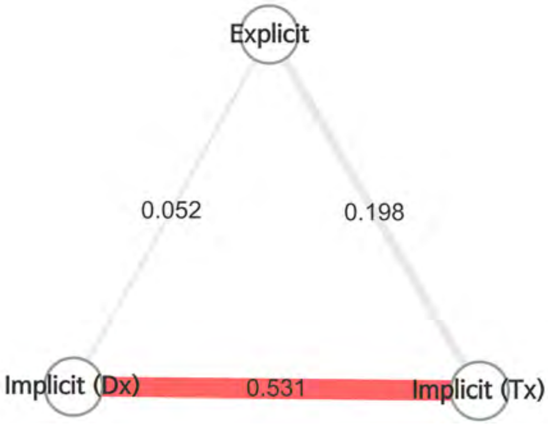


- To examine the overall correlation between the explicit and implicit knowledge of individuals, M2 was presented as nodes and edges, which represented the types of importance scores and the correlations between importance scores, respectively.
- Color of edge : the results of the Bonferroni post hoc test
  - Dark red: statistically significant
  - light red: not significant, but p-value<0.05
  - grey : p-value>0.05
- Thickness of edge (statistic): the thicker the edge, the greater the correlation.

- It is analyzed that there is no correlation between the explicit importance and implicit diagnostic importance (ρ = 0.052, p-value = 0.755). Explicit importance and implicit prescription importance have been identified as having no correlation(ρ = 0.198, p-value = 0.228).

### Doctor #5 (M3) Radar chart analysis

All the features used in the analysis were classified into eight categories and visualized as a radar chart to provide the information about the feature categories that the doctor referred to for treatment. The categories consisted of demographic features of the patient, nose symptoms, cold-hot, digestion, feces and urine, tongue diagnosis, pulse diagnosis and other symptoms

- The explicit importance score, implicit importance score for diagnosis, and implicit importance score for prescription are visualized in orange, cyan, and blue, respectively.
- Individual results from each doctor and the average of all doctors' results are represented as a solid line and a dotted line, respectively.

Comparison of Doctor 5 and average explicit importance scores

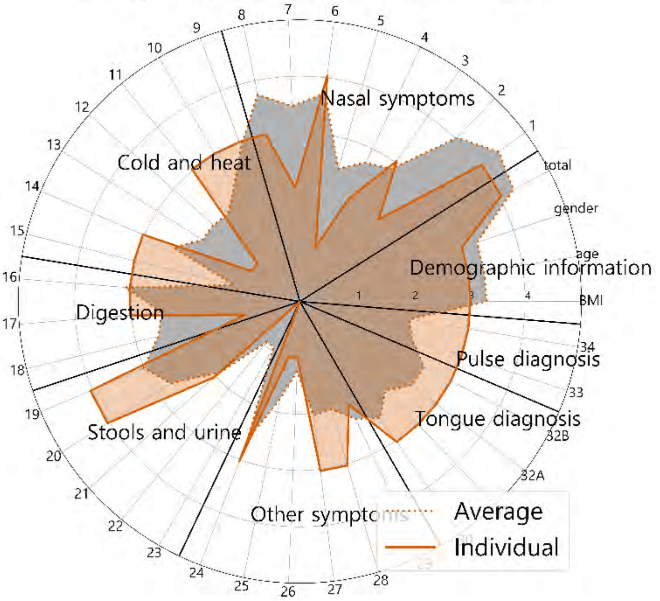


Comparison of Doctor 5 and average implicit importance scores for diagnosis

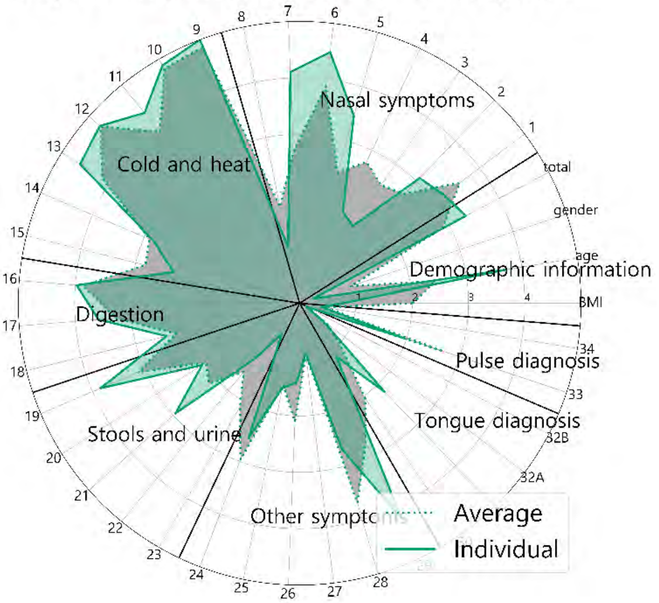


Comparison of Doctor 5 and average implicit importance scores for prescription

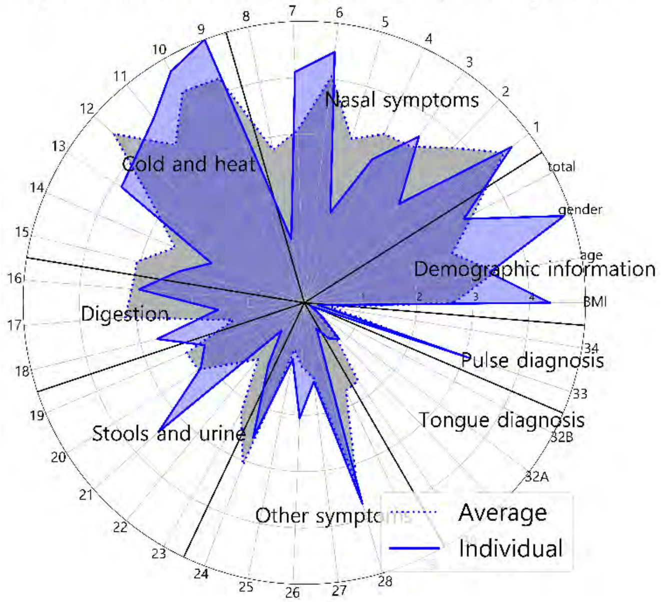


Comparison of explicit and implicit importance scores

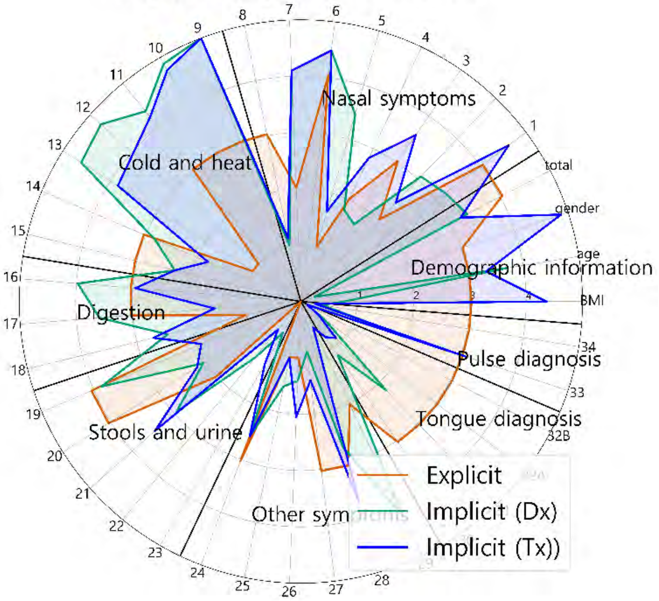


Comparison of explicit and implicit importance scores (average results)

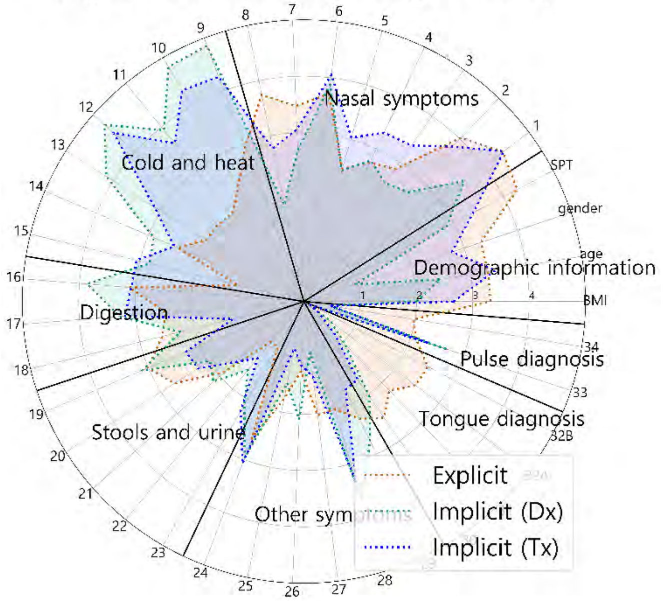

### Doctor #5



(S1) Intra-individual comparison of explicit importance score and implicit importance score for diagnosis

(S2) Intra-individual comparison of explicit importance score and implicit importance score for prescription

S1 and S2 were represented as scatter plots. The correlation statistics between the explicit and implicit scores were calculated using Spearman's rank correlation coefficient test. The X-axis represents the normalized value of the implicit importance and the Y-axis represents the normalized value of the explicit importance.

| Result of Spearman test | | |
|---|---|---|
| | statistic | 0.052 |
| | p-value | 0.755 |

| Result of Spearman test | | |
|---|---|---|
| | statistic | 0.195 |
| | p-value | 0.228 |

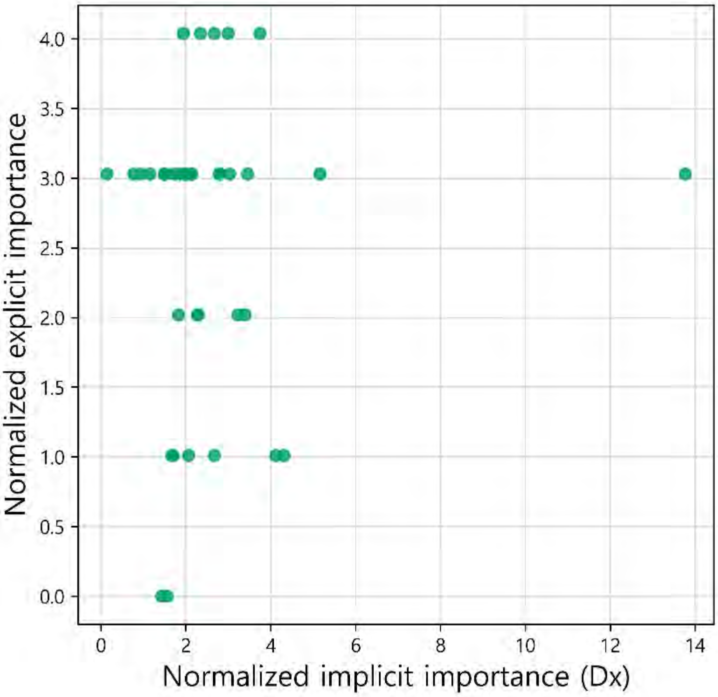


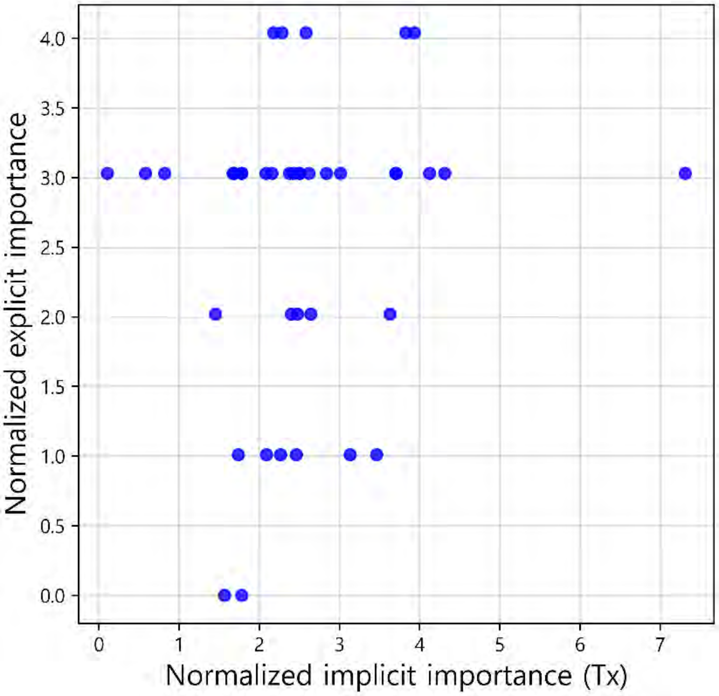


(S3) Comparison of the individual and average explicit importance scores

S3 was constructed to compare the average importance score of the doctors as a group with the individual doctor's importance score. Individual results from each doctor and the average of all doctors' results are expressed as a bar graph and a dotted line, respectively. The graphs are sorted in descending order according to the importance score of the average results.

| Result of Spearman test | |
|---|---|
| statistic | 0.520 |
| p-value | 0.001 |

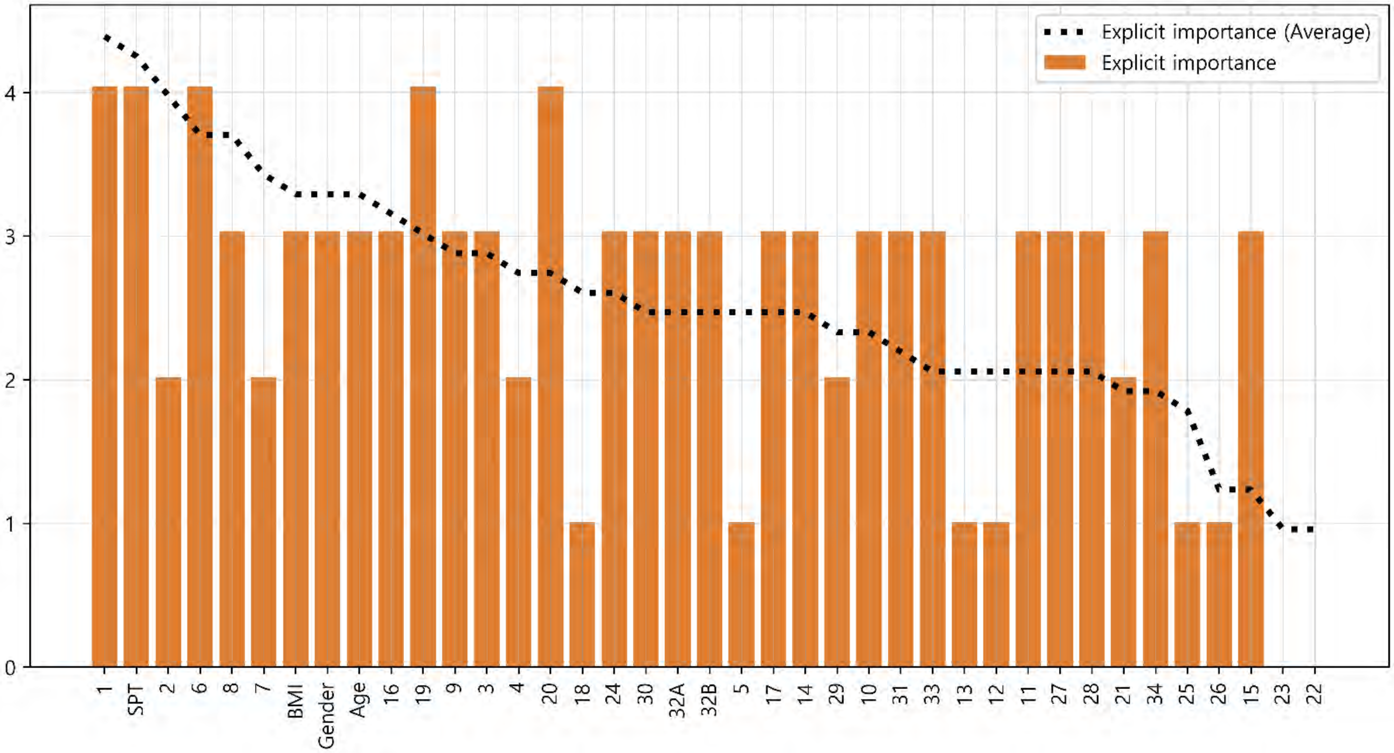

#### Doctor #5

#### Supplementary sheet

##### (S4) Comparison of the individual and average implicit importance scores for diagnosis

S4 was constructed to compare the average importance score of the doctors as a group with the individual doctor's importance score. Individual results from each doctor and the average of all doctors' results are expressed as a bar graph and a dotted line, respectively. The graphs are sorted in descending order according to the importance score of the average results.

| Result of Spearman test | |
|---|---|
| statistic | 0.889 |
| p-value | < 0.001 |

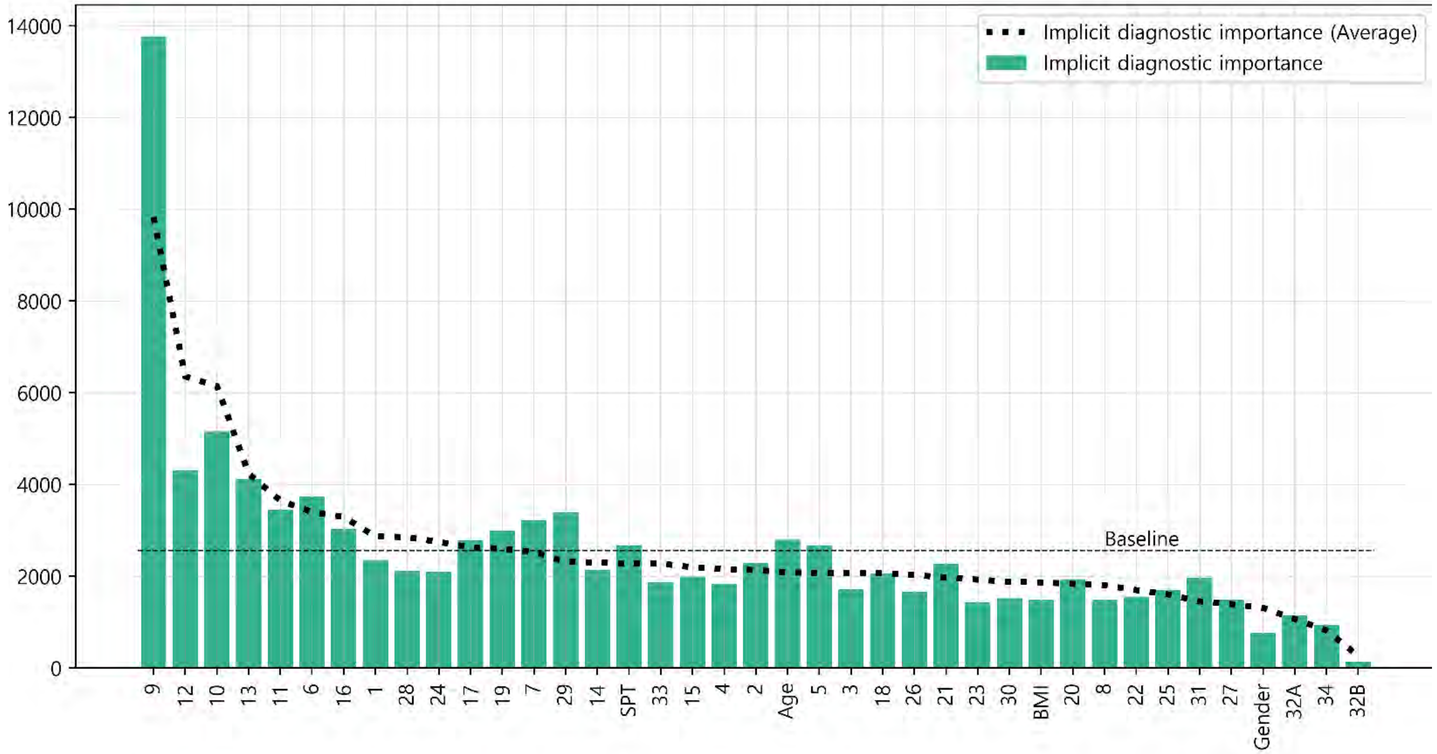


##### (S5) Comparison of the individual and average implicit importance scores for prescription

S5 was constructed to compare the average importance score of the doctors as a group with the individual doctor's importance score. Individual results from each doctor and the average of all doctors' results are expressed as a bar graph and a dotted line, respectively. The graphs are sorted in descending order according to the importance score of the average results.

| Result of Spearman test | |
|---|---|
| statistic | 0.747 |
| p-value | < 0.001 |

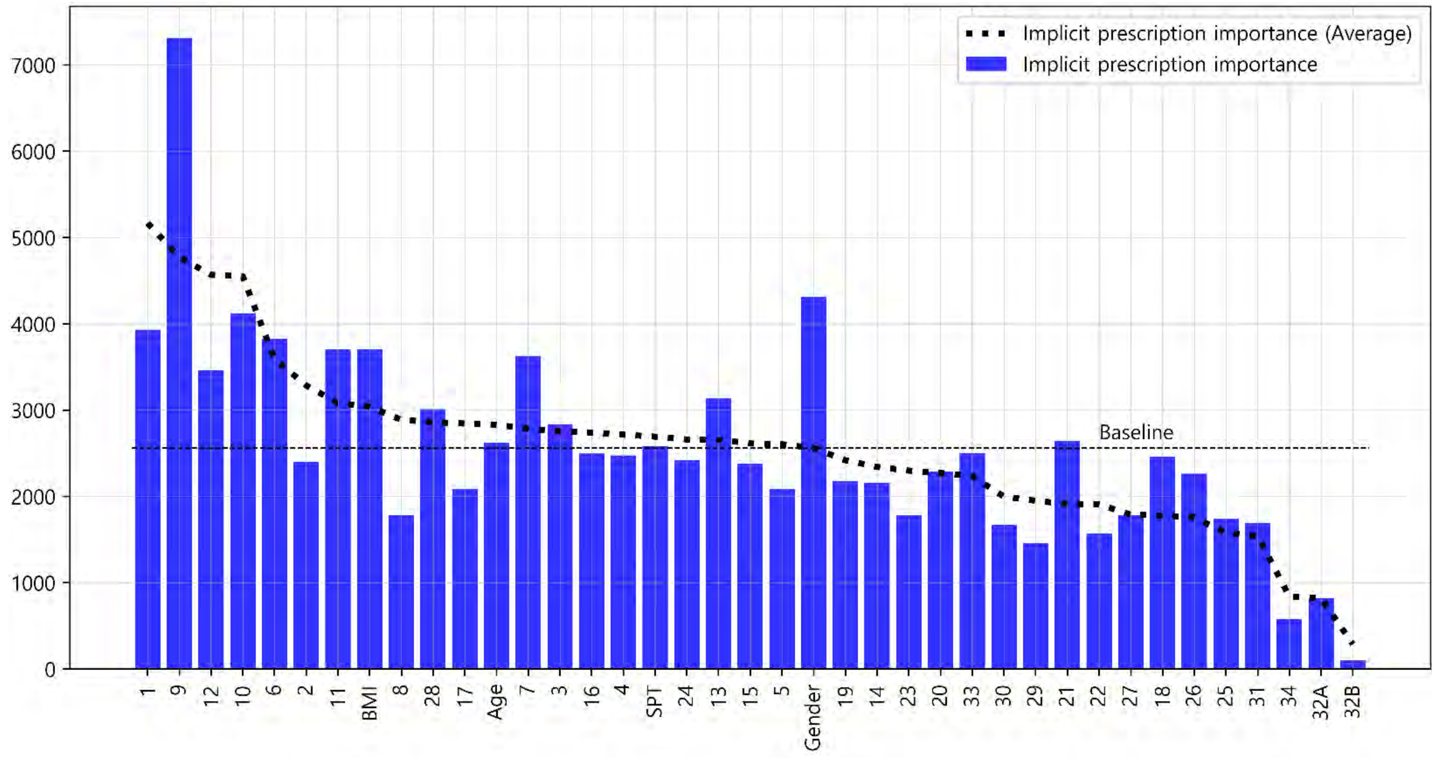

### Doctor #6

#### Glossary & Tips

1. Explicit knowledge & implicit knowledge

- Knowledge can be classified into two different categories: explicit and implicit knowledge. Explicit knowledge refers to knowledge that can be expressed in words and in a form that can be codified. In contrast, implicit knowledge refers to knowledge that an individual obtains through experience but does not convey to others.
- In this analysis, the explicit importance score was defined for the features obtained from doctors' label indicating the degree of importance of each symptom for diagnosing and treating AR patients. Implicit importance was defined based on the weights of the features learned by the machine learning algorithm when the learning algorithm effectively reproduced the doctor's decision-making process.

2. Baseline

- The baseline is the average value of the implicit importance score of individual results.

3. The explicit importance score, implicit importance score for diagnosis, and implicit importance score for prescription are visualized in orange, cyan, and blue, respectively. Individual results from each doctor and the average of all doctors' results are expressed as a bar graph and a dotted line, respectively

4. The statistic analysis was calculated by Spearman's rank correlation coefficient (p-value < 0.05).

(M1) Comparison of the individual and average explicit and implicit importance scores

To provide an overview of the differences in all importance scores, M1 was visualized with line graphs that consisted of all six scores. Scores were sorted in descending order according to the average explicit importance score of average results

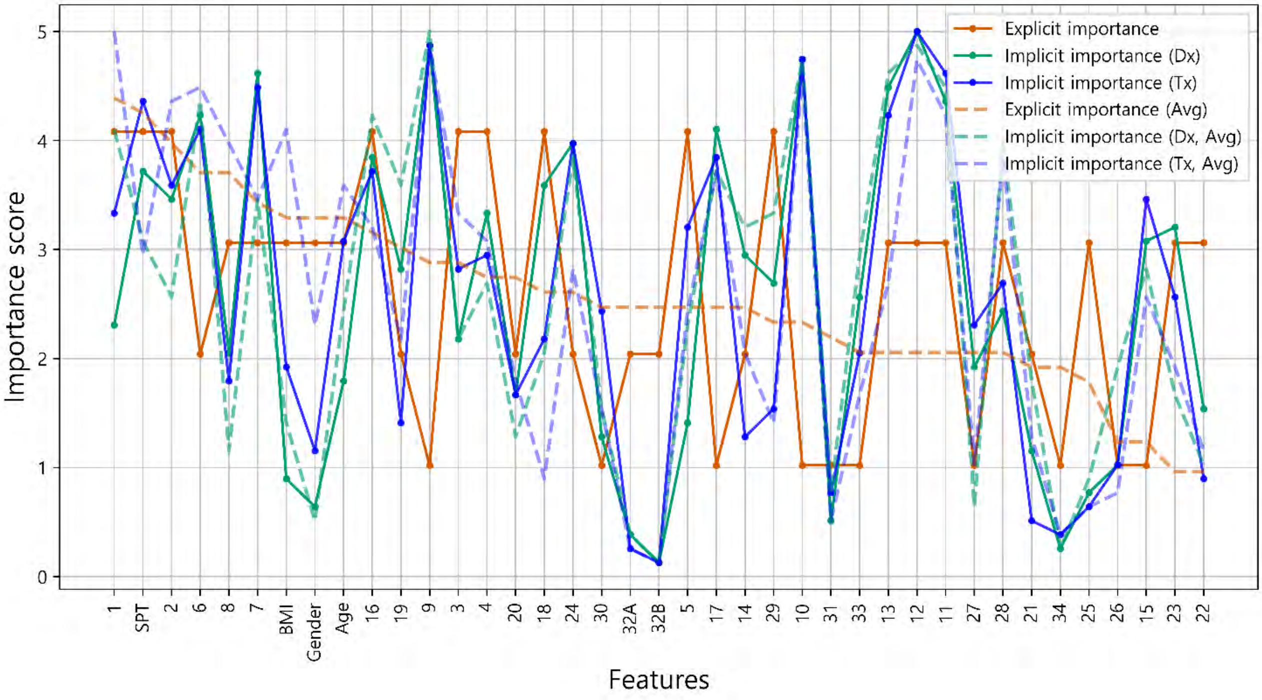


(M2) Intra-individual correlation of explicit and implicit importance

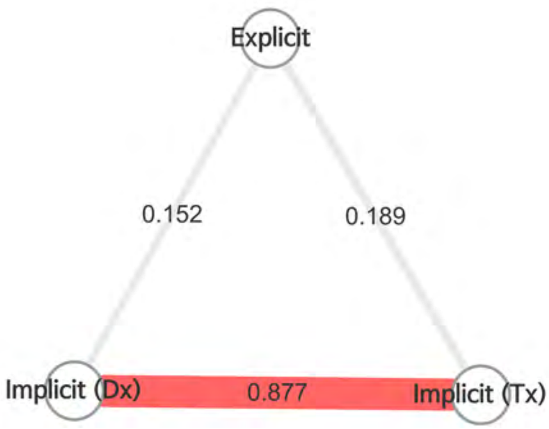


- To examine the overall correlation between the explicit and implicit knowledge of individuals, M2 was presented as nodes and edges, which represented the types of importance scores and the correlations between importance scores, respectively.
- Color of edge : the results of the Bonferroni post hoc test
  - Dark red: statistically significant
  - light red: not significant, but p-value<0.05
  - grey : p-value>0.05
- Thickness of edge (statistic): the thicker the edge, the greater the correlation.

- It is analyzed that there is no correlation between the explicit importance and implicit diagnostic importance (ρ = 0.152, p-value = 0.357). Explicit importance and implicit prescription importance have been identified as having no correlation (ρ = 0.189, p-value = 0.250).

### Doctor #6 (M3) Radar chart analysis

All the features used in the analysis were classified into eight categories and visualized as a radar chart to provide the information about the feature categories that the doctor referred to for treatment. The categories consisted of demographic features of the patient, nose symptoms, cold-hot, digestion, feces and urine, tongue diagnosis, pulse diagnosis and other symptoms

- The explicit importance score, implicit importance score for diagnosis, and implicit importance score for prescription are visualized in orange, cyan, and blue, respectively.
- Individual results from each doctor and the average of all doctors' results are represented as a solid line and a dotted line, respectively.

Comparison of Doctor 6 and average explicit importance scores

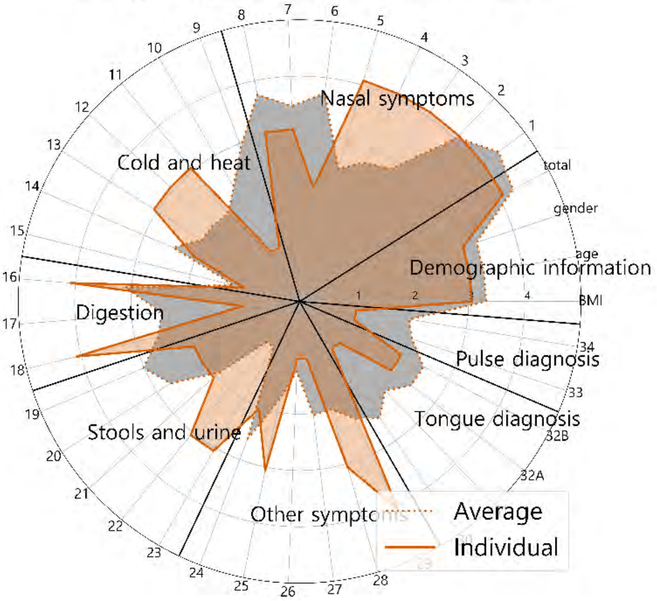


Comparison of Doctor 6 and average implicit importance scores for diagnosis

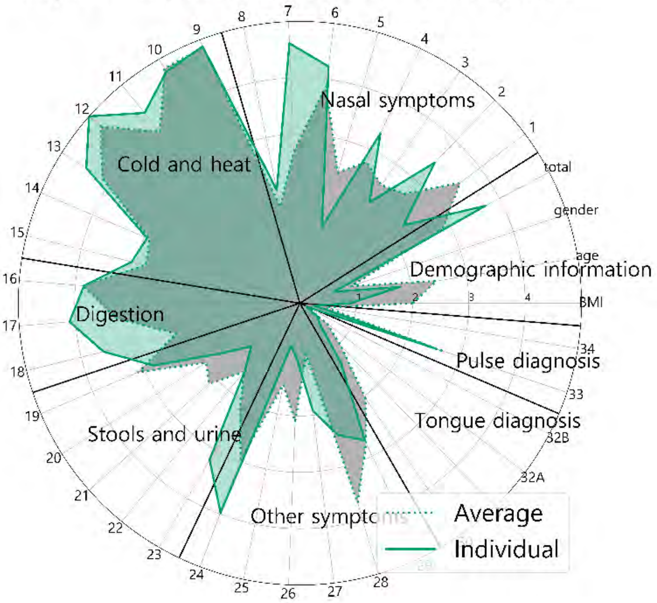


Comparison of Doctor 6 and average implicit importance scores for prescription

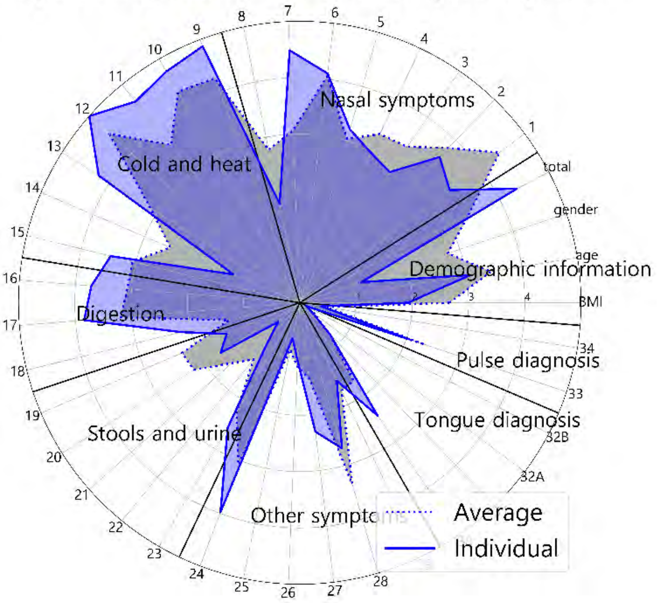


Comparison of explicit and implicit importance scores

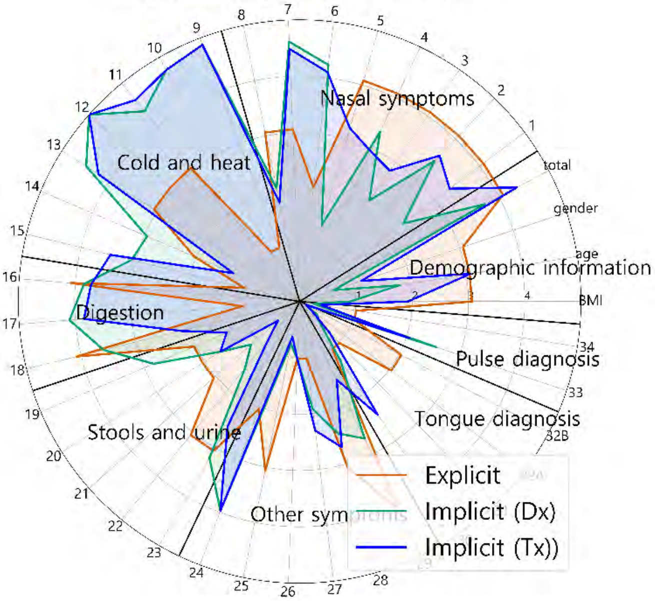


Comparison of explicit and implicit importance scores (average results)

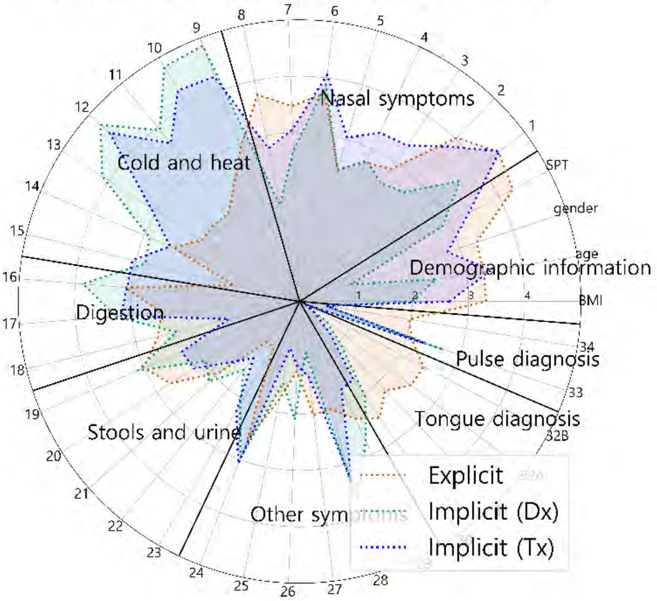

### Doctor #6

### Supplementary sheet

(S1) Intra-individual comparison of explicit importance score and implicit importance score for diagnosis

(S2) Intra-individual comparison of explicit importance score and implicit importance score for prescription

S1 and S2 were represented as scatter plots. The correlation statistics between the explicit and implicit scores were calculated using Spearman's rank correlation coefficient test. The X-axis represents the normalized value of the implicit importance and the Y-axis represents the normalized value of the explicit importance.

| Result of Spearman test (S1) | | |
|---|---|---|
| | statistic | 0.152 |
| | p-value | 0.357 |

| Result of Spearman test (S2) | | |
|---|---|---|
| | statistic | 0.189 |
| | p-value | 0.250 |

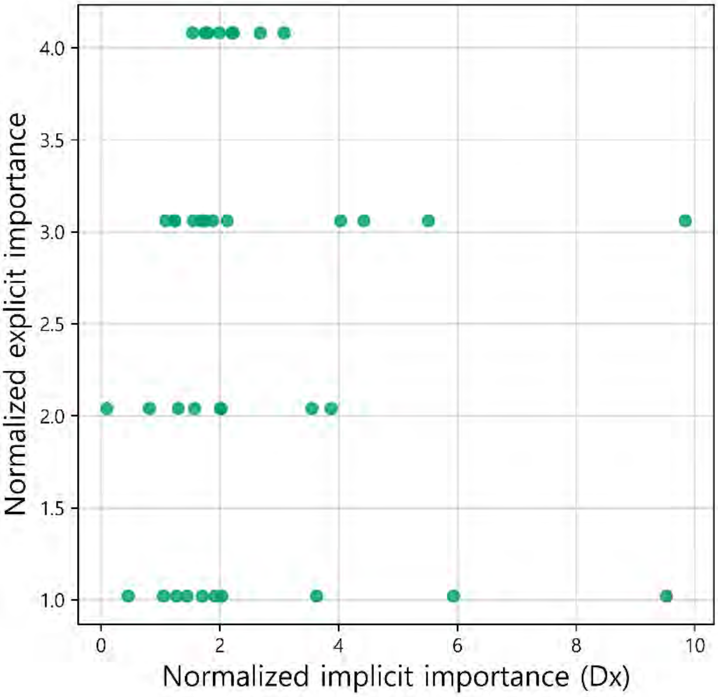


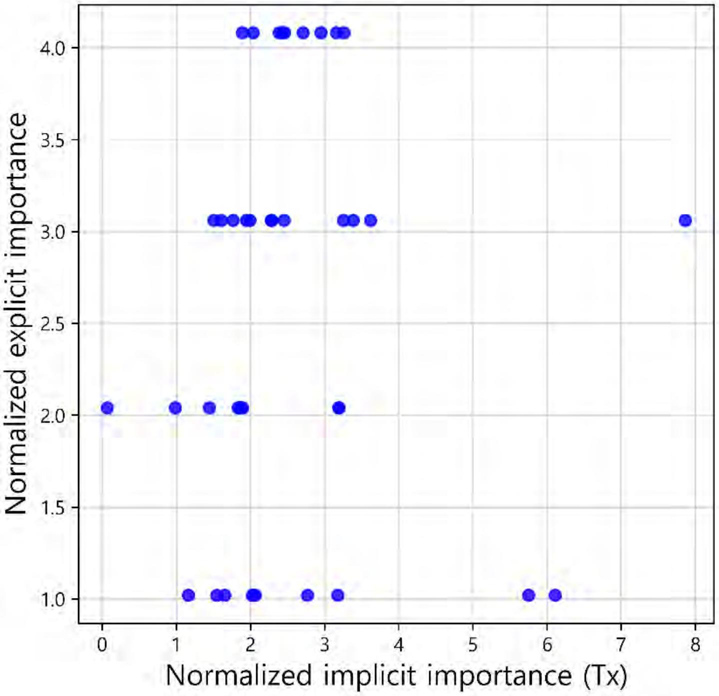


(S3) Comparison of the individual and average explicit importance scores

S3 was constructed to compare the average importance score of the doctors as a group with the individual doctor's importance score. Individual results from each doctor and the average of all doctors' results are expressed as a bar graph and a dotted line, respectively. The graphs are sorted in descending order according to the importance score of the average results.

| Result of Spearman test | |
|---|---|
| statistic | 0.406 |
| p-value | 0.010 |

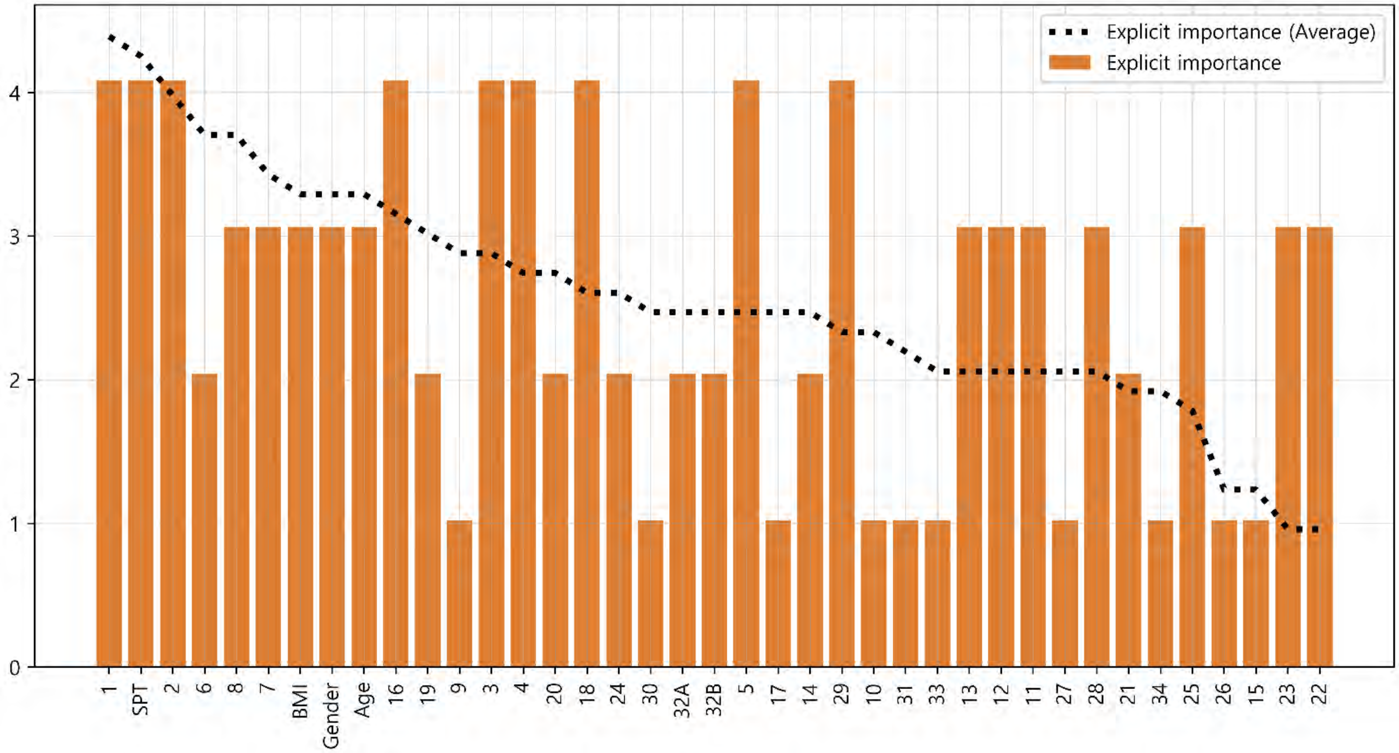

### Doctor #6

### Supplementary sheet

#### (S4) Comparison of the individual and average implicit importance scores for diagnosis

S4 was constructed to compare the average importance score of the doctors as a group with the individual doctor's importance score. Individual results from each doctor and the average of all doctors' results are expressed as a bar graph and a dotted line, respectively. The graphs are sorted in descending order according to the importance score of the average results.

| Result of Spearman test | |
|---|---|
| statistic | 0.873 |
| p-value | < 0.001 |

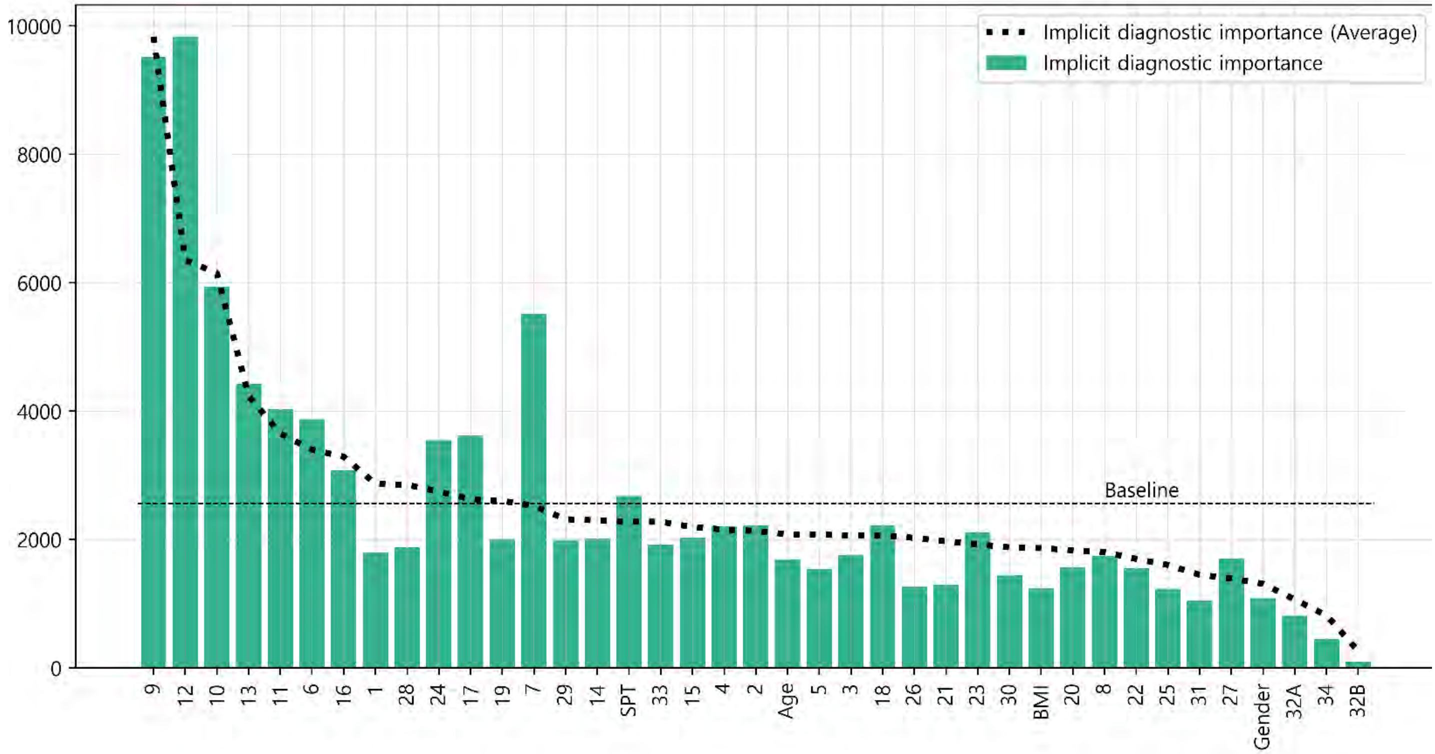


#### (S5) Comparison of the individual and average implicit importance scores for prescription

S5 was constructed to compare the average importance score of the doctors as a group with the individual doctor's importance score. Individual results from each doctor and the average of all doctors' results are expressed as a bar graph and a dotted line, respectively. The graphs are sorted in descending order according to the importance score of the average results.

| Result of Spearman test | |
|---|---|
| statistic | 0.806 |
| p-value | < 0.001 |

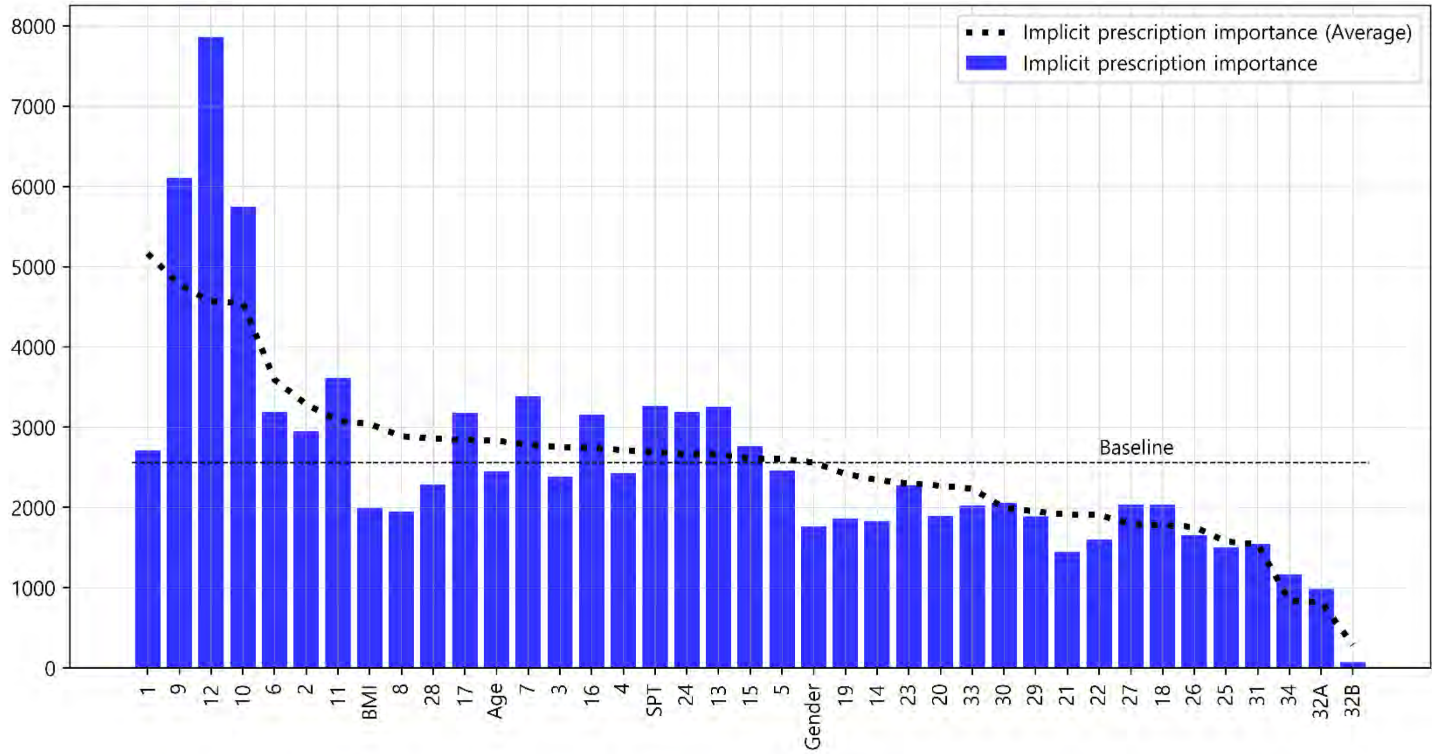

### Doctor #7

#### Glossary & Tips

1. Explicit knowledge & implicit knowledge

- Knowledge can be classified into two different categories: explicit and implicit knowledge. Explicit knowledge refers to knowledge that can be expressed in words and in a form that can be codified. In contrast, implicit knowledge refers to knowledge that an individual obtains through experience but does not convey to others.
- In this analysis, the explicit importance score was defined for the features obtained from doctors' label indicating the degree of importance of each symptom for diagnosing and treating AR patients. Implicit importance was defined based on the weights of the features learned by the machine learning algorithm when the learning algorithm effectively reproduced the doctor's decision-making process.

2. Baseline

- The baseline is the average value of the implicit importance score of individual results.

3. The explicit importance score, implicit importance score for diagnosis, and implicit importance score for prescription are visualized in orange, cyan, and blue, respectively. Individual results from each doctor and the average of all doctors' results are expressed as a bar graph and a dotted line, respectively

4. The statistic analysis was calculated by Spearman's rank correlation coefficient (p-value < 0.05).

(M1) Comparison of the individual and average explicit and implicit importance scores

To provide an overview of the differences in all importance scores, M1 was visualized with line graphs that consisted of all six scores. Scores were sorted in descending order according to the average explicit importance score of average results

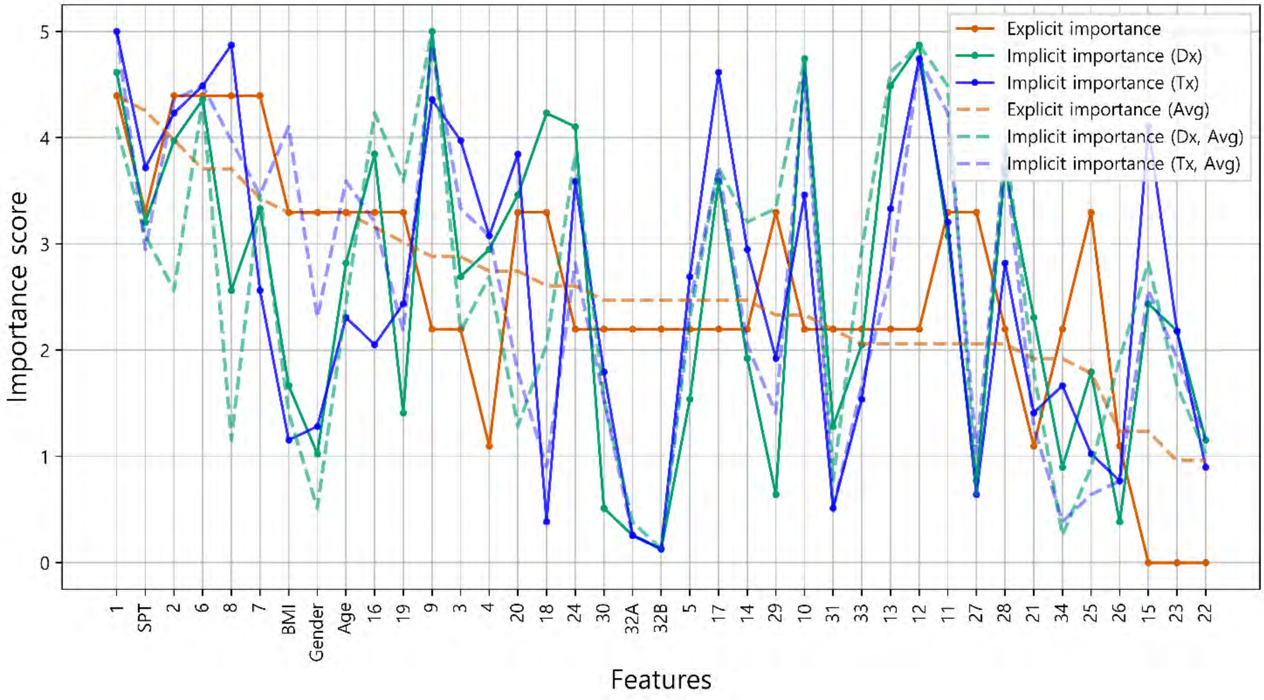


(M2) Intra-individual correlation of explicit and implicit importance

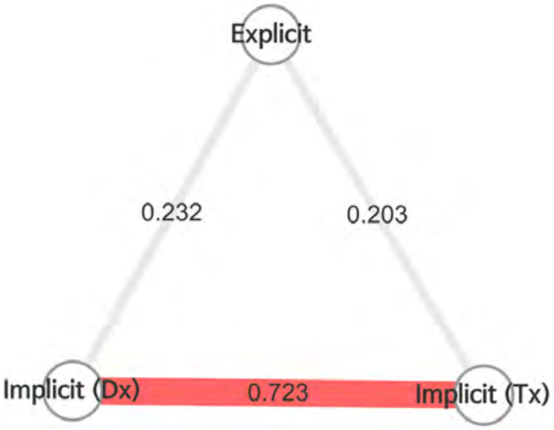


- To examine the overall correlation between the explicit and implicit knowledge of individuals, M2 was presented as nodes and edges, which represented the types of importance scores and the correlations between importance scores, respectively.
- Color of edge : the results of the Bonferroni post hoc test
  - Dark red: statistically significant
  - light red: not significant, but p-value<0.05
  - grey : p-value>0.05
- Thickness of edge (statistic): the thicker the edge, the greater the correlation.

- It is analyzed that there is no correlation between the explicit importance and implicit diagnostic importance (ρ = 0.232, p-value = 0.154). Explicit importance and implicit prescription importance have been identified as having no correlation (ρ = 0.203, p-value = 0.215).

### Doctor #7 (M3) Radar chart analysis

All the features used in the analysis were classified into eight categories and visualized as a radar chart to provide the information about the feature categories that the doctor referred to for treatment. The categories consisted of demographic features of the patient, nose symptoms, cold-hot, digestion, feces and urine, tongue diagnosis, pulse diagnosis and other symptoms

- The explicit importance score, implicit importance score for diagnosis, and implicit importance score for prescription are visualized in orange, cyan, and blue, respectively.
- Individual results from each doctor and the average of all doctors' results are represented as a solid line and a dotted line, respectively.

Comparison of Doctor 7 and average explicit importance scores

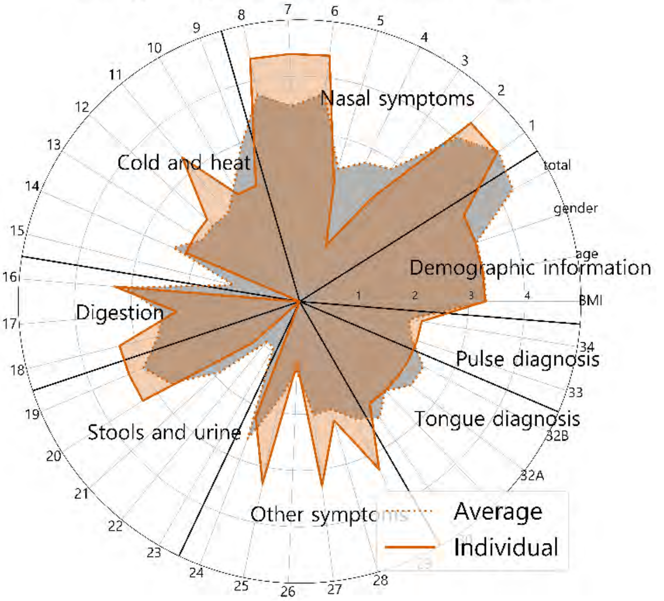


Comparison of Doctor 7 and average implicit importance scores for diagnosis

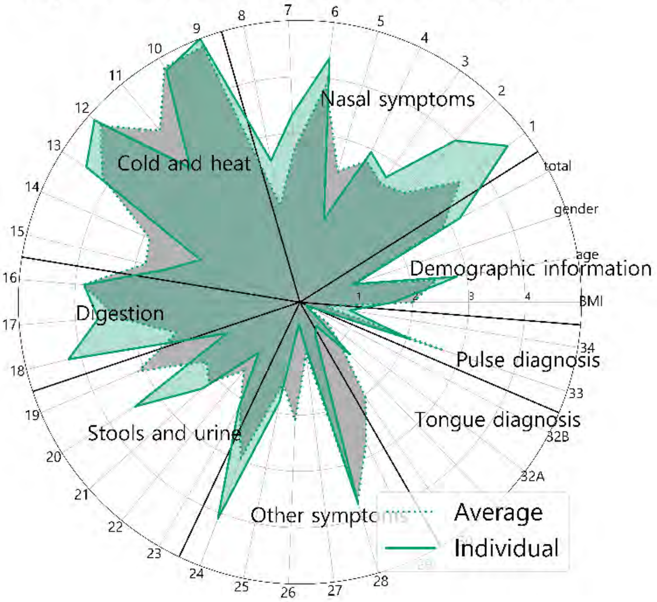


Comparison of Doctor 7 and average implicit importance scores for prescription

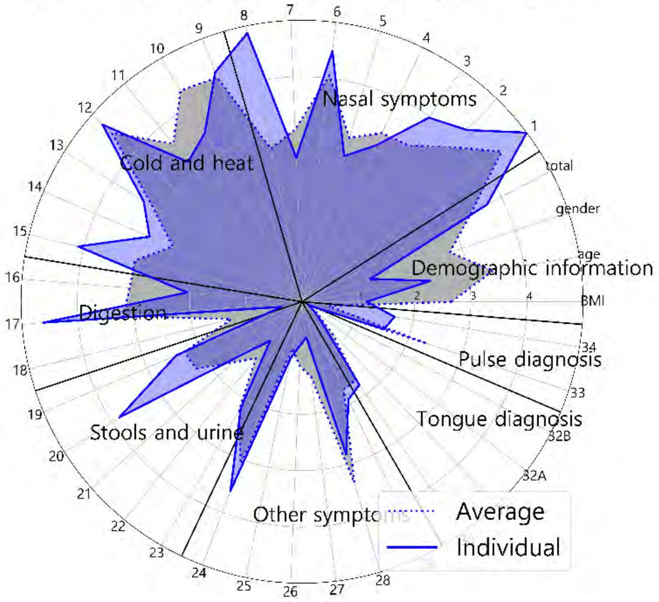


Comparison of explicit and implicit importance scores

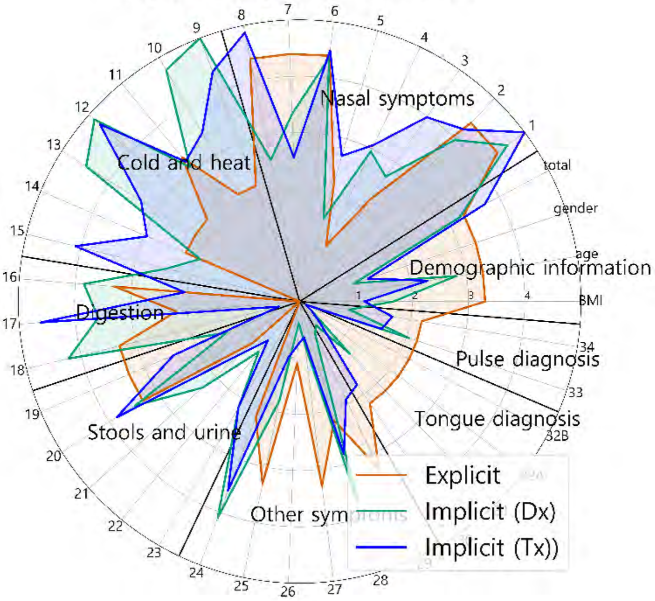


Comparison of explicit and implicit importance scores (average results)

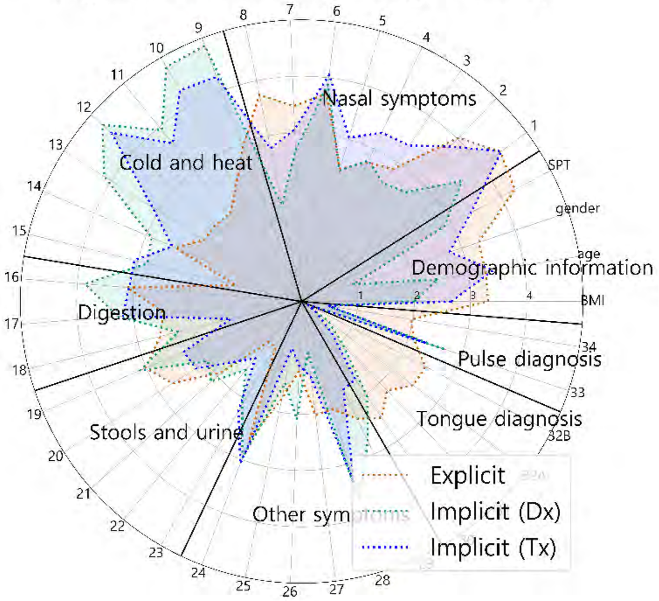

### Doctor #7

### Supplementary sheet

(S1) Intra-individual comparison of explicit importance score and implicit importance score for diagnosis

(S2) Intra-individual comparison of explicit importance score and implicit importance score for prescription

S1 and S2 were represented as scatter plots. The correlation statistics between the explicit and implicit scores were calculated using Spearman's rank correlation coefficient test. The X-axis represents the normalized value of the implicit importance and the Y-axis represents the normalized value of the explicit importance.

| Result of Spearman test | statistic | 0.232 |
|---|---|---|
| | p-value | 0.154 |

| Result of Spearman test | statistic | 0.203 |
|---|---|---|
| | p-value | 0.215 |

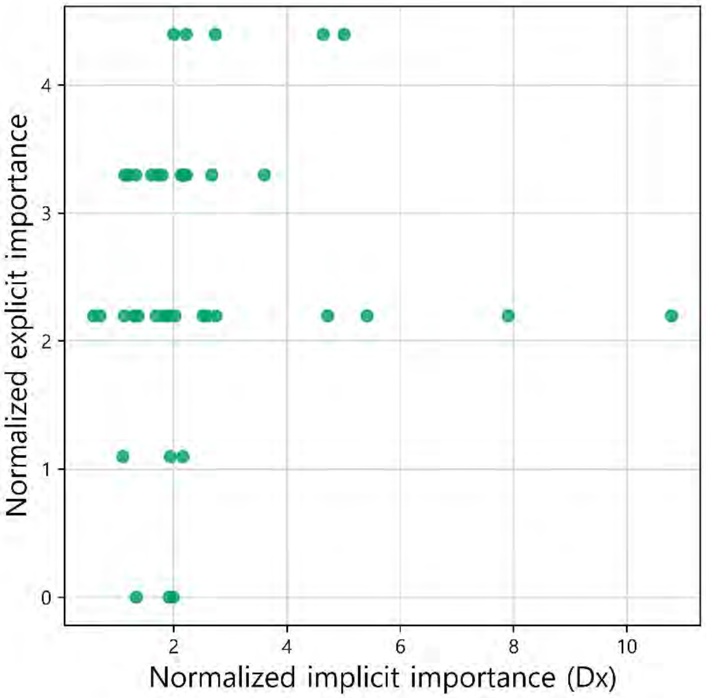


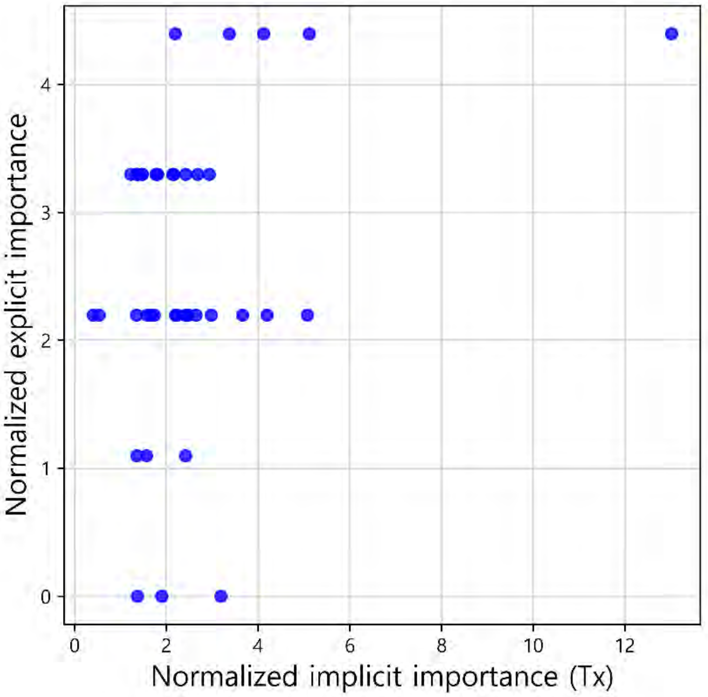


(S3) Comparison of the individual and average explicit importance scores

S3 was constructed to compare the average importance score of the doctors as a group with the individual doctor's importance score. Individual results from each doctor and the average of all doctors' results are expressed as a bar graph and a dotted line, respectively. The graphs are sorted in descending order according to the importance score of the average results.

| Result of Spearman test | |
|---|---|
| statistic | 0.714 |
| p-value | < 0.001 |

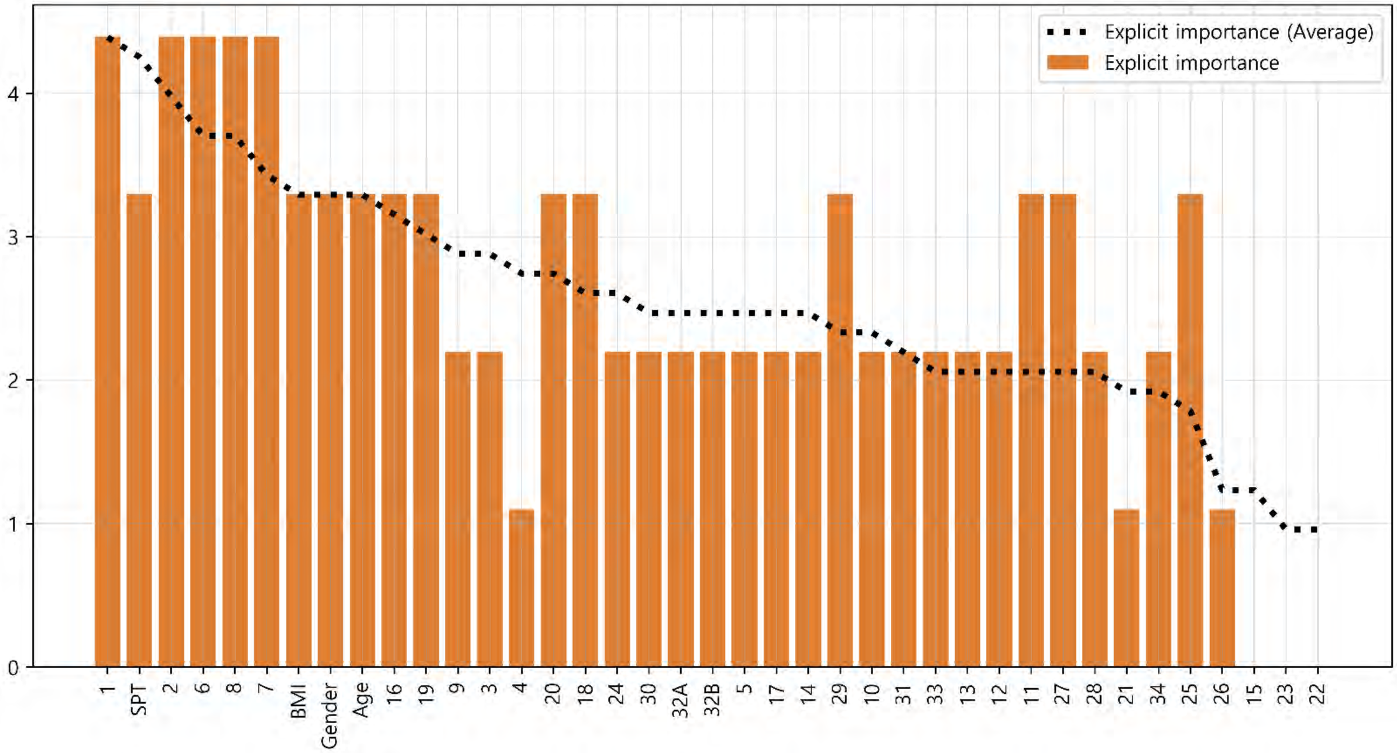

#### Doctor #7



##### (S4) Comparison of the individual and average implicit importance scores for diagnosis

S4 was constructed to compare the average importance score of the doctors as a group with the individual doctor's importance score. Individual results from each doctor and the average of all doctors' results are expressed as a bar graph and a dotted line, respectively. The graphs are sorted in descending order according to the importance score of the average results.

| Result of Spearman test | |
|---|---|
| statistic | 0.769 |
| p-value | < 0.001 |

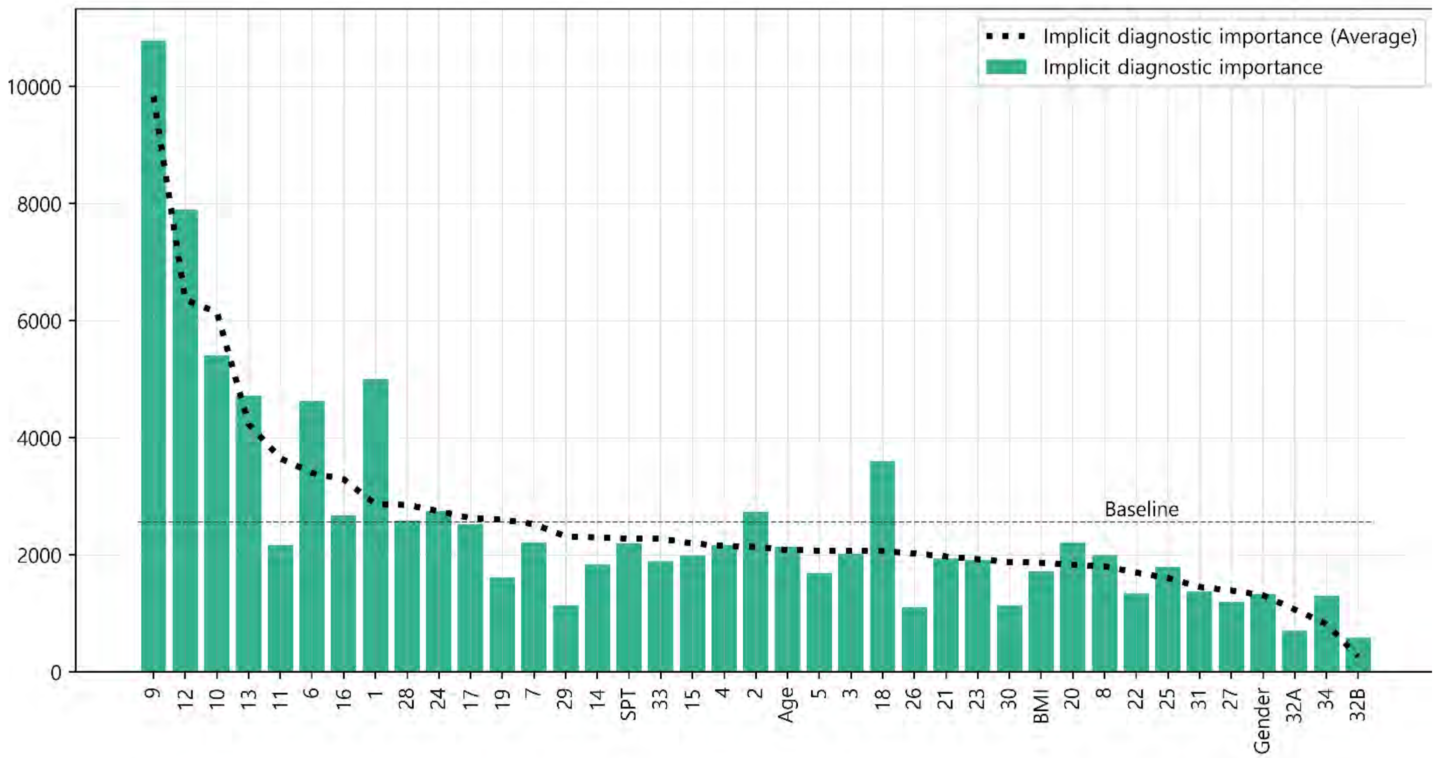


##### (S5) Comparison of the individual and average implicit importance scores for prescription

S5 was constructed to compare the average importance score of the doctors as a group with the individual doctor's importance score. Individual results from each doctor and the average of all doctors' results are expressed as a bar graph and a dotted line, respectively. The graphs are sorted in descending order according to the importance score of the average results.

| Result of Spearman test | |
|---|---|
| statistic | 0.809 |
| p-value | < 0.001 |

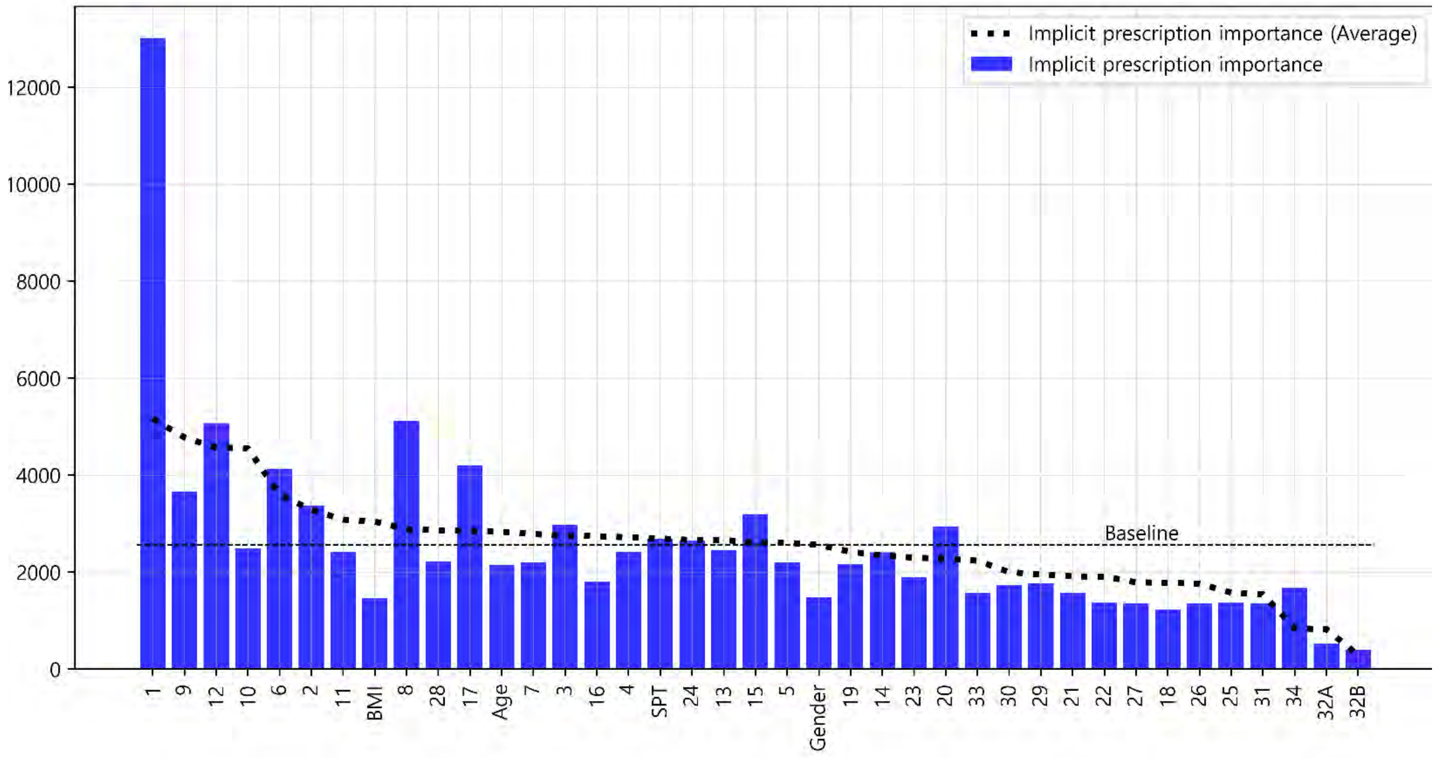

### Doctor #8

#### Glossary & Tips

1. Explicit knowledge & implicit knowledge

- Knowledge can be classified into two different categories: explicit and implicit knowledge. Explicit knowledge refers to knowledge that can be expressed in words and in a form that can be codified. In contrast, implicit knowledge refers to knowledge that an individual obtains through experience but does not convey to others.
- In this analysis, the explicit importance score was defined for the features obtained from doctors' label indicating the degree of importance of each symptom for diagnosing and treating AR patients. Implicit importance was defined based on the weights of the features learned by the machine learning algorithm when the learning algorithm effectively reproduced the doctor's decision-making process.

2. Baseline

- The baseline is the average value of the implicit importance score of individual results.

3. The explicit importance score, implicit importance score for diagnosis, and implicit importance score for prescription are visualized in orange, cyan, and blue, respectively. Individual results from each doctor and the average of all doctors' results are expressed as a bar graph and a dotted line, respectively

4. The statistic analysis was calculated by Spearman's rank correlation coefficient (p-value < 0.05).

(M1) Comparison of the individual and average explicit and implicit importance scores

To provide an overview of the differences in all importance scores, M1 was visualized with line graphs that consisted of all six scores. Scores were sorted in descending order according to the average explicit importance score of average results

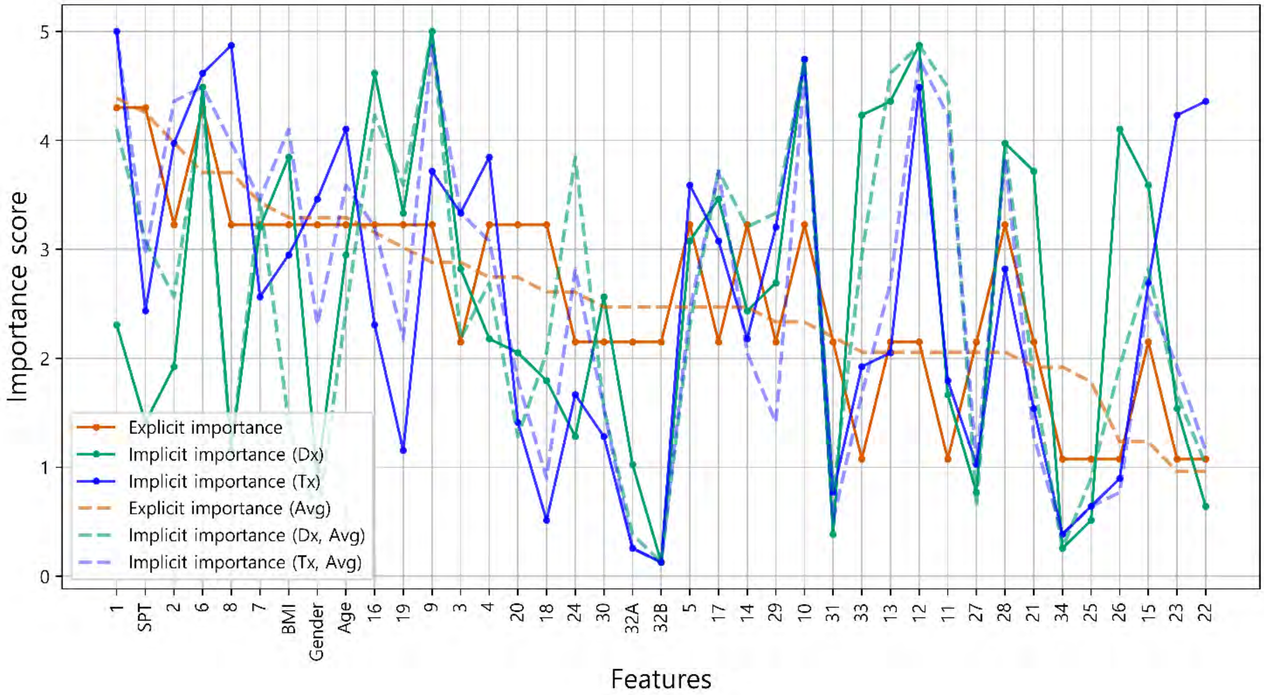


(M2) Intra-individual correlation of explicit and implicit importance

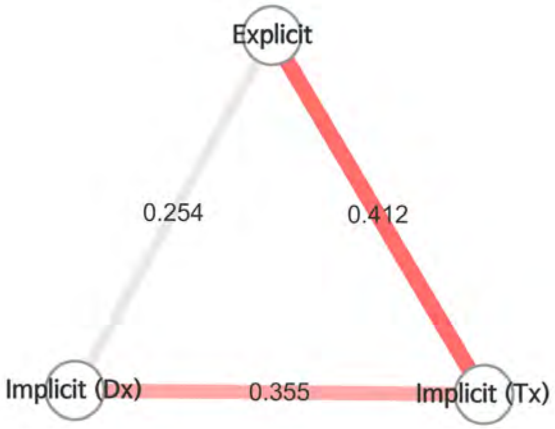


- To examine the overall correlation between the explicit and implicit knowledge of individuals, M2 was presented as nodes and edges, which represented the types of importance scores and the correlations between importance scores, respectively.
- Color of edge : the results of the Bonferroni post hoc test
  - Dark red: statistically significant
  - light red: not significant, but p-value<0.05
  - grey : p-value>0.05
- Thickness of edge (statistic): the thicker the edge, the greater the correlation.

- It is analyzed that there is no correlation between the explicit importance and implicit diagnostic importance ($\rho$ = 0.254, p-value = 0.119). Explicit importance and implicit prescription importance have been identified as having a weak correlation ($\rho$ = 0.412, p-value = 0.009).

### Doctor #8

(M3) Radar chart analysis

All the features used in the analysis were classified into eight categories and visualized as a radar chart to provide the information about the feature categories that the doctor referred to for treatment. The categories consisted of demographic features of the patient, nose symptoms, cold-hot, digestion, feces and urine, tongue diagnosis, pulse diagnosis and other symptoms

- The explicit importance score, implicit importance score for diagnosis, and implicit importance score for prescription are visualized in orange, cyan, and blue, respectively.
- Individual results from each doctor and the average of all doctors' results are represented as a solid line and a dotted line, respectively.

Comparison of Doctor 8 and average explicit importance scores

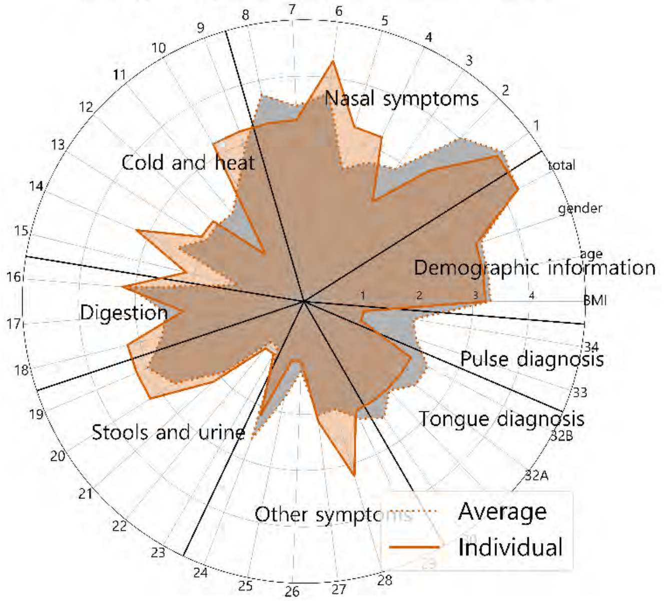


Comparison of Doctor 8 and average implicit importance scores for diagnosis

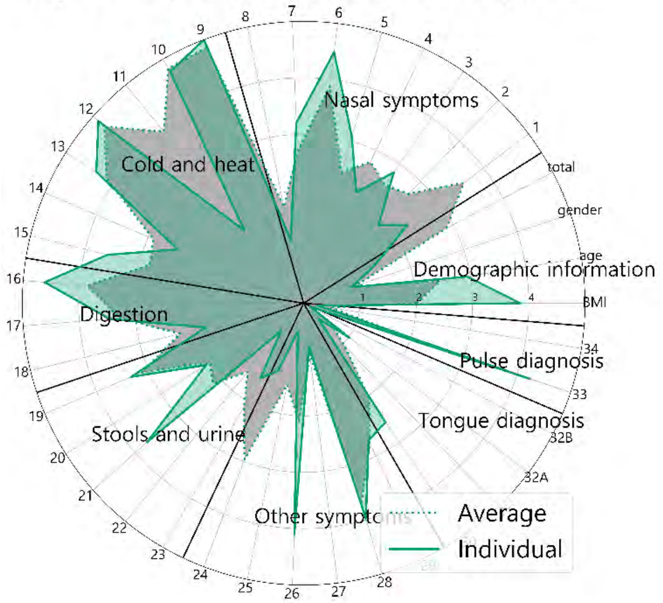


Comparison of Doctor 8 and average implicit importance scores for prescription

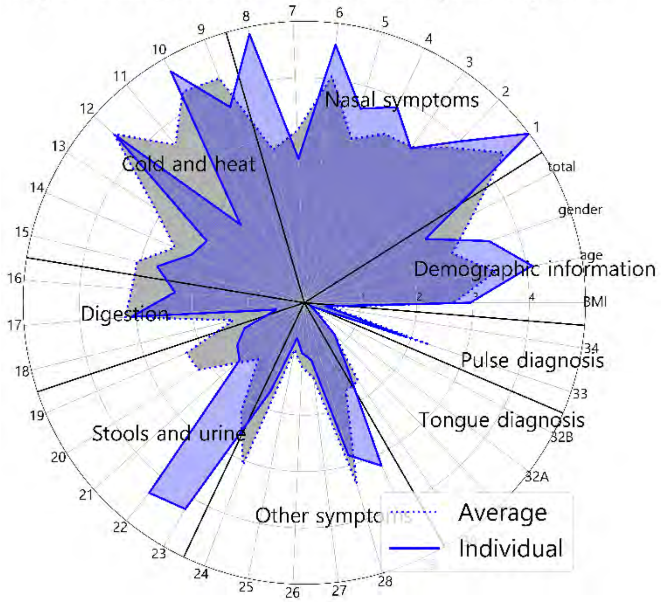


Comparison of explicit and implicit importance scores

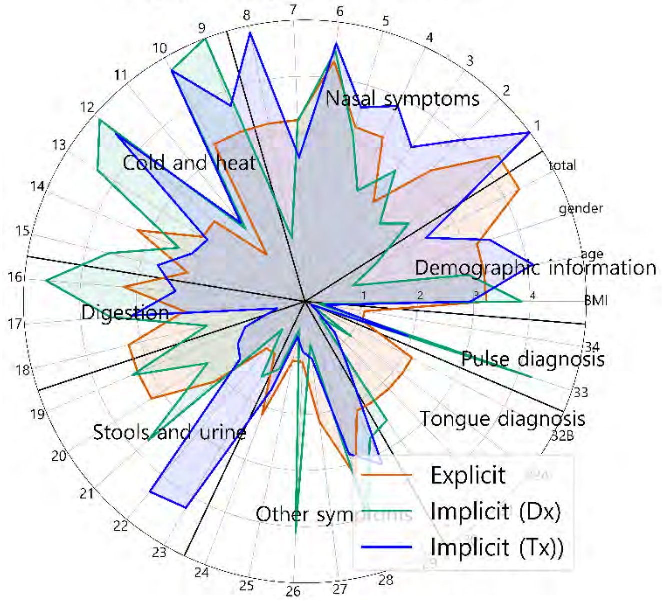


Comparison of explicit and implicit importance scores (average results)

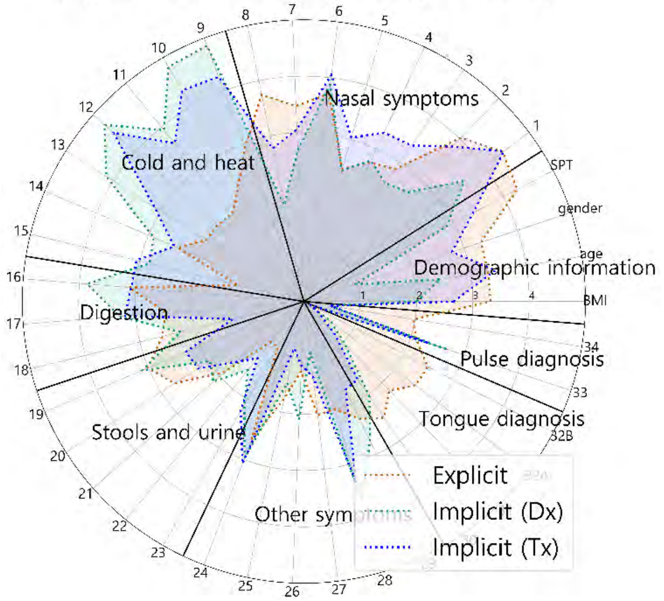

### Doctor #8

### Supplementary sheet

(S1) Intra-individual comparison of explicit importance score and implicit importance score for diagnosis

(S2) Intra-individual comparison of explicit importance score and implicit importance score for prescription

S1 and S2 were represented as scatter plots. The correlation statistics between the explicit and implicit scores were calculated using Spearman's rank correlation coefficient test. The X-axis represents the normalized value of the implicit importance and the Y-axis represents the normalized value of the explicit importance.

| Result of Spearman test | statistic | 0.254 |
| --- | --- | --- |
| | p-value | 0.119 |

| Result of Spearman test | statistic | 0.412 |
| --- | --- | --- |
| | p-value | 0.009 |

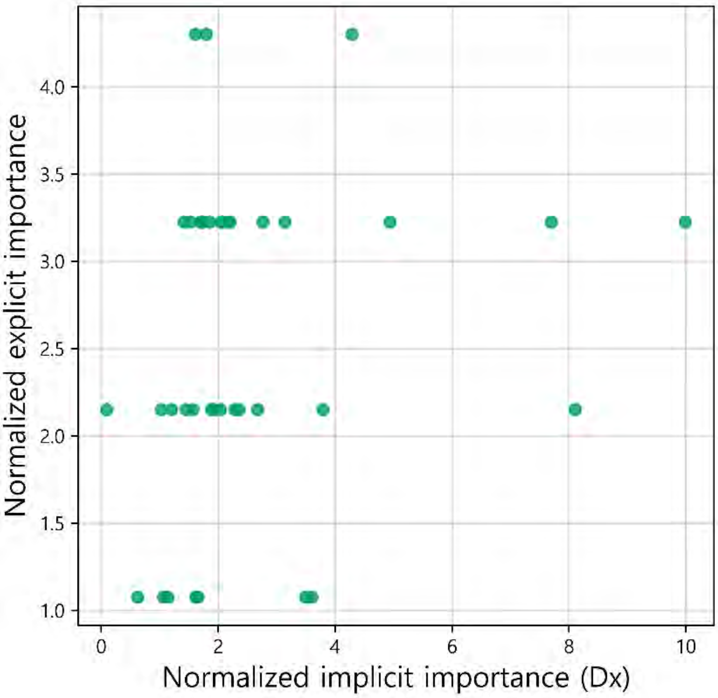


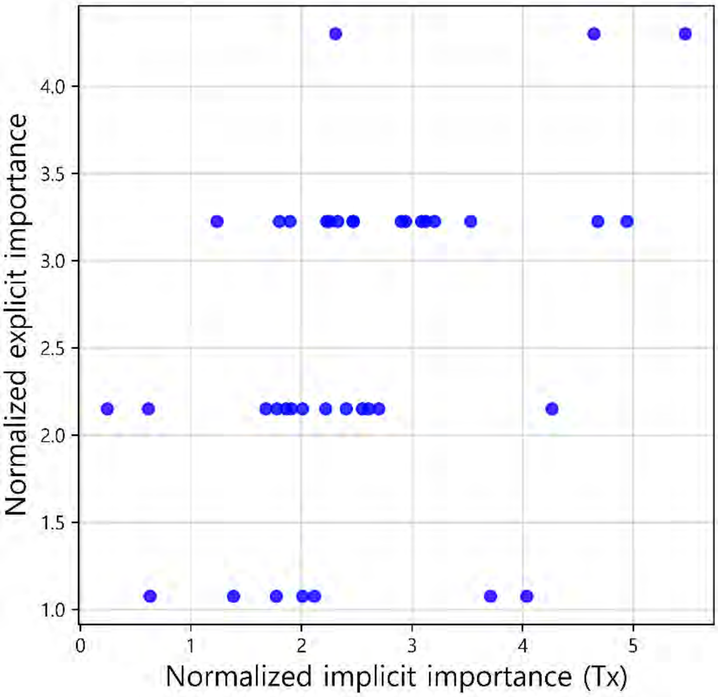


(S3) Comparison of the individual and average explicit importance scores

S3 was constructed to compare the average importance score of the doctors as a group with the individual doctor's importance score. Individual results from each doctor and the average of all doctors' results are expressed as a bar graph and a dotted line, respectively. The graphs are sorted in descending order according to the importance score of the average results.

| Result of Spearman test | |
| --- | --- |
| statistic | 0.829 |
| p-value | < 0.001 |

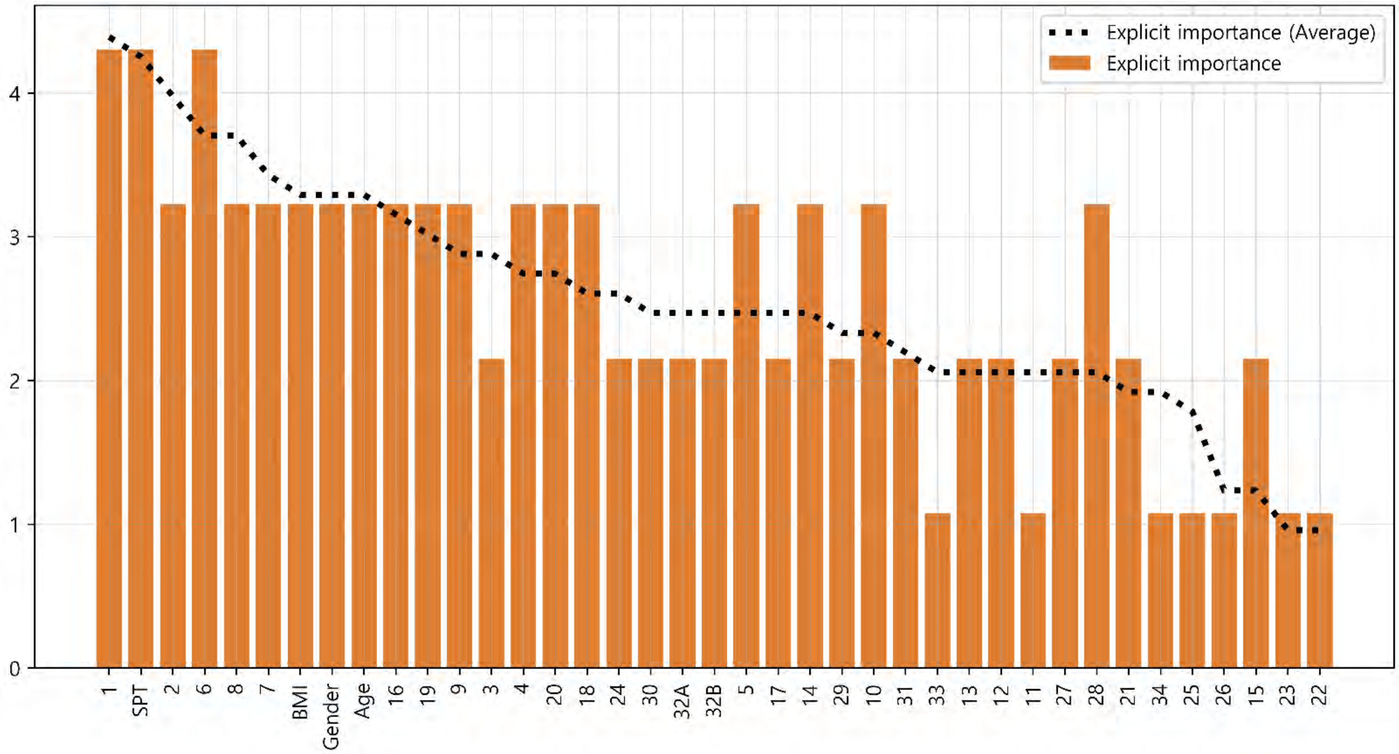

### Doctor #8



#### (S4) Comparison of the individual and average implicit importance scores for diagnosis

S4 was constructed to compare the average importance score of the doctors as a group with the individual doctor's importance score. Individual results from each doctor and the average of all doctors' results are expressed as a bar graph and a dotted line, respectively. The graphs are sorted in descending order according to the importance score of the average results.

| Result of Spearman test | |
|---|---|
| statistic | 0.734 |
| p-value | < 0.001 |

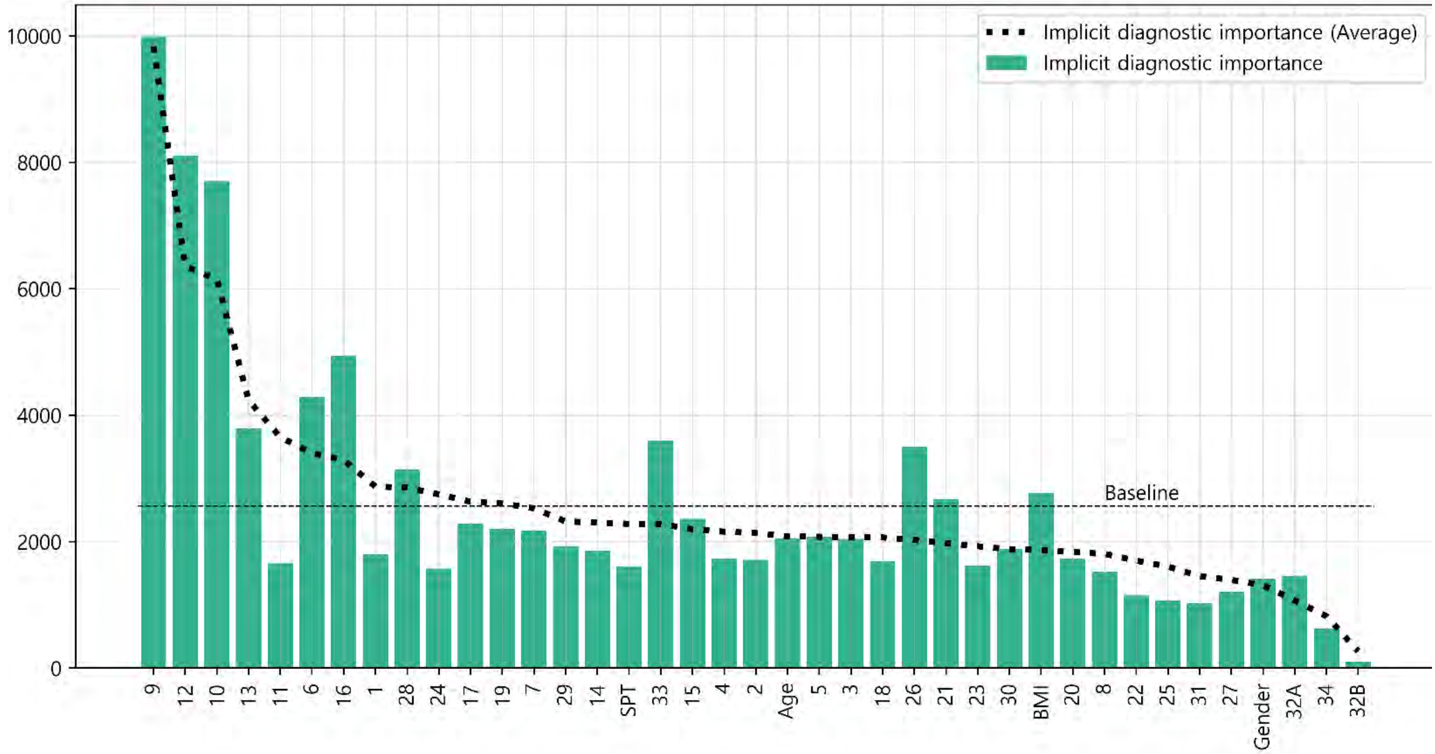


#### (S5) Comparison of the individual and average implicit importance scores for prescription

S5 was constructed to compare the average importance score of the doctors as a group with the individual doctor's importance score. Individual results from each doctor and the average of all doctors' results are expressed as a bar graph and a dotted line, respectively. The graphs are sorted in descending order according to the importance score of the average results.

| Result of Spearman test | |
|---|---|
| statistic | 0.762 |
| p-value | < 0.001 |

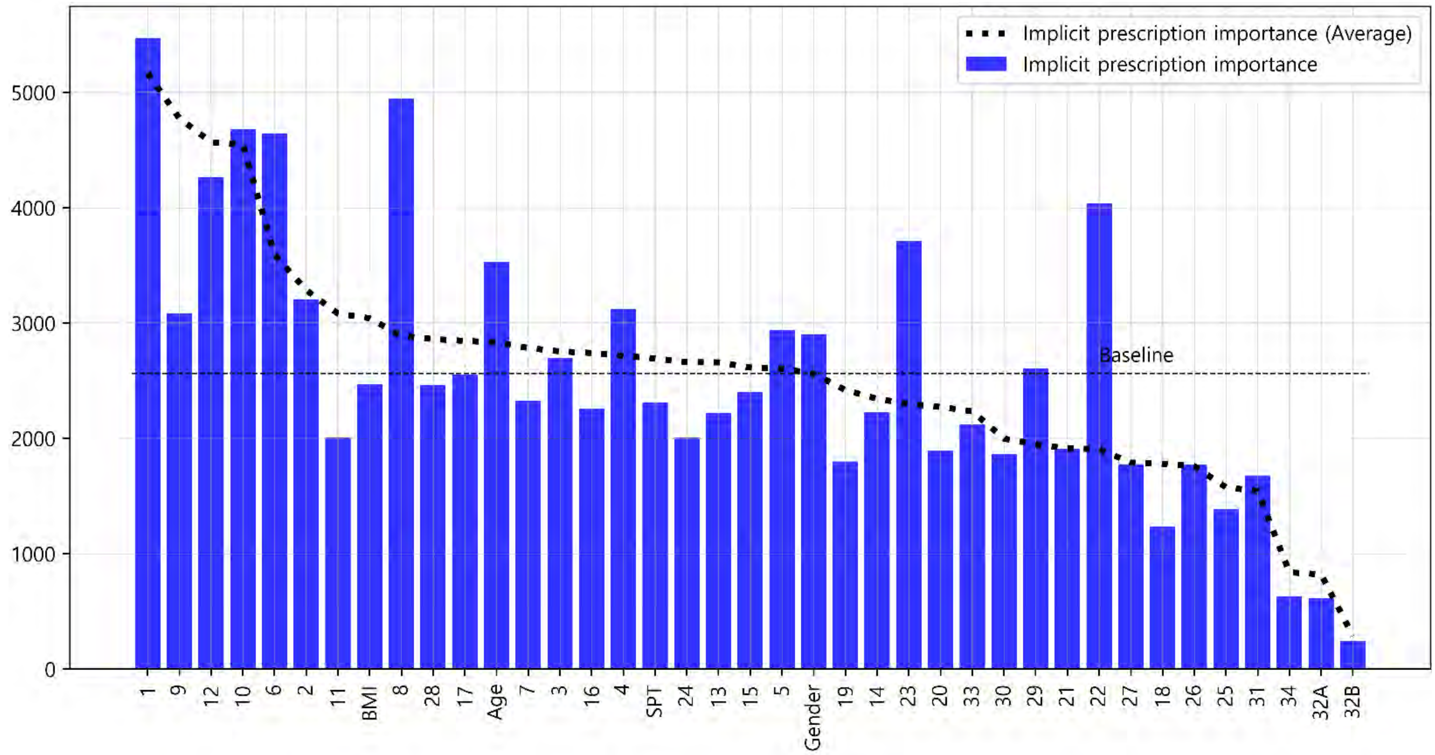
